# Valid inference for machine learning-assisted GWAS

**DOI:** 10.1101/2024.01.03.24300779

**Authors:** Jiacheng Miao, Yixuan Wu, Zhongxuan Sun, Xinran Miao, Tianyuan Lu, Jiwei Zhao, Qiongshi Lu

## Abstract

Machine learning (ML) has revolutionized analytical strategies in almost all scientific disciplines including human genetics and genomics. Due to challenges in sample collection and precise phenotyping, ML-assisted genome-wide association study (GWAS) which uses sophisticated ML to impute phenotypes and then performs GWAS on imputed outcomes has quickly gained popularity in complex trait genetics research. However, the validity of associations identified from ML-assisted GWAS has not been carefully evaluated. In this study, we report pervasive risks for false positive associations in ML-assisted GWAS, and introduce POP-GWAS, a novel statistical framework that reimagines GWAS on ML-imputed outcomes. POP-GWAS provides valid statistical inference irrespective of the quality of imputation or variables and algorithms used for imputation. It also only requires GWAS summary statistics as input. We employed POP-GWAS to perform the largest GWAS of bone mineral density (BMD) derived from dual-energy X-ray absorptiometry imaging at 14 skeletal sites, identifying 89 novel loci reaching genome-wide significance and revealing skeletal site-specific genetic architecture of BMD. Our framework may fundamentally reshape the analytical strategies in future ML-assisted GWAS.

## Introduction

Genome-wide association study (GWAS) is a powerful tool for identifying genetic variants associated with complex human traits^1^. However, even in the era of biobank cohorts with tens of thousands of individuals, high-quality phenotype data is often lacking due to the costly technology for phenotypic measurement, invasive procedure for sample collection, or a lack of commitment to study participation^2-7^. These challenges severely reduce the statistical power of GWAS on many valuable phenotypes, compromising genetic discoveries and efforts to uncover new therapeutic targets^3^. To overcome these issues, an emerging solution quickly gaining traction in the field is to use machine learning (ML) to impute missing phenotypes based on observed variables, then perform subsequent GWAS on the imputed phenotypes. We refer to this design as ML-assisted GWAS. Many recent studies have adopted this design and demonstrated its superior statistical power compared to GWAS based on measured phenotypes alone^2-6,8,9^. However, despite its growing popularity, the validity of associations identified in ML-assisted GWAS has not been carefully evaluated. In this paper, we have two main objectives. First, through extensive theoretical analysis, simulation studies, and benchmarking in large biobank samples, we alert the field about the risk of having pervasive false-positive associations using current ML-assisted GWAS strategies. Second, we introduce a novel and principled statistical framework for ML-assisted GWAS analysis with no assumption on the degree of phenotype missingness, accuracy of phenotype imputation, and choice of ML algorithm.

A common use case for ML-assisted GWAS is when a gold-standard phenotype is only measured in a small fraction of genomic samples which we refer to as labeled data. The remaining (unlabeled) samples do not have this phenotype measured. Prominent genetic datasets, such as the UK Biobank (UKB) and All of Us, often have incomplete phenotypic data^10^. For example, as of November of 2023, proteomics^11^, brain magnetic resonance imaging (MRI)^12^, heart MRI^13^, dual-energy X-ray absorptiometry (DXA) imaging^6^, electrocardiogram^14^, and metabolomics^15^ data in UKB have missing rates ranging from 45% to 94%. Similarly, All of Us has a missing rate of 96% for phenotypes in the Labs & Measurements category of the electronic health record (**Supplementary Figure 1**). Fortunately, the past decade has seen significant advances in the development of sophisticated ML algorithms^16-18^ and collection of extensive demographic and clinical information in large biobanks. These innovations have enabled phenotype imputation in unlabeled samples, fostering a rapidly growing interest in using ML-assisted GWAS to increase statistical power.

Several approaches have been introduced to carry out ML-assisted GWAS. Some studies choose to impute the phenotype in unlabeled samples, then perform a GWAS on it using unlabeled samples alone^2,8^. An alternative approach merges the imputed phenotype in unlabeled samples with the measured phenotype in labeled samples, and performs GWAS on the combined dataset^2^. Other studies perform phenotypic imputation in both labeled and unlabeled samples, and follow with a GWAS on the imputed phenotype using the whole sample^4,5^. All these approaches treat the imputed phenotype as observed, ignoring the uncertainty in imputation. The validity of their results is often justified based on heuristic, *ad hoc* analysis such as showing comparable effect sizes or a moderate genetic correlation between GWAS of imputed and observed phenotypes^2,4,5,8^ or efforts to account for phenotypic heterogeneity during meta-analysis^2,19,20^. Importantly, there is a general lack of understanding of how these methods compare to each other, particularly regarding whether they in fact estimate genetic effect on the gold-standard phenotype which is the intended parameter of interest. As we demonstrate below, existing approaches do not ensure the validity of association findings.

Here, we reveal major limitations in current ML-assisted GWAS approaches, and introduce a statistical framework named **Po**st-**p**rediction **GWAS** (POP-GWAS) for valid and powerful inference in ML-assisted GWAS. Our method provides unbiased estimates and well-calibrated type-I error, is universally more powerful than conventional GWAS on the observed phenotype, and has minimal assumption on the variables used for imputation, quality of imputation, and choice of prediction algorithm. Furthermore, it only requires GWAS summary statistics as input. We showcase the performance of POP-GWAS in an extensive case study of bone mineral density (BMD) across 14 skeletal sites.

## Results

### Conventional GWAS on imputed phenotypes may have pervasive false positive associations

We begin by assessing the validity of conventional ML-assisted GWAS using real-data benchmarking. We carried out a GWAS on type 2 diabetes (T2D) using 408,325 individuals (18,147 cases and 390,178 controls) with European ancestry in UKB. We treated associations in this GWAS as ground truth T2D associations. Next, we randomly split the full dataset into two subsamples with 25% and 75% of all individuals. We trained the SoftImpute^21^ algorithm for T2D imputation on the 25% subsample (**Methods**). Then, we masked real T2D phenotypes in the 75% subsample and applied this model to impute T2D. We achieved reasonable imputation quality (correlation of measured and imputed phenotypes: r = 0.6) that is comparable to what has been reported in published studies^2,3^. We then performed a GWAS on imputed T2D liability in the 75% subsample. We calculated replication failure rate, defined as the proportion of independent significant loci (P < 5e-8) that failed to replicate in the ground truth GWAS (P > 5e-8 or with flipped effect direction). Strikingly, GWAS on imputed T2D had a high replication failure rate of 81% (**Figure 1a**). Even when we relaxed the replication P-value threshold to 5e-6 and 0.05, the replication failure rate remained high (i.e., 69% and 17%, respectively). We further sought replication using the DIAMENTE study – the largest T2D case-control GWAS with 74,124 cases and 824,006 controls of European ancestry^22^. We once again observed high replication failure rates of 48%, 39%, and 16% at P-value thresholds of 5e-8, 5e-6, and 0.05, respectively.

**Figure 1.**
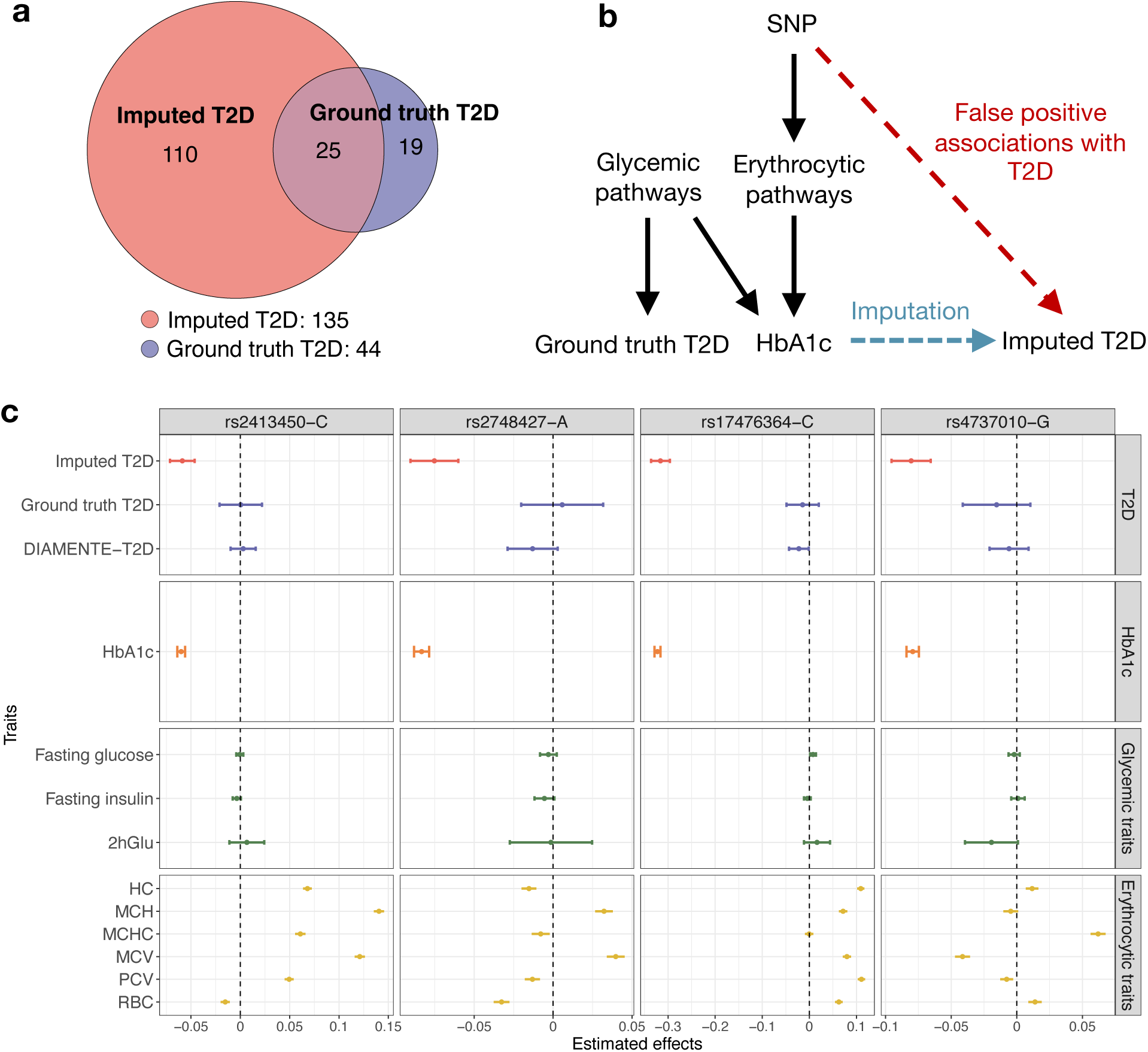
Pervasive false positive associations in the GWAS on imputed T2D. **(a)** Venn diagram comparing the number of independent loci identified by GWAS of imputed and ground truth T2D **(b)** The chart displays an example for how a false positive SNP in the GWAS on imputed T2D may be involved in glycemic and erythrocytic pathways which lead to T2D and HbA1c associations. SNPs can have false positive associations with imputed T2D due to their effects on HbA1c through erythrocytic pathways. **(c)** Estimated effects of four SNPs on T2D, HbA1c, glycemic traits, and erythrocytic traits. The vertical dashed line at 0 serves as a reference for no effect. Error bars show the 95% confidence intervals. The DIAMENTE-T2D is the largest T2D case-control GWAS to date. Abbreviations: 2hGlu (2-h glucose after an oral glucose challenge), HC (haemoglobin concentration), MCH (mean corpuscular haemoglobin), MCHC (mean corpuscular haemoglobin concentration), MCV (mean corpuscular volume), PCV (haematocrit percentage), RBC (red blood cell count),

Next, we investigated why many loci identified using the imputed phenotype could not be replicated. Unsurprisingly, we found that hemoglobin A1C (HbA1c) is the strongest predictor for T2D in the imputation model (**Supplementary Table 1**). Although elevated HbA1c is one of the clinically used diagnostic criteria for T2D, it is known that genetic variants may affect HbA1c levels through both glycemic and non-glycemic pathways^23-25^ (**Figure 1b**). Glycemic pathways involve mechanisms that affect blood glucose levels, which are central to the development and management of T2D^26^. Non-glycemic pathways, on the other hand, influence HbA1c levels through factors less relevant to glycemia, such as the lifespan or properties of red blood cells (erythrocytes)^26^. Therefore, GWAS on imputed T2D identified many non-glycemic variants for HbA1c that are not associated with T2D risk. These associations failed to replicate in the ground truth T2D GWAS. For example, single-nucleotide polymorphisms (SNPs) rs2413450, rs2748427, rs17476364, and rs4737010 are all significantly associated with imputed T2D but not ground truth T2D in UKB or the DIAMENTE study. We looked up their associations with three glycemic traits^27^ (i.e., glucose, fasting glucose, and fasting insulin levels) and six erythrocyte traits^24^ (i.e., haemoglobin concentration, mean corpuscular haemoglobin, mean corpuscular haemoglobin concentration, mean corpuscular volume, haematocrit percentage, and red blood cell count). All four SNPs showed substantial associations with erythrocyte traits but not glycemic traits (**Figure 1c**), which explains their strong associations with HbA1c and imputed T2D but not with ground truth T2D risk. These false positive associations were identified despite good imputation quality and a near-perfect genetic correlation (cor = 0.99, se = 0.04) between the imputed and ground truth GWAS. This demonstrates that high imputation accuracy and genetic correlation cannot guarantee the validity of ML-assisted GWAS associations.

We also provide a theoretical explanation for false positive findings in ML-assisted GWAS. We found that all methods we outlined earlier for ML-assisted GWAS are non-negative weighted sums of GWAS on observed and imputed phenotypes (**Methods** and **Supplementary Note**). This suggests that the estimand for these methods is different from the true parameter of interest, i.e., SNP effect on the observed phenotype, unless true GWAS effects on the observed and imputed phenotypes are identical for all SNPs. This condition is strong and unrealistic. We present several straightforward instances where it is not met (**Supplementary Figure 2**). In conclusion, the validity of conventional ML-assisted GWAS depends on strong conditions that need to be met by all SNPs. However, given the inherent uncertainty in identifying the true data-generating process, it is challenging to empirically validate these strong conditions even after careful selection of imputation models and post-GWAS sensitivity checks. Therefore, we need a valid and powerful inference framework for ML-assisted GWAS that is robust to even mis-specified phenotype imputation from "black-box" ML algorithms.

### POP-GWAS: valid GWAS inference for ML-imputed outcomes

We introduce a novel statistical framework named POP-GWAS for GWAS inference on imputed phenotypes (**Figure 2**). POP-GWAS is a weighted sum of three estimators:

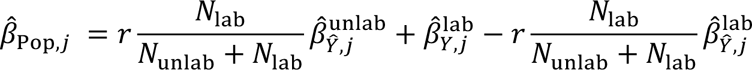

**Figure 2.**
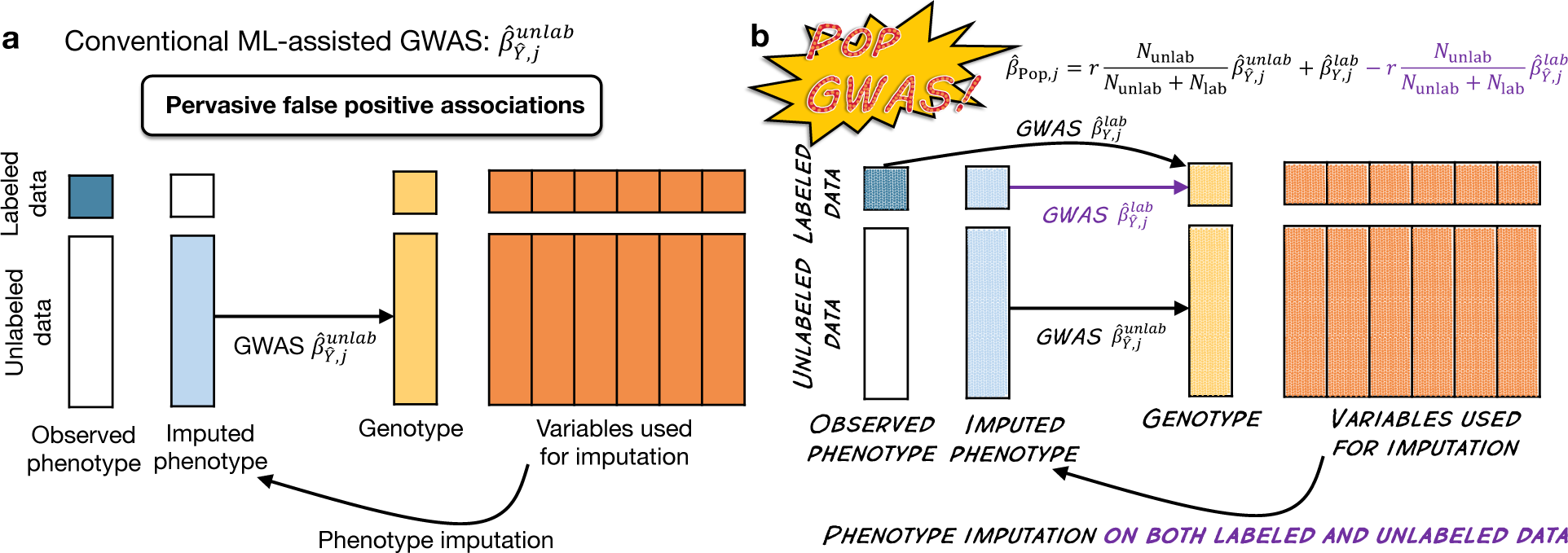
Comparison of POP-GWAS and a conventional design for ML-assisted GWAS. **(a)** A conventional design performs GWAS on the imputed phenotype using unlabeled samples. **(b)** POP-GWAS imputes the phenotype in both labeled and unlabeled samples, and performs three GWAS: GWAS of the observed and imputed phenotype in labeled samples, and GWAS on the imputed phenotype in unlabeled samples. Then, summary statistics of these three GWAS are used to obtain POP-GWAS estimates.

where 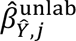 is the estimated effect of *j* -th SNP on imputed phenotype 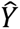 in the unlabeled samples, 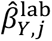 is the SNP effect on observed phenotype in labeled samples, and 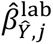 is the SNP effect on imputed phenotype in labeled samples. Following similar notations, we refer to the GWAS that produce these three sets of estimates as 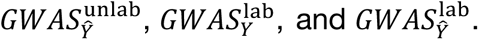 *N*_unlab_and *N*_lab_ are the sample sizes for unlabeled and labeled data, respectively. *r* is the correlation between observed and imputed phenotypes after adjusting for covariates (**Supplementary Note**) which quantifies imputation quality.

The key idea behind POP-GWAS is to use the difference between estimated SNP effects on observed and imputed phenotypes in labeled data (i.e., 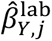 and 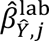) to debias conventional ML-assisted GWAS (i.e., 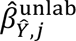) on a SNP-by-SNP basis. Intuitively, if there is no difference between GWAS effects on observed and imputed phenotypes, then we can trust the conventional ML-assisted GWAS. Since the bias is quantified at the SNP level, it ensures the validity of estimation results for each SNP. This is a key difference compared to current approaches that use genome-wide metrics (such as genetic correlation) to verify and correct ML-assisted GWAS which can produce many false positives for individual SNPs. This also proposes a major revision to the current practice of ML-assisted GWAS: we need to impute phenotypes in both labeled and unlabeled samples, and perform GWAS on the imputed phenotypes in labeled samples as part of the routine.

We show several special cases of POP-GWAS to provide more intuition on the approach. When the imputation is perfect (i.e., *r* = 1 and 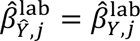), we can ignore the phenotypic heterogeneity and trust the GWAS results on imputed phenotype. In this case, POP-GWAS degenerates to a meta-analysis of 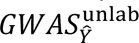 and 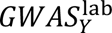 weighted by sample size. If the imputation quality is terrible (i.e., *r* is close to 0), GWAS on the imputed phenotype does not provide any useful information, and POP-GWAS degenerates to 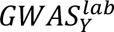 in this scenario.

The POP-GWAS framework has several important features^28^:

1. It always provides unbiased estimates and valid p-values regardless of the imputation algorithm, variables included in imputation, quality of the imputation, and the genetic architecture of the phenotype. POP-GWAS is assumption-free regarding the imputation procedure.
2. It is always more powerful than the GWAS limited to samples with observed phenotypes, i.e., 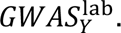 Statistical power further improves with higher imputation quality and a larger sample size ratio between the unlabeled and labeled datasets. Features 1) and 2) ensure that POP-GWAS is a "no-harm" approach compared to 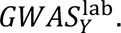
3. It only requires GWAS summary statistics as input. We note that users do not always need to provide the correlation *r* for POP-GWAS since it can be estimated using the intercept of bivariate linkage disequilibrium score regression (LDSC)^29^ between 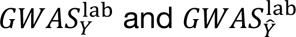^’^ (**Methods**). We also note that misspecification of *r* does not affect the validity of POP-GWAS, but only its estimation efficiency (**Supplementary Note**).
4. It is computationally efficient. It only takes several minutes to produce results for a GWAS with 10 million SNPs.

In **Supplementary Note**, we provide theoretical guarantees for POP-GWAS, including unbiasedness, consistency, asymptotic normality, and statistical efficiency. We also provide solutions for handling binary phenotype, sample relatedness, sample overlap between input GWAS, and selection bias in GWAS samples (**Supplementary Note** and **Supplementary Figure 3-6**).

### Simulations and real data benchmarking for POP-GWAS performance

We performed extensive simulations to validate our theoretical results (**Methods**). We found that conventional ML-assisted GWAS (i.e., 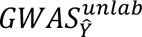) leads to biased estimates and inflated type-I error under heterogeneous genetic effects on observed and imputed phenotypes, while both 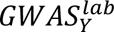 and POP-GWAS remain unbiased and control the type-I error well (**Figure 3a-c**). Moreover, POP-GWAS is consistently more powerful than 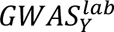 (**Figure 3d**), and its power improves with a greater imputation r² (**Figure 3e**) and a larger sample size ratio between unlabeled and labeled data (**Figure 3f**). We found the same conclusions when applying POP-GWAS to binary phenotypes, GWAS with overlapping samples, and cross-validation (**Supplementary Figures 3-5**). Further simulations were conducted to investigate whether correlated effect sizes of top SNPs on observed and imputed phenotypes ensures the validity of ML-assisted GWAS (**Supplementary Figure 7**). We found no guarantee to the validity of imputed GWAS even with a correlation close to 1. This occurs when most top SNPs have similar effects on observed and imputed phenotypes which drives the high effect correlation, but a small fraction of SNPs only associate with the imputed phenotype. This subset of SNPs identified in the imputed GWAS would become false positive associations. This aligns with our observation in the T2D example, which indicates that cross-SNP metrics, such as high correlation of top SNP effects or genetic correlation based on genome-wide SNPs, cannot guarantee the validity of GWAS on imputed outcomes.

**Figure 3.**
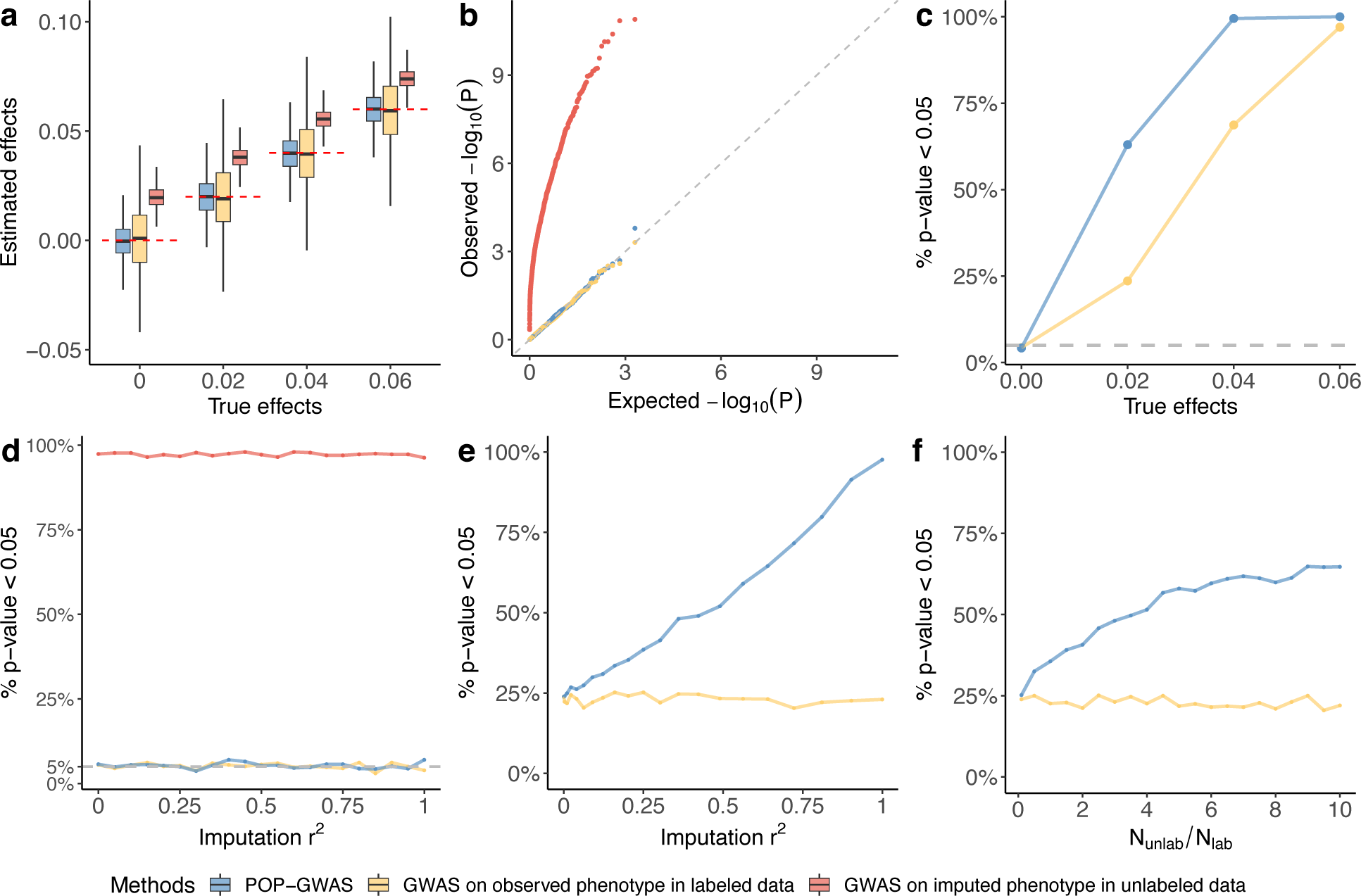
Simulation results. This figure compares POP-GWAS, GWAS of the observed phenotype in labeled data, and GWAS of the imputed phenotype in unlabeled data. **(a)** Point estimation for SNP effects. The red dashed line represents the true effect sizes. **(b)** QQ plot of P-value under the null (i.e., no SNP effects). **(c)** Statistical power under different true effect sizes. **(d)** Type-I error under different imputation r^2^. **(e)** Statistical power under different imputation r^2^. **(f)** Statistical power under different sample size ratio between unlabeled and labeled data.

Next, we applied POP-GWAS to the T2D benchmarking example we have described previously. We performed 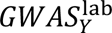 and 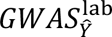 in the 25% labeled sample, and 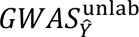 in the 75% unlabeled sample. Phenotype imputation in the labeled sample was implemented through cross-validation to avoid overfitting. We found that all loci identified by POP-GWAS were replicated in the ground truth T2D GWAS (P < 5e-8). None of the erythrocyte variants reported in the conventional GWAS on imputed T2D were significant in POP-GWAS (**Supplementary Figure 8**). Additionally, POP-GWAS identified 116% more loci compared to 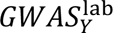 (13 versus 6), suggesting that POP-GWAS leads to an increase in statistical power while ensuring the validity of association findings.

### POP-GWAS is statistically optimal for ML-assisted GWAS

Having demonstrated that POP-GWAS increases power without compromising the validity of GWAS results, we provide evidence for its statistical optimality. We provide a theoretical proof that POP-GWAS is the best linear unbiased estimator (BLUE) given the observed and imputed phenotypes (**Supplementary Note**). This suggests that any attempt to improve POP-GWAS with a linear estimator would result in either estimation bias or lower efficiency. This conclusion leads to a closed-form formula for the upper bound on the effective sample size of a valid ML-assisted GWAS, which is achieved by POP-GWAS.

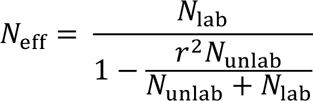

This formula has several implications. First, imputation quality is crucial for effective sample size (**Figure 4a**). With a zero imputation r^2^, the effective sample size for POP-GWAS is equal to the labeled sample size *N*_lab_. With a high imputation r^2^ close to 1, the effective sample size for POP-GWAS is the total sample size *N*_unlab_ + *N*_lab_. Second, this formula shows that given a fixed and imperfect imputation r^2^, there is an upper bound on the effective sample size, even as the unlabeled sample size goes to infinity (**Figure 4b**). This contrasts with the existing formula for effective sample size in the literature (i.e., *N*_lab_ + *r*^2^*N*_unlab_), which suggests it will go to infinity if we keep adding unlabeled samples. The derivation of the existing formula assumes that SNP effects on imputed and observed phenotypes are proportional across all SNPs^19^, which is the same strong (genome-wide) assumption for conventional ML-assisted GWAS to be valid. Third, this allows fast and rigorous statistical power calculation for ML-assisted GWAS. We have implemented the calculator into a Shiny app freely available to the research community (**Data and code availability**).

**Figure 4.**
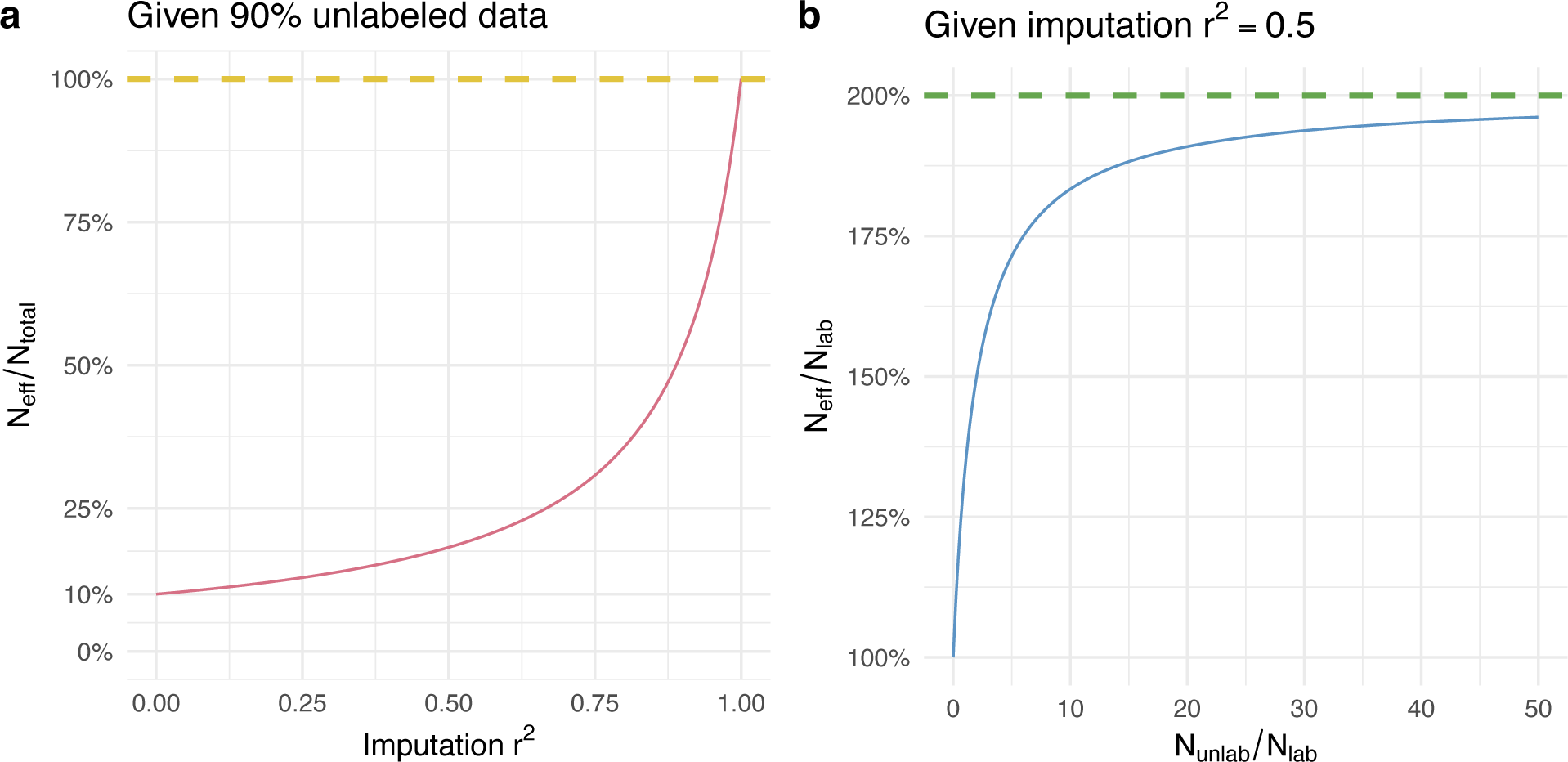
Effective sample size calculation for ML-assisted GWAS. **(a)** For a dataset comprising 90% unlabeled data, the graph illustrates the relationship between the ratio of the effective sample to the total sample size (Y-axis) and the imputation r^2^ of various ML algorithms (X-axis). **(b)** For algorithms with an imputation r^2^ of 0.5, the graph depicts the efficiency gain, represented by the ratio of the effective sample size to the labeled sample size (Y-axis), against the increase in unlabeled sample collection, represented by the ratio of the unlabeled sample size to the labeled sample size. There is an upper bound for the effective sample size given a fixed imputation r^2^.

### POP-GWAS for bone mineral density across 14 skeletal sites

Next, we applied POP-GWAS to conduct the largest GWAS on DXA-derived BMD measures (DXA-BMD) across 14 skeletal sites in UKB. DXA-BMD is the best indicator and primary diagnostic marker for osteoporosis and fracture risk in the clinic^30-32^. It also enables studying the site-specific genetic architecture of BMD which may lead to more accurate assessment of fracture risk in different parts of the skeleton^33-37^. However, DXA-BMD is currently only measured in around 10% of UKB participants. This presents an opportunity for POP-GWAS to uncover new associations. We imputed DXA-BMD in both labeled and unlabeled samples using SoftImpute^21^ (**Methods**). Notable strong predictors include lean body mass, body weight, and heel BMD measured by ultrasound (**Supplementary Table 2**). Cross-validation was implemented for labeled samples to avoid overfitting. Imputation quality, quantified by residual correlation *r* after adjusting for sex, age, their interaction, and top 20 genetic principal components ranged from 0.31 to 0.61 across 14 sites. We conducted 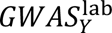 and 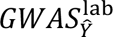 in 40,403 labeled samples, and 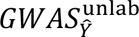 in 367,749 unlabeled samples. These three GWAS were used as input for POP-GWAS.

POP-GWAS achieved a 9.7%-50.7% gain in effective sample size (effective N: 44,267∼60,829) compared to conventional GWAS for measured DXA-BMD. Heritability estimates were 0.18-0.32 across sites (**Supplementary Table 3**). We found significant enrichment of DXA-BMD heritability in conserved DNA regions, super-enhancers, and H3K27ac histone marks (**Supplementary Figure 9**). Across tissue and cell types, heritability enrichment was the strongest in bone and connective tissues (**Supplementary Figure 10**). We found significant enrichment in mesenchymal stem cell-derived chondrocyte cultured cells for all skeletal sites (fold enrichment ranging from 9.5-12.4).

We identified 188 independent loci at p < 1.4e-8 (i.e., 5e-8/3.5, where 3.5 is the effective number of independent traits; **Methods**) across 14 skeletal sites (**Figure 5a**), which is 39% more than the 135 loci identified by conventional GWAS (**Figure 5b** and **Supplementary Figure 11**). Previously, large-scale DXA-BMD GWAS have primarily focused on four skeletal sites, i.e., head, lumbar spine (labeled as L1-L4 in UKB), femur neck, and total body. Therefore, we used existing GWAS based on these 4 sites for replication. We found that 86 of 86 (100%), 54 of 62 (87%), 47 of 52 (90%), and 85 of 90 (94%) of our identified loci reached nominal significance (P<0.05) with consistent effect directions in previous DXA-based GWAS for head, L1-L4, femur neck, and total body BMD, respectively (**Supplementary Figure 12**). POP-GWAS findings showed higher replication rates in independent BMD GWAS from similar sites. However, cross-site replication rates were also high, ranging from 53% to 92% (**Supplementary Figure 12**). We performed meta-analysis to combine POP-GWAS with site-matched DXA-BMD associations from independent studies, reaching total sample sizes of 81,622, 79,071, 79,940, and 117,015 for head, L1-L4, femur neck, and total body DXA-BMD, respectively. We found 115 more loci reaching genome-wide significance in meta-analysis. In total, we identified 89 novel loci that had not been previously implicated in BMD studies (**Supplementary Figure 12** and **Supplementary Table 4**).

**Figure 5.**
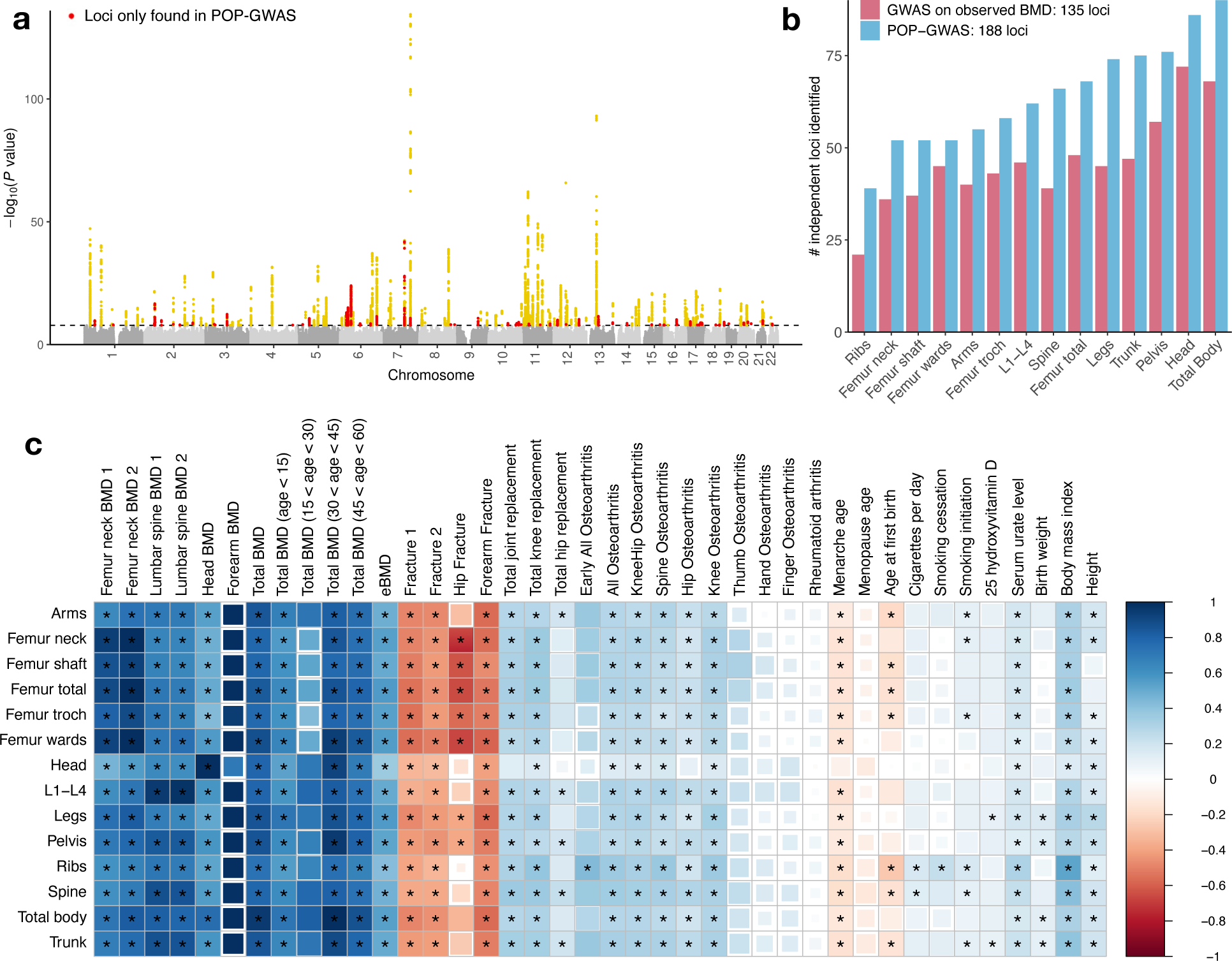
POP-GWAS for DXA-BMD across 14 skeletal sites. **(a)** Manhattan plot for DXA-BMD POP-GWAS. Red dots represent the loci only found in POP-GWAS, but not in conventional GWAS of observed DXA-BMD. The P-value displayed for each SNP is the smallest P-value across 14 sites. **(b)** The number of genome-wide significant loci identified for each skeletal site based on conventional GWAS and POP-GWAS. **(c)** Genetic correlation between BMD GWAS and 40 complex traits. The color represents the point estimates. The size of the square represents the P-value. Asterisks highlight significant genetic correlations after Bonferroni correction (P < 3.6e-4). Femoral neck BMD 1 and 2 represent two different GWAS on the same phenotype (**Supplementary Table 8**). We use similar labels for lumbar spine and fracture studies.

Gene set enrichment analysis (**Methods**; **Supplementary Table 5**) identified significant enrichment of BMD associations in skeletal growth (e.g., clock-controlled autophagy in bone metabolism, osteoblast differentiation and related diseases, chondrocyte differentiation, and cartilage development) and signaling pathways involved in bone biology (e.g., *WNT*, Hedgehog, *ALK*, TGFβ, *PITX2* signaling pathways). We also found evidence for co-localization of novel DXA-BMD GWAS loci and osteoclast cis-eQTL^38,39^. 18 genes at 14 distinct loci reached a co-localization posterior probability of 50% (**Supplementary Table 6** and **Supplementary Figures 14-15**). Several identified genes have shown functional evidence in cell and mouse models. For instance, *COL4A2* enhances osteogenic differentiation of periodontal ligament stem cells by negatively regulating the *Wnt*/β-catenin pathway within the extracellular matrix^40^. Mice deficient in *Wwox* exhibit osteopenia, a condition marked by reduced bone density^41^. Using Mendelian randomization, we identified 12 genes whose expression in osteoclast may causally link to BMD (false discovery rate [FDR] < 0.05; **Supplementary Table 7**). In particular, we found that upregulation of *WWOX* may causally increase BMD (FDR-adjusted P = 7e-3).

Our analyses also revealed skeletal site-specific genetic architecture for DXA-BMD. Head BMD exhibited the highest heritability (h² = 0.32, se = 0.03; **Supplementary Table 3**), yet only shows modest genetic correlations (ranging from 0.5 to 0.67) with BMD at other sites (**Supplementary Figure 16**). In comparison, associations identified at other skeletal sites showed substantial pleiotropy, with genetic correlations spanning from 0.7 to 0.97. We also estimated genetic correlations of DXA-BMD with 40 published GWAS, including 12 previous BMD studies, 4 fracture studies, osteoarthritis at 12 skeletal sites, and 12 other complex traits (**Figure 5c** and **Supplementary Table 8**). Genetic correlations of independent BMD GWAS from the same skeletal site were stronger than cross-site BMD genetic correlations. For instance, existing femur neck DXA-BMD GWAS showed strong correlations with our association results from femur sites (e.g., cor = 0.94 with femur neck POP-GWAS), and lumbar spine DXA-BMD is strongly correlated with L1-L4 POP-GWAS (cor = 0.98). Site-specific genetic sharing was also observed beyond BMD phenotypes^31,32^. For example, hip fracture risk showed particularly strong genetic correlation with DXA-BMD in femur sites^36^.We also found significant correlations between DXA-BMD and osteoarthritis^33^ in weight-bearing joints (knee, hip, and spine) but not in non-weight-bearing joints (hand, finger, and thumb) osteoarthritis. Notably, estimated BMD (eBMD) using ultrasound in the heel only exhibited moderate correlations with DXA-BMD (ranging from 0.36 to 0.69), which is consistent with the known limitations of eBMD measurement^42^. In fact, we found a more substantial genetic correlation of hip fracture with femur neck DXA-BMD (cor = -0.71)than with heel eBMD (cor = -0.51) (**Supplementary Figure 17**). This suggests that POP-GWAS obtained clinically more relevant genetic associations than heel eBMD while using heel eBMD as a key predictor for phenotype imputation. Furthermore, we observed differences in BMD genetic association across age groups. The BMD in younger individuals displayed weaker genetic correlations with our GWAS conducted in middle-aged groups^43^.

Given the weaker genetic correlation between head BMD and other sites, we investigated genomic loci showing site-specific association with head BMD alone. We found 3 loci with strong head-specific effects (not reaching nominal significance P < 0.05 at any other skeletal sites; **Supplementary Table 4**). One example is the *LGR5* locus on chromosome 12 (lead SNP rs12308154; p=1.5e-9; **Figure 6a** and **b**). *LGR5* regulates the *WNT* signaling pathway and is crucial for bone formation, remodeling, and homeostasis^44^. *LGR5*-expressing cells are primarily located in the mesenchyme adjacent to craniofacial epithelial structures that are undergoing folding, such as the nasopharyngeal duct, lingual groove, and vomeronasal organ. During early craniofacial development, *LGR5* mRNA was observed in the mesenchyme surrounding the mandibular cleft and the lateral aspects of the tongue, indicating its involvement in key stages of embryonic development^45^. Additionally, *LGR5* is critical during embryogenesis, as mice lacking *Lgr5* incurred 100% neonatal mortality accompanied by several craniofacial distortions, such as ankyloglossia and gastrointestinal dilation, highlighting its importance in the proper formation of craniofacial features^46^.

**Figure 6.**
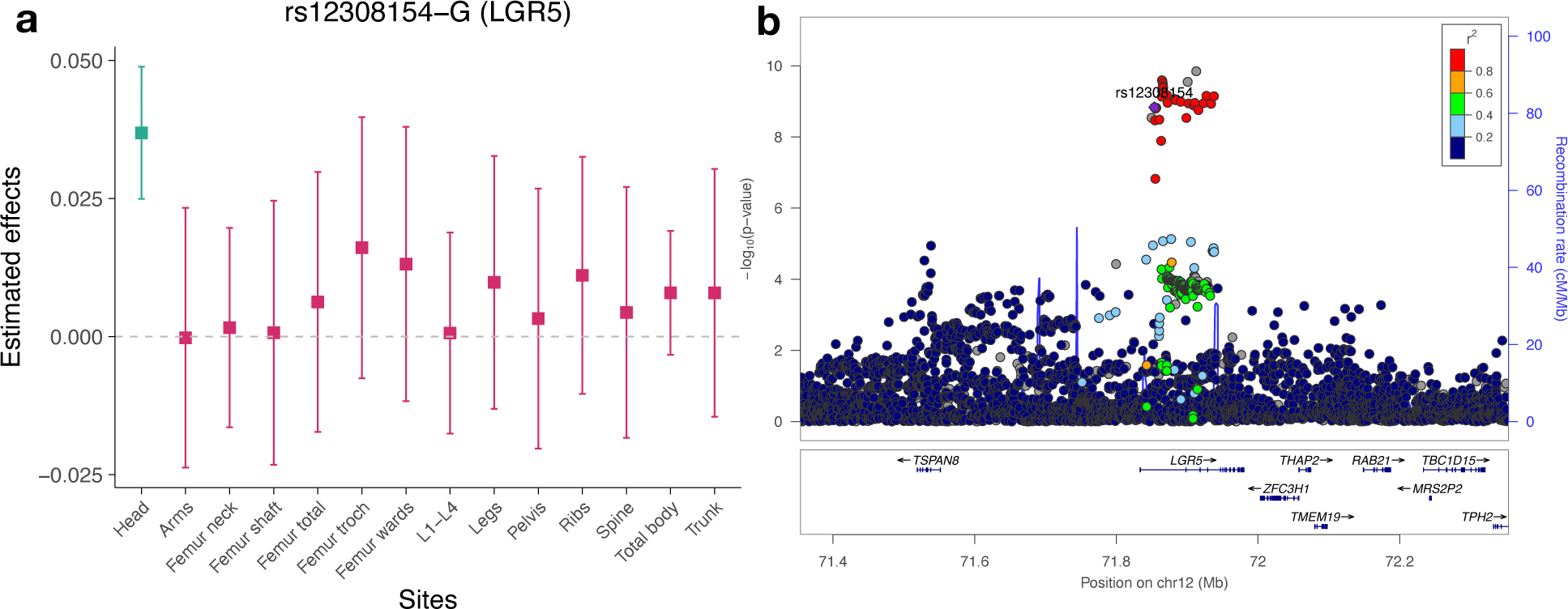
LGR5 as a head-specific GWAS signal. **(a)** Effects of rs12308154-G (*LGR5*) on DXA-BMD across 14 sites in UKB. **(b)** Associations at the *LGR5* locus from head DXA-BMD meta-analysis.

## Discussion

In recent years, GWAS of ML-imputed phenotypes has quickly emerged as a crucial study design for complex trait genetics, gaining popularity due to its ability to leverage large biobank samples and identify new associations. However, existing approaches do not sufficiently account for the distinction between observed and imputed outcomes and have high risks of identifying false positive associations. To address this issue, we introduced POP-GWAS, a principled statistical framework for valid and powerful inference in ML-assisted GWAS based on summary statistics alone. POP-GWAS uses imputed phenotypes in labeled samples to ensure valid inference and then leverages phenotype imputation in unlabeled samples to boost statistical power. We have demonstrated its statistical superiority through extensive theoretical and empirical analyses.

We highlight several major advances in our study that will reshape future ML-assisted GWAS applications. First, our study cautions against the false positive associations in conventional ML-assisted GWAS. This is highlighted by a shocking 81% replication failure rate we observed in the GWAS of imputed T2D. This discrepancy stems partly from the use of HbA1c for imputation, where associated genetic variations are influenced by both glycemic and erythrocytic mechanisms but the glycemic processes are more relevant for T2D risk. This revelation of false positive associations has broad implications, suggesting a pervasive issue in current ML-assisted GWAS especially when the causal relationships between predictor variables and the primary outcome remain unclear. We also demonstrated that current approaches that use genome-wide metrics to guard against false positive associations cannot guarantee the validity of ML-assisted GWAS. Our theoretical analyses reinforce these empirical findings by establishing conditions under which such false associations are expected to occur. As demonstrated in our T2D example, these conditions are not merely hypothetical but are frequently encountered in real-world studies. We further examined several crucial yet long-neglected issues in ML-assisted GWAS practice. We showed that current studies may overestimate the power increase of ML-assisted GWAS if GWAS covariates are used for phenotype imputation. Non-random missingness for the phenotype is also likely to affect the validity of ML-assisted GWAS, and we have developed an approach to correct for such biases.

Second, several key features make POP-GWAS a superior choice for ML-based GWAS compared to existing methods. It is an "assumption-free" and “no-harm” method, having no requirements about the predictor variables, algorithms, or quality of phenotype imputation, while still improving statistical power and ensuring validity of associations. This flexibility gives researchers important practical convenience, and also embraces a large body of machine learning literature on statistical inference based on predicted outcomes^47,48^. Additionally, POP-GWAS is both user-friendly and computationally efficient. Compared to joint models for primary and surrogate phenotypes^49^, our approach only requires three sets of GWAS summary statistics as input and completes GWAS analysis for millions of SNPs within minutes. We have also made necessary extensions to account for binary phenotypes, sample relatedness, and overlapping samples between summary statistics datasets, making POP-GWAS a versatile tool suitable for broad applications. POP-GWAS also sets a pivotal course for the future developments of ML-assisted GWAS. Clearly, ML is going to continue revolutionizing big data analytics and offer new ways to uncover genetic insights. However, these opportunities come with significant risks due to the "black-box" nature of modern ML algorithms. We demonstrate that the adoption of ML in genetic research should be paralleled by the development of accompanying statistical methods. These methods are essential for ensuring the reliability and interpretability of findings obtained using ML-assisted approaches.

Third, we provide rigorous and closed-form power calculation for ML-assisted GWAS based on the accuracy of phenotype imputation and sample sizes of labeled and unlabeled data. Given budget constraints, researchers often need to choose between measuring phenotypes of higher quality in a smaller sample and measuring less expensive but imprecise variables in a larger population. Our method offers a strategic framework for adding ML-assisted phenotype imputation into the equation, enabling scientists to design efficient GWAS that yield more accurate and generalizable results in genetic research. This is a crucial advance that can facilitate informed decisions regarding resource allocation and cost-benefit analyses, ensuring optimal use of funding and time in future studies.

In addition to these conceptual and methodological advances, we also employed POP-GWAS to conduct the largest GWAS of DXA-BMD across 14 skeletal sites. POP-GWAS identified 39% more loci compared to conventional approaches and provided crucial insights into the skeletal site-specific genetic architecture of BMD. In total, we identified 303 genome-wide significant loci for DXA-BMD, including 89 novel loci not previously implicated in GWAS meta-analyses, marking a significant advancement in understanding BMD’s genetic landscape. The strong genetic correlations between fracture and DXA-BMD at similar skeletal sites underscore the importance of using site-matched BMD in fracture risk assessment. Our genetic correlation results also highlight BMD as a risk factor for osteoarthritis specifically in weight-bearing joints. Several novel GWAS loci identified in our study demonstrate colocalization with osteoclast cis-eQTL, suggesting their potential regulatory impact. These evidence, coupled with functional data for genes such as *COL4A2* and *WWOX*, may offer novel targets for therapeutic intervention. In addition, our analyses also revealed individual genomic loci exhibiting skeletal site-specific effects on BMD. One example is the convincing head-specific BMD association at the leucine-rich repeat-containing, G protein-coupled receptor gene *LGR5*, which is further supported by existing evidence of *Lgr5*-deleted mice exhibiting a range of craniofacial abnormalities^46^. These findings offer a deeper understanding of the genomic underpinning of BMD and hold significant implications for the study of osteoporosis, fracture, osteoarthritis, and related skeletal conditions, potentially guiding new approaches in diagnosis and treatment. We note that our DXA-BMD GWAS was mainly restricted to participants of European ancestry. Therefore, future studies are needed to investigate how these results can be generalized to other populations.

In conclusion, we have uncovered major limitations of current ML-assisted GWAS, introduced a methodological solution that may reshape future study design, demonstrated its superiority over existing methods, and employed a largest GWAS to date of DXA-BMD. We believe that POP-GWAS offers an innovative solution to the challenges in ML-assisted human genetics research and has broad applications in future complex trait genetic studies.

## Methods

### POP-GWAS

As described in the main text, POP-GWAS estimator is

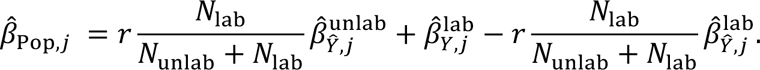

Its corresponding standard error is

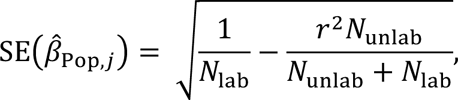

Therefore, the effective sample size can be calculated as

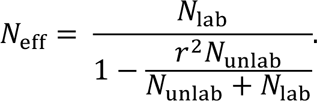

The derivation assumes that the beta is the effect of the standardized allele on the standardized phenotype. However, we will use SNP allele frequencies to convert POP-GWAS estimates to per-allele effect sizes in the phenotypic standard deviation unit. Our implemented algorithm can be found in **Supplementary Note.**

POP-GWAS ensures valid inference with its test statistic following an asymptotically normal distribution:

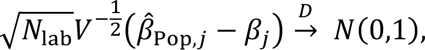

where 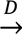 denotes converge in distribution, and *V* is defined as

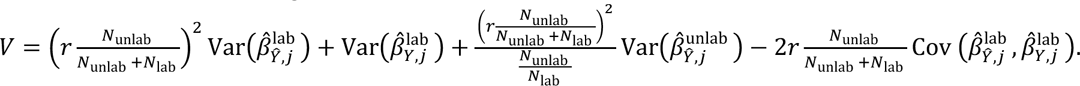

POP-GWAS ensures powerful inference with its improved efficiency over GWAS on observed phenotype. The relative efficiency between 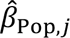 and 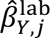 is

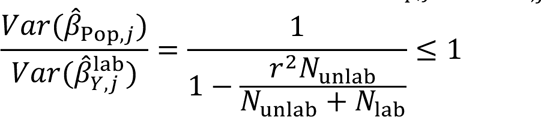

POP-GWAS is the statistical optimal estimator because it has the smallest variance among the class of linear unbiased estimator denoted as

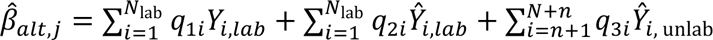

where *q*_1*i*_, *q*_2*i*_, and *q*_3*i*_ are weights that ensure 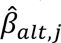 is unbiased.

With POP-GWAS, we present a new protocol for ML-assisted GWAS that consists of three steps:

1. Perform phenotypic imputation on both the labeled and unlabeled data, using any user-preferred imputation variables and algorithms. Use cross-validation in labeled data to avoid overfitting.
2. Conduct 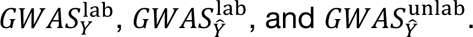
3. Feed summary statistics from these three GWAS into POP-GWAS, and obtain a valid and powerful ML-assisted GWAS.

### T2D imputation and GWAS in UKB

We randomly split the 408,325 individuals with European ancestry in UKB into two subsets with 25% and 75% of all samples. We treated the 25% subsample as labeled data. We masked the phenotype in the 75% subsample and treated it as unlabeled data. The ground truth T2D phenotype is defined based on data field 41202 (Diagnoses - main ICD10: E11 Non-insulin-dependent diabetes mellitus). To select the variables used for imputation, we calculate the phenome-wide correlation (after adjusting for GWAS covariates) with T2D in the labeled data using 463 other phenotypes which are measured in more than 200,000 in the UKB (**Supplementary Table 9**). Data fields that include T2D diagnosis (e.g. self-reported T2D) were excluded from phenotype imputation. We selected the top 50 variables with highest correlations and used the residuals after adjusting for GWAS covariates as variables for imputation. We used the labeled samples to train the SoftImpute algorithm. Then, we applied this model to impute T2D liability in the unlabeled samples. We used 10-fold cross-validation for T2D imputation in the labeled samples.

We applied pre-GWAS quality control (QC) by keeping autosomal biallelic SNPs with MAF > 0.01, missing call rate ≤ 0.01, Hardy-Weinberg equilibrium test p-value ≥ 1.0e-6. We further excluded participants with discrepancies between genetically inferred (data field 22001) and self-reported sex (data field 31), as well as those who had withdrawn or were recommended for exclusion by UKB (data field 22010). We conducted the GWAS of ground truth T2D using Regenie^23^ in all 408,325 samples. We further conducted 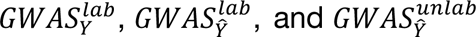 in the 25% labeled and 75% unlabeled data. We adjusted for sex, age, their interaction, and top 20 principal components in each GWAS. Then, we applied POP-GWAS using these three GWAS as input. To count the number of independent genome-wide significant associations, we performed LD clumping with PLINK. We calculated LD from 10,000 randomly selected independent individuals of European ancestry in UKB, and set clumping parameters p1 = p2 =1.5e-8, r2 = 0.01, and clump-kb = 5000. We further collapsed the resulting SNPs to within 100kb of each other.

### Comparison of ML-assisted GWAS methods

We compared several ML-assisted GWAS methods. The detailed derivation and technical discussion can be found in **Supplementary Note**. We use the same notations 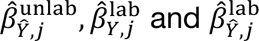 to denote three GWAS summary statistics as described in the main texts. The estimator in existing methods can be written as the non-negative weighted sum of these three GWAS estimators:

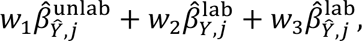

where *w*_1_, *w*_2_, *w*_3_ are all non-negative weights. All existing methods have valid confidence intervals if and only if 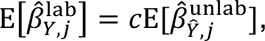 where 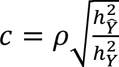 for MTAG (*ρ* is the genetic orrelation, 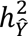 and 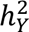 are the heritability for imputed and observed phenotypes), and *c* = 1 for other methods.

### Simulations

We compared POP-GWAS with other approaches using simulations. Each simulation was repeated 1,000 times. We simulated a quantitative phenotype with the following model:

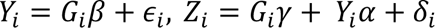

where *Y*_*i*_ is the phenotype, *G*_*i*_ is the SNP, and *Z*_*i*_ is the variable used for imputation. We first generated the SNP *G*_*i*_from Binomial(2, 0.25), where 0.25 is the minor allele frequency. We set *β* to be 0, 0.02, 0.04, and 0.06 and simulated *ϵ*_*i*_ independently from a normal distribution with mean zero and variance such that *Var*(*Y*_*i*_) = 1. We simulated values of *γ* to ensure that *G*_*i*_ explains 0.015% of the variance of *Z*_*i*_. We simulated *δ*_*i*_ from *N*(0, 0.2), and set *α* to let *Var*(*Z*_*i*_) = 1. We generated 120,000 samples and then spilt the sample into labeled and unlabeled dataset. Sample sizes for labeled dataset and unlabeled dataset were set to be 20,000 and 100,000, respectively. We used half of the labeled data to fit a linear regression between *Y*_*i*_ and *Z*_*i*_ and then imputed *Y*_*i*_ in the remaining half labeled and unlabeled samples. We calculated 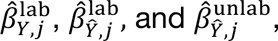 and used them as input for POP-GWAS. We changed the imputation *r*^2^ by altering the proportion of *Z*_*i*_’s variance explained by *Y*_*i*_, ranging from 0% to 95% with increments of 5%. We also varied the sample size ratio between unlabeled and labeled data, setting the sample size at 10,000 for labeled data and 1,000 to 5,000 for unlabeled data with increments of 1,000, and also 10,000 to 100,000 with increments of 10,000. Details for other simulations can be found in **Supplementary Note**.

### POP-GWAS application to DXA-BMD

We followed the same procedures on data QC, phenotype imputation and GWAS outlined in the “T2D imputation and GWAS in UKB” section to analyze 14 DXA-BMD phenotypes (i.e., arms: data field 23225, femur neck (left): data field 23299, femur shaft (left): data field 23290, femur total (left): data field 23291, femur troch (left): data field 23295, femur wards (left): data field 23297, head: data field 23226, L1-L4: data field 23203, legs: data field 23231, pelvis: data field 23232, ribs: data field 23233, spine: data field 23234, trunk: data field 23241, total: data field 23236). We employed the METAL software^50^ for meta-analysis, focusing on four sites (i.e., L1-L4, head, total body, and femur neck) with previously published large GWAS. We performed sample overlap correction implemented in METAL for head and total body BMD due to the overlap of a small subset of UKB individuals in our analysis and published GWAS. Novel BMD loci were defined as genome-wide significant POP-GWAS loci that are not in LD with six previous BMD GWAS^30,31,35,42,43^ (tag-r2 0.01 and tag-kb 100).

We used LDSC^29,51^ to compute heritability and genetic correlation. We used the ‘coloc’ package^52^ in R with its default settings for co-localization analysis (window size = 2MB). We considered a posterior probability greater than 50% for hypothesis H4 (indicating association with both trait 1 and trait 2, with one shared variant) as evidence of colocalization. We conducted heritability enrichment analysis using stratified LDSC^53^ with the baselineLD V2.2^54^ genomic annotations and GenoSkyline-Plus^55^ tissue and cell-type specific annotations. We performed gene set enrichment analysis in FUMA^56^ v1.6.0 with default MAGMA^57^ settings. We performed Mendelian randomization using the SMR approach^58^ with osteoclast cis-eQTL summary statistics. Significant genes were found based on FDR-adjusted SMR P-value < 0.05 and p_HEIDI > 0.05. To determine the effective number of independent traits, we used the formula 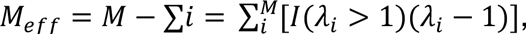 where *M* is the total number of traits and *λ*_*i*_ is the eigenvalue of the genetic correlation matrix across 14 skeletal sites for BMD^59^.

## Data Availability

All data produced are available online at https://qlulab.org/data.html

## Data and code availability

GWAS summary statistics for skeletal site-specific DXA-BMD are available at https://qlu-lab.org/data.html. POP-GWAS software and the power calculator app for ML-assisted GWAS are publicly available at https://github.com/qlu-lab/POP-TOOLS.

## Acknowledgments

The authors gratefully acknowledge research support from National Institutes of Health (NIH) grant U01 HG012039, and support from the University of Wisconsin-Madison Office of the Chancellor and the Vice Chancellor for Research and Graduate Education with funding from the Wisconsin Alumni Research Foundation (WARF). We also acknowledge use of the facilities of the Center for Demography of Health and Aging at the University of Wisconsin-Madison, funded by NIA Center Grant P30 AG017266. We thank members of the Social Genomics Working Group at University of Wisconsin for helpful comments. This research has been conducted using the UK Biobank Resource under Application 42148. The font choice in Figure2B is inspired by pop art.

## Author contribution

J.M. conceived the study and developed the statistical framework.

J.M., Y.W., and Z.S. performed data analysis.

Y.W. implemented the software.

X.M. developed the method to account for selection bias.

T.L. advised on result interpretation.

J.Z. and Q.L. advised on statistical issues.

Q.L. advised on genetic issues.

J.M. and Q.L. wrote the manuscript.

All authors contributed to manuscript editing and approved the manuscript.

