## Supplementary Figure for "Valid inference for machine learning-assisted GWAS"

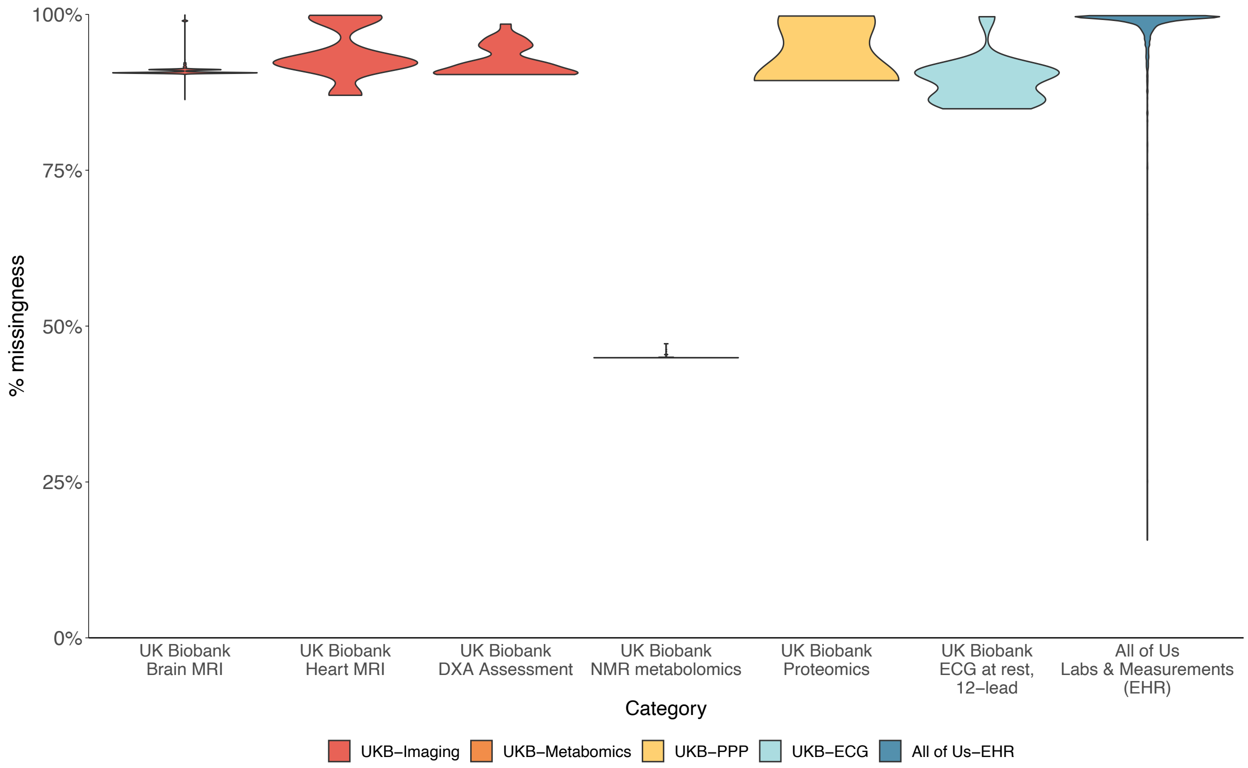


**Supplementary Figure 1. Date missing rate across variable types in UKB and All of Us.**

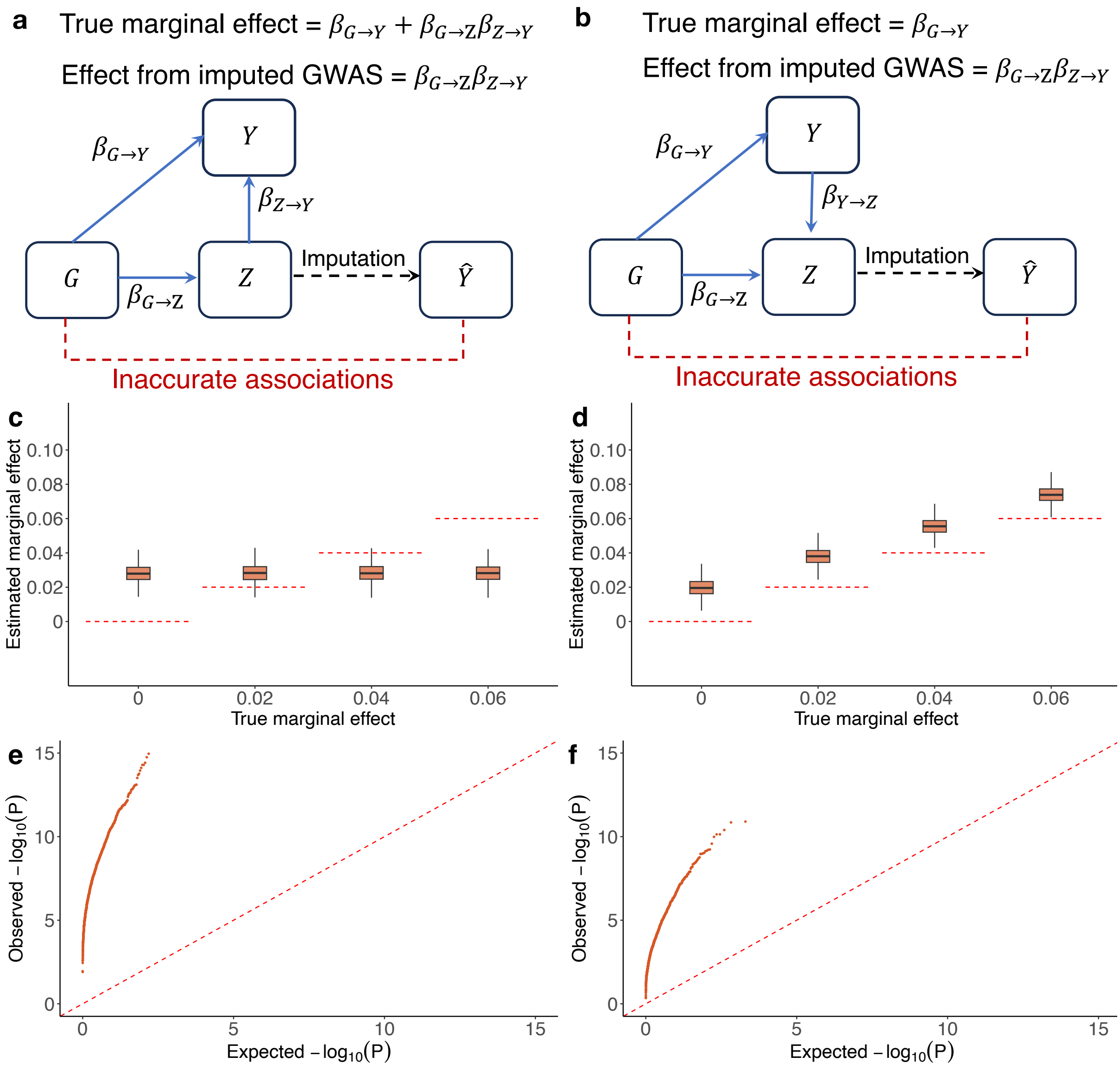


**Supplementary Figure 2. Instances where the equal SNP effect condition is violated. (a)-(b)** Data generating process for SNP $G$, phenotype $Y$, and variables used for imputation $Z$. **(c)-(d)** Point estimation for SNP effects. The red dashed line represents the true effect sizes. **(e)-(f)** QQ plot of P-value under the null (i.e., no SNP effects).

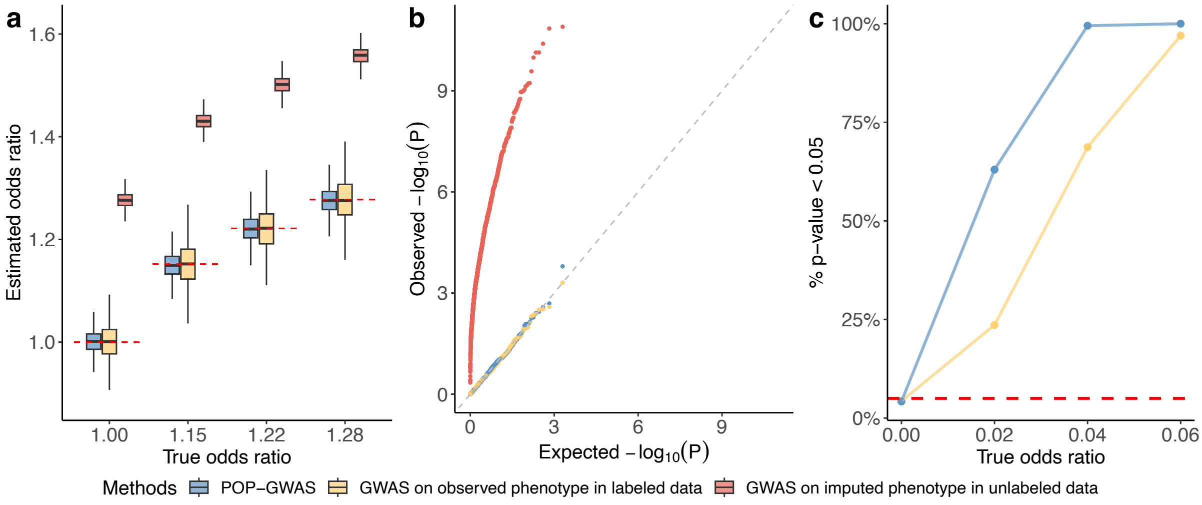


**Supplementary Figure 3. Simulations for binary phenotype. (a)** Point estimation for SNP effects. The red dashed line represents the true effect sizes. **(b)** QQ plot of P-value under the null (i.e., no SNP effects). **(c)** Statistical power under different true effect sizes.

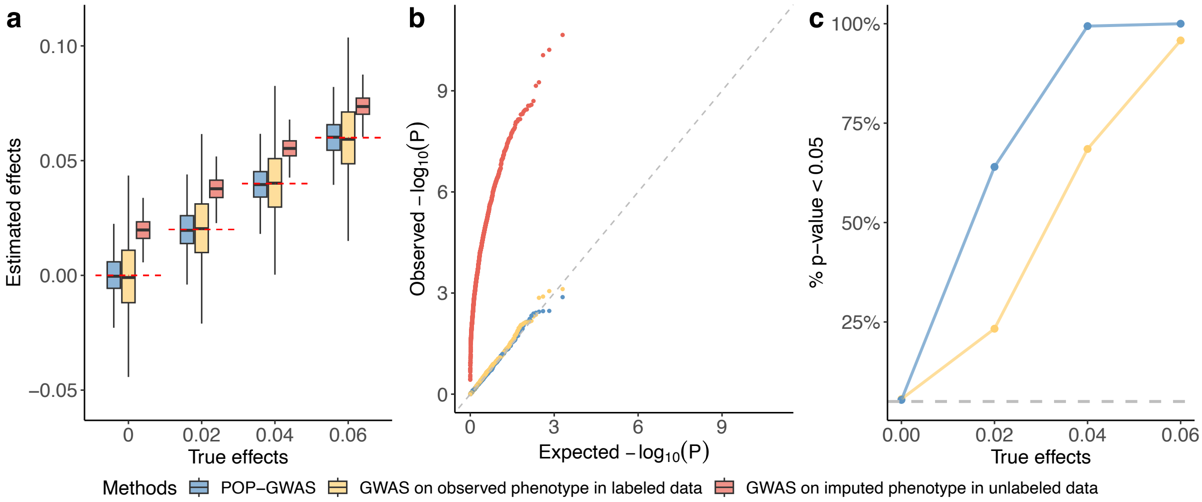


**Supplementary Figure 4. Simulations using GWAS with overlapping samples. (a)** Point estimation for SNP effects. The red dashed line represents the true effect sizes. **(b)** QQ plot of P-value under the null (i.e., no SNP effects). **(c)** Statistical power under different true effect sizes.

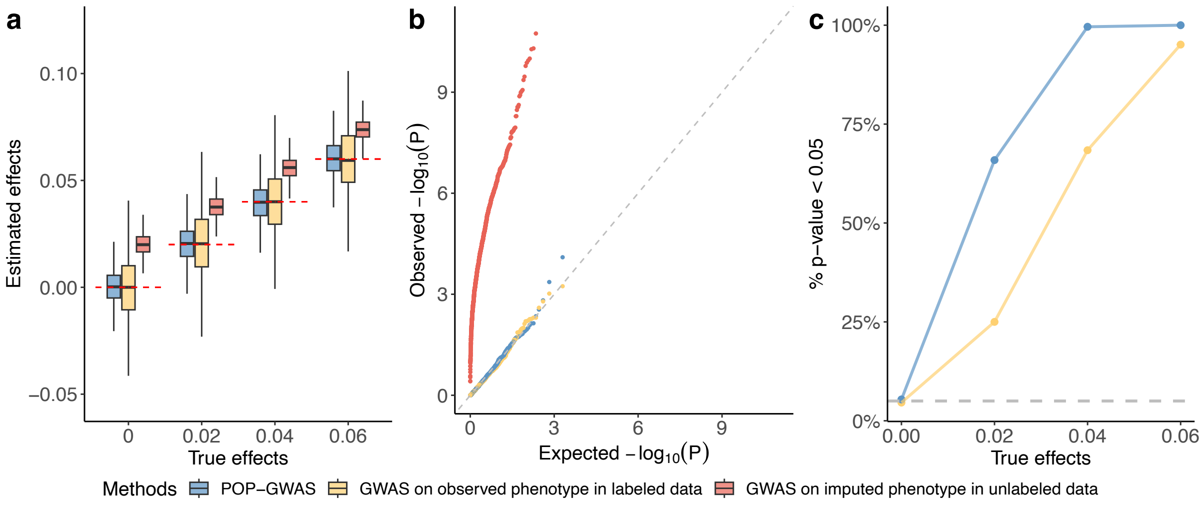


**Supplementary Figure 5. Simulations using cross-validation for phenotype imputation in labeled data. (a)** Point estimation for SNP effects. The red dashed line represents the true effect sizes. **(b)** QQ plot of P-value under the null (i.e., no SNP effects). **(c)** Statistical power under different true effect sizes.

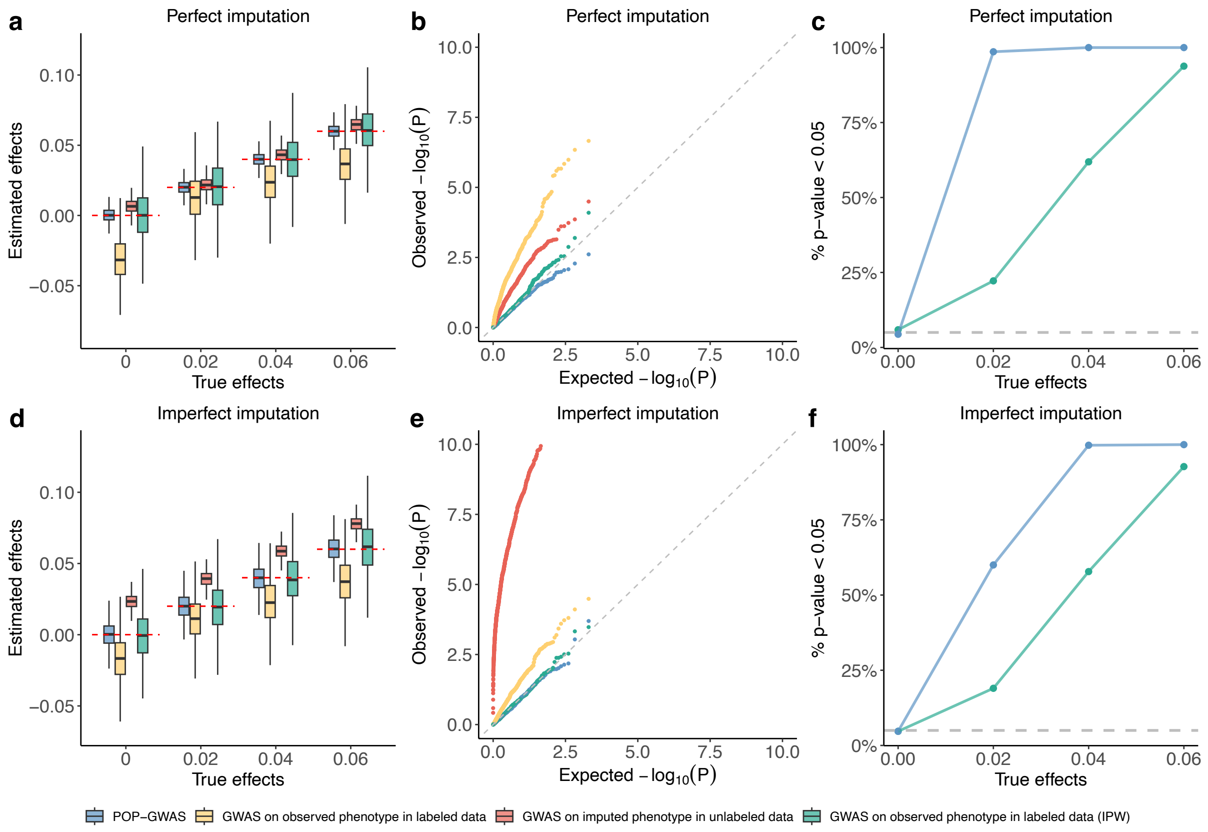


**Supplementary Figure 6. Simulations for selection bias. (a)** Point estimation for SNP effects. The red dashed line represents the true effect sizes. **(b)** QQ plot of P-value under the null (i.e., no SNP effects). **(c)** Statistical power under different true effect sizes. **(d)-(f)** is the same as **(a)-(c)** but with imperfect imputation.

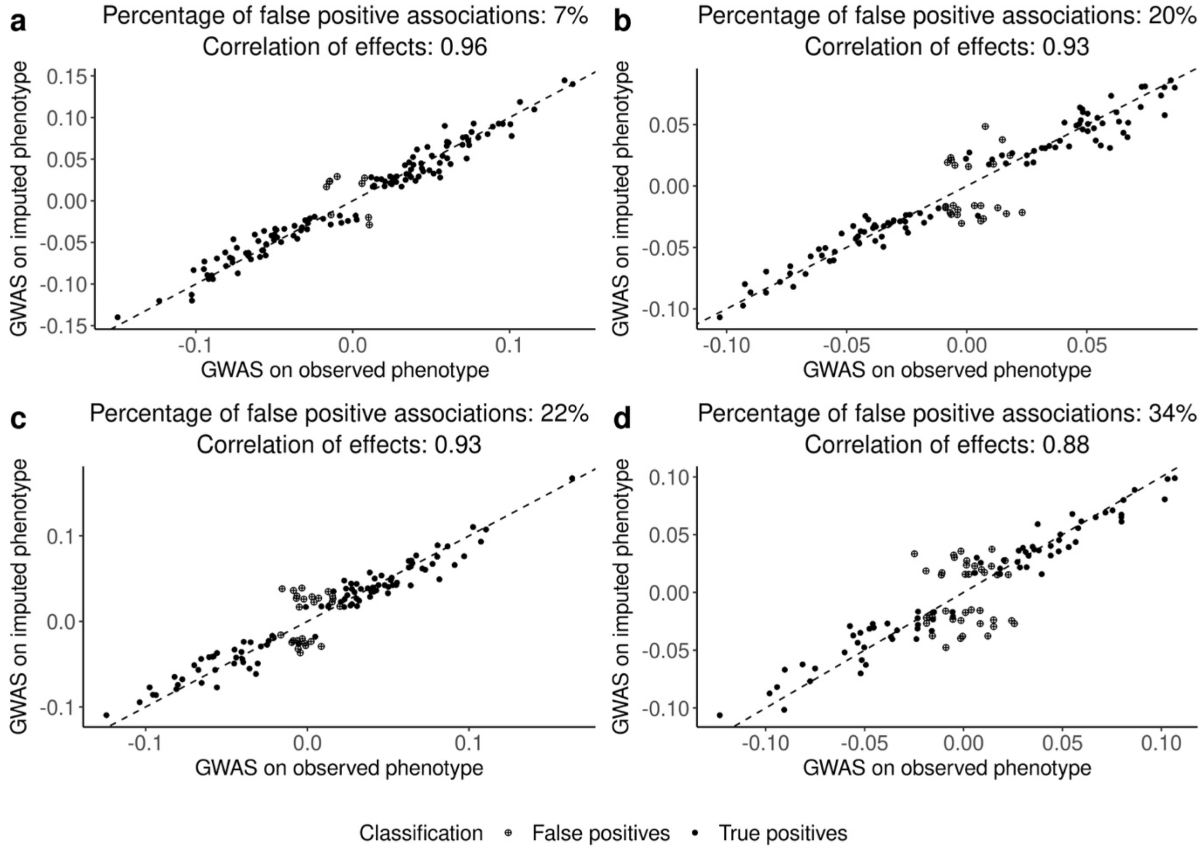


**Supplementary Figure 7. Correlated effect sizes of top SNPs on observed and imputed phenotypes cannot ensure the validity of ML-assisted GWAS.** Each point is a SNP with P<5e-8 in the GWAS on imputed phenotype.

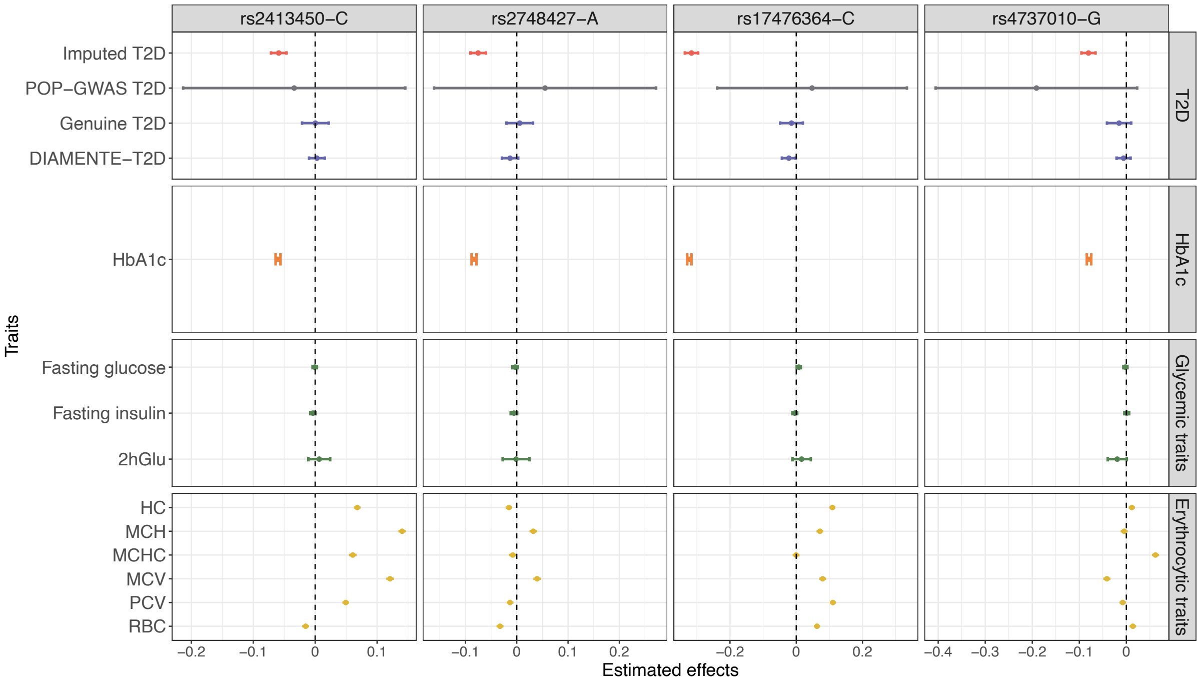


**Supplementary Figure 8. Estimated effects of four erythrocytic SNPs on T2D, HbA1c, glycemic traits, and erythrocytic traits with POP-GWAS.** The vertical dashed line at 0 serves as a reference for no effect. Error bars show the 95% confidence intervals. Abbreviations: 2hGlu (2-h glucose after an oral glucose challenge), HC (haemoglobin concentration), MCH (mean corpuscular haemoglobin), MCHC (mean corpuscular haemoglobin concentration), MCV (mean corpuscular volume), PCV (haematocrit percentage), RBC (red blood cell count).

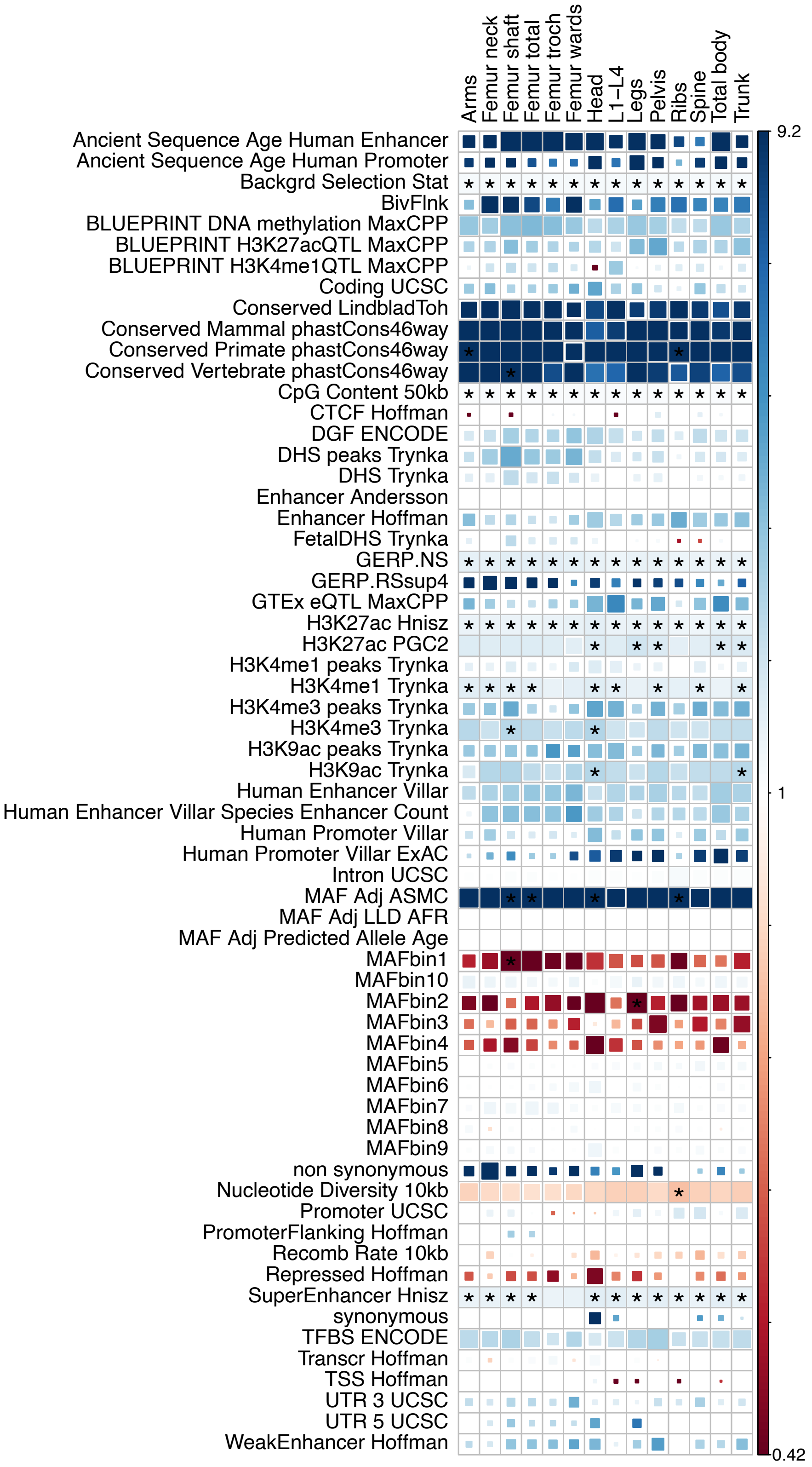


**Supplementary Figure 9. Heritability enrichment using baseline LD v2.2.** The color represents the point estimates. The size of the square represents the P-value. Asterisks highlight significant enrichment after Bonferroni correction (P < 0.05/63/3.5 = 2.2e-4).

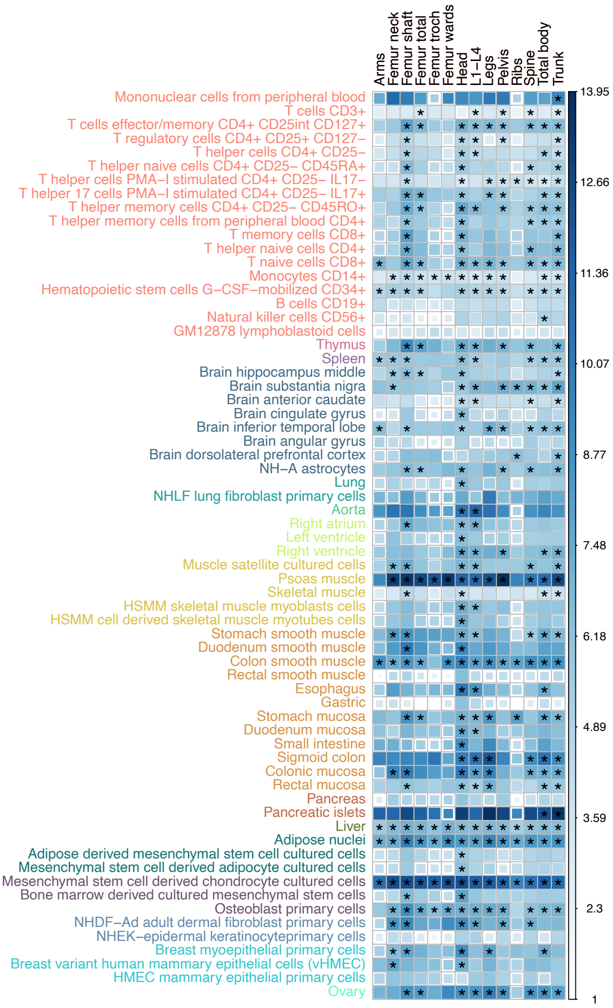


**Supplementary Figure 10. Heritability enrichment using GenoSkyline-Plus.** The color represents the point estimates. The size of the square represents the P-value. Asterisks highlight significant enrichment after Bonferroni correction (P < 0.05/66/3.5 = 2.2e-4).

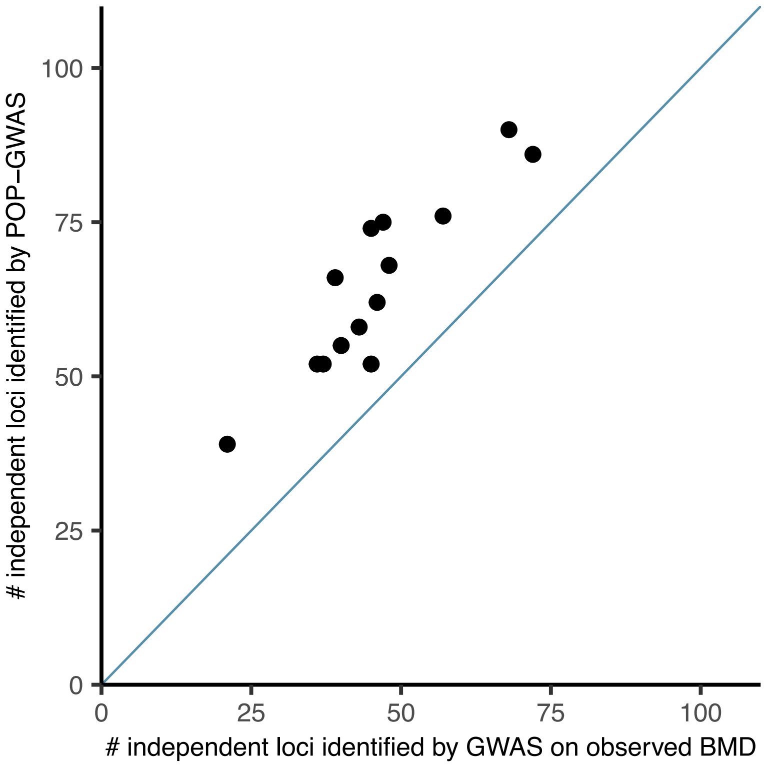


**Supplementary Figure 11. Number of loci identified by GWAS on observed BMD and POP-GWAS.** The blue line represents y = x.

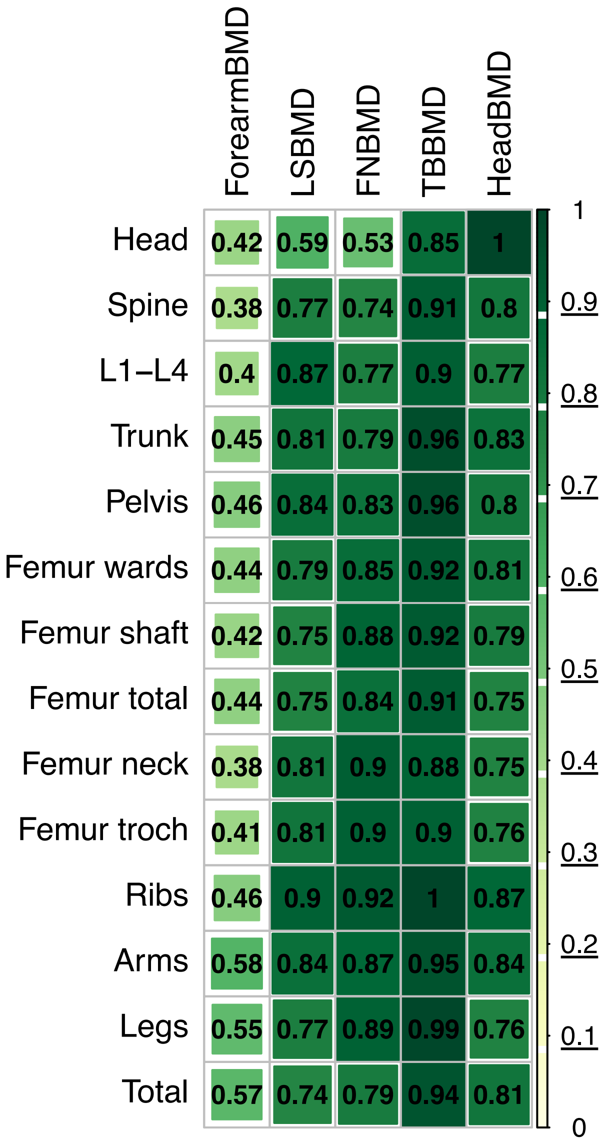


**Supplementary Figure 12. Replication of POP-GWAS associations.** The number in the figure represents the replication rate. The X-axis is the existing GWAS for BMD and Y-axis is our POP-GWAS. Darker color and larger square size represent higher replication rate. Abbreviations: LSBMD: lumbar spine BMD, FNBMD: femur neck BMD, TBBMD: Total Body BMD.

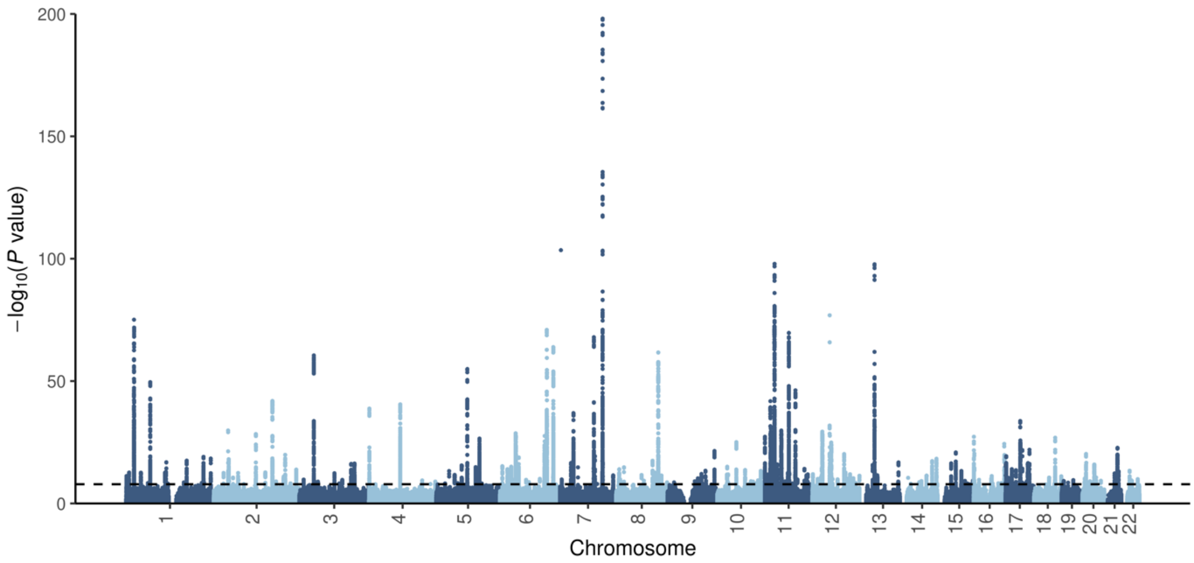


**Supplementary Figure 13. Manhattan plot for 4 meta-analyzed + 10 POP-GWAS GWAS for DXA-BMD.** The p-value for each SNP is the smallest P-value across 14 skeletal sites. The P-value cutoff is 1.4e-8.

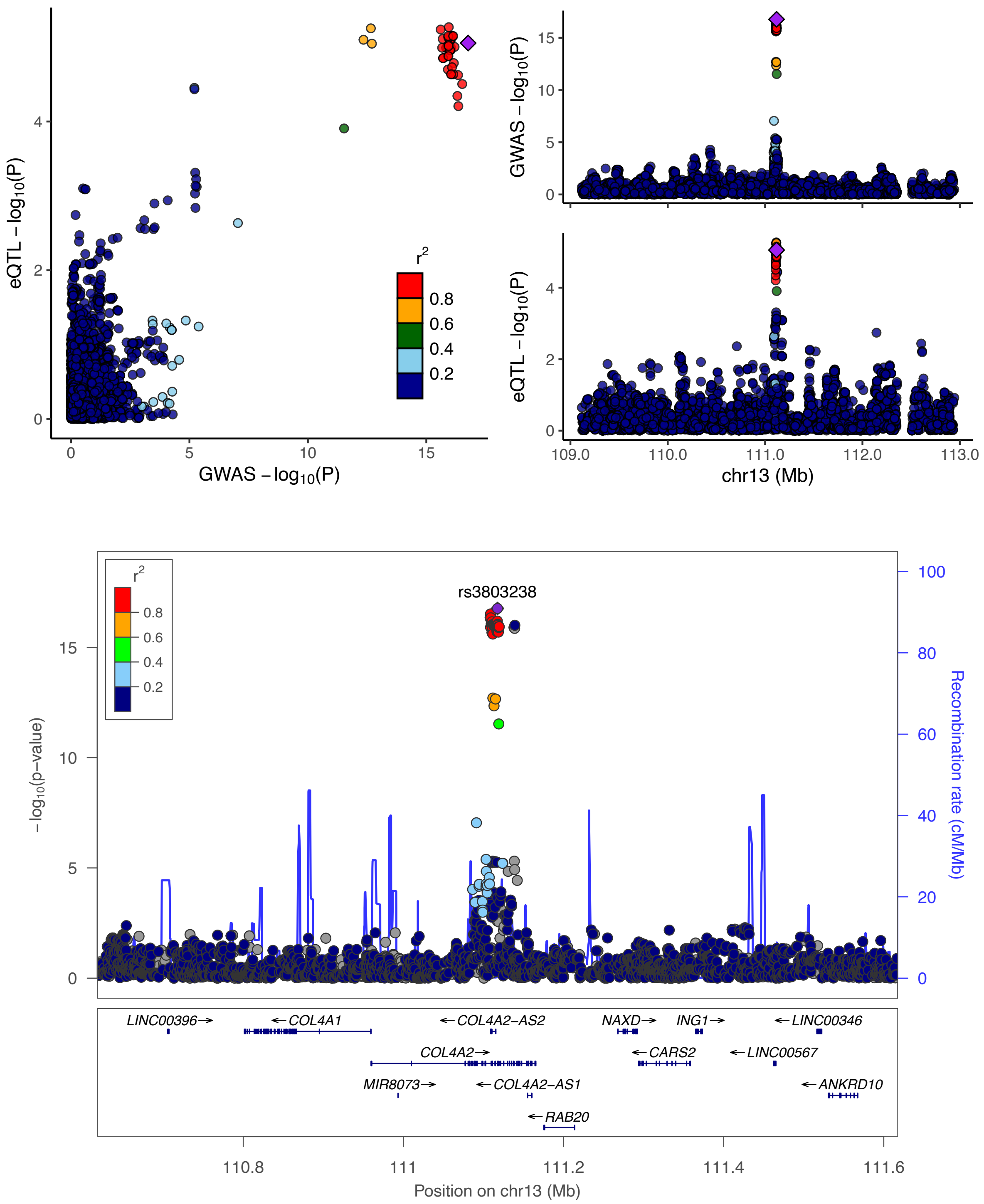


**Supplementary Figure 14. Colocalization between rs3803238 and osteoclast cis-eQTL for *COL4A2*.**

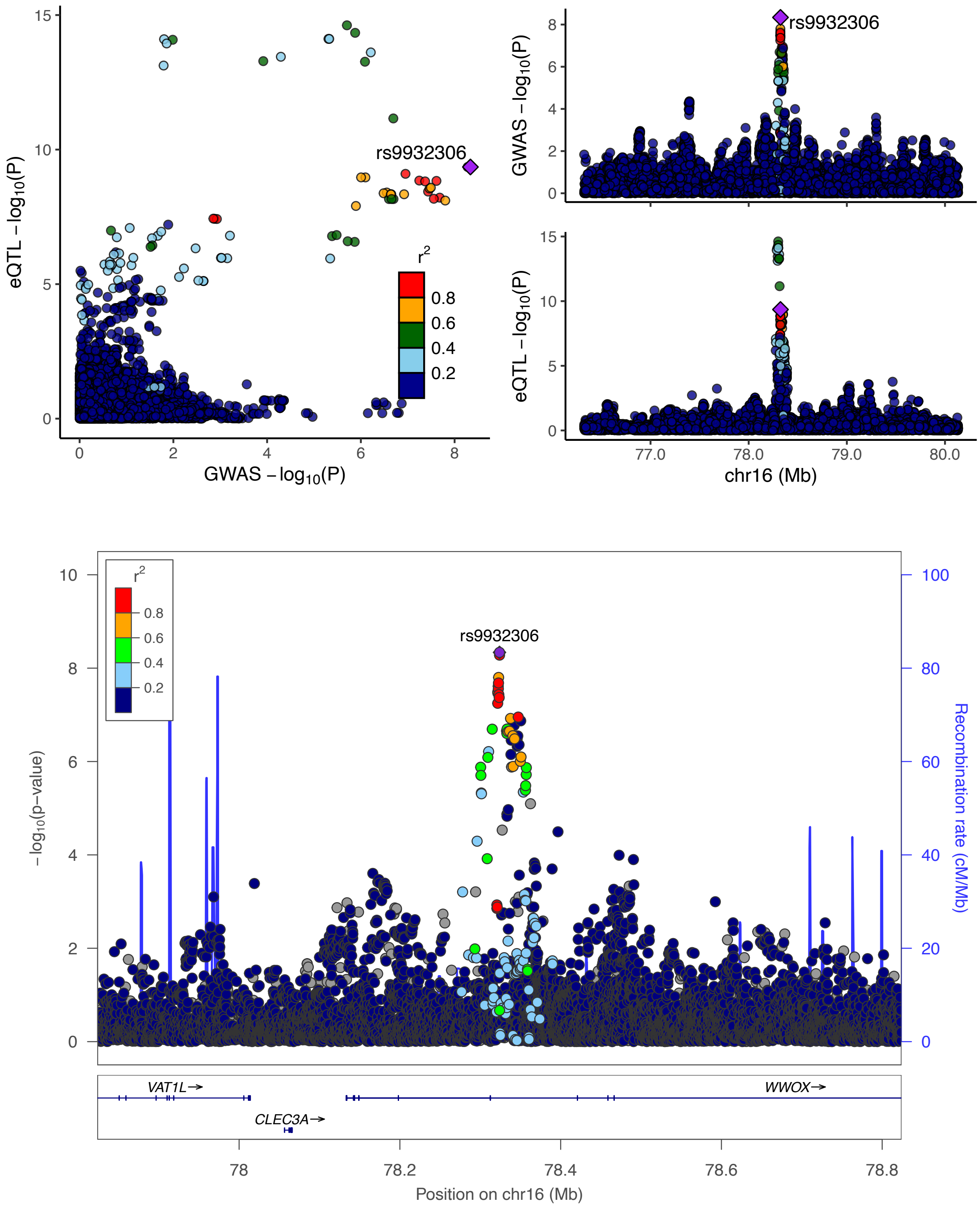


**Supplementary Figure 15. Colocalization between rs9932306 and osteoclast cis-eQTL for *WWOX*.**

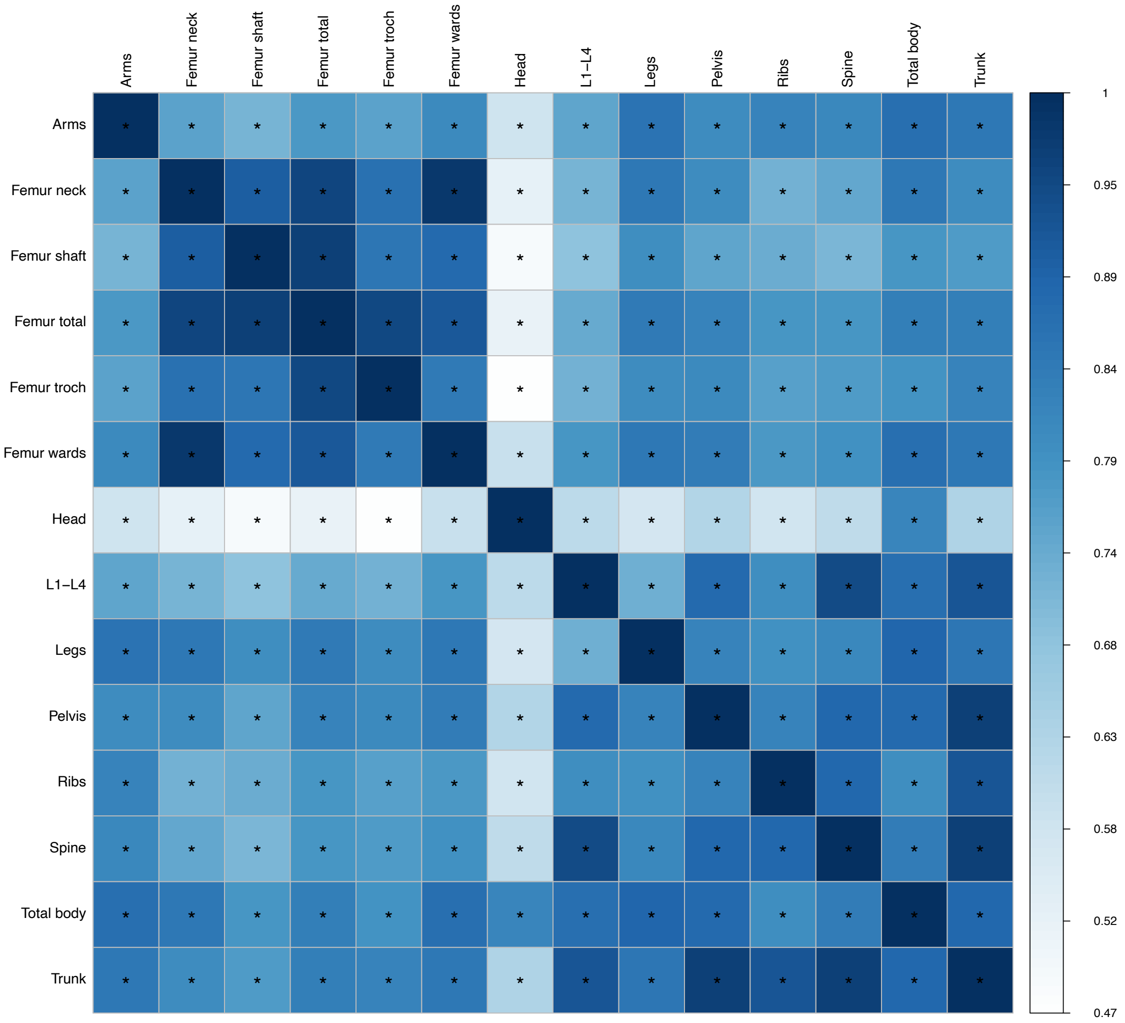


**Supplementary Figure 16. Genetic correlation of DXA-BMD across 14 skeletal sites.** The color represents the point estimates. The size of the square represents the P-value. Asterisks highlight significant genetic correlations after Bonferroni correction (P < 0.05/3.5/14 = 1.0e-3).

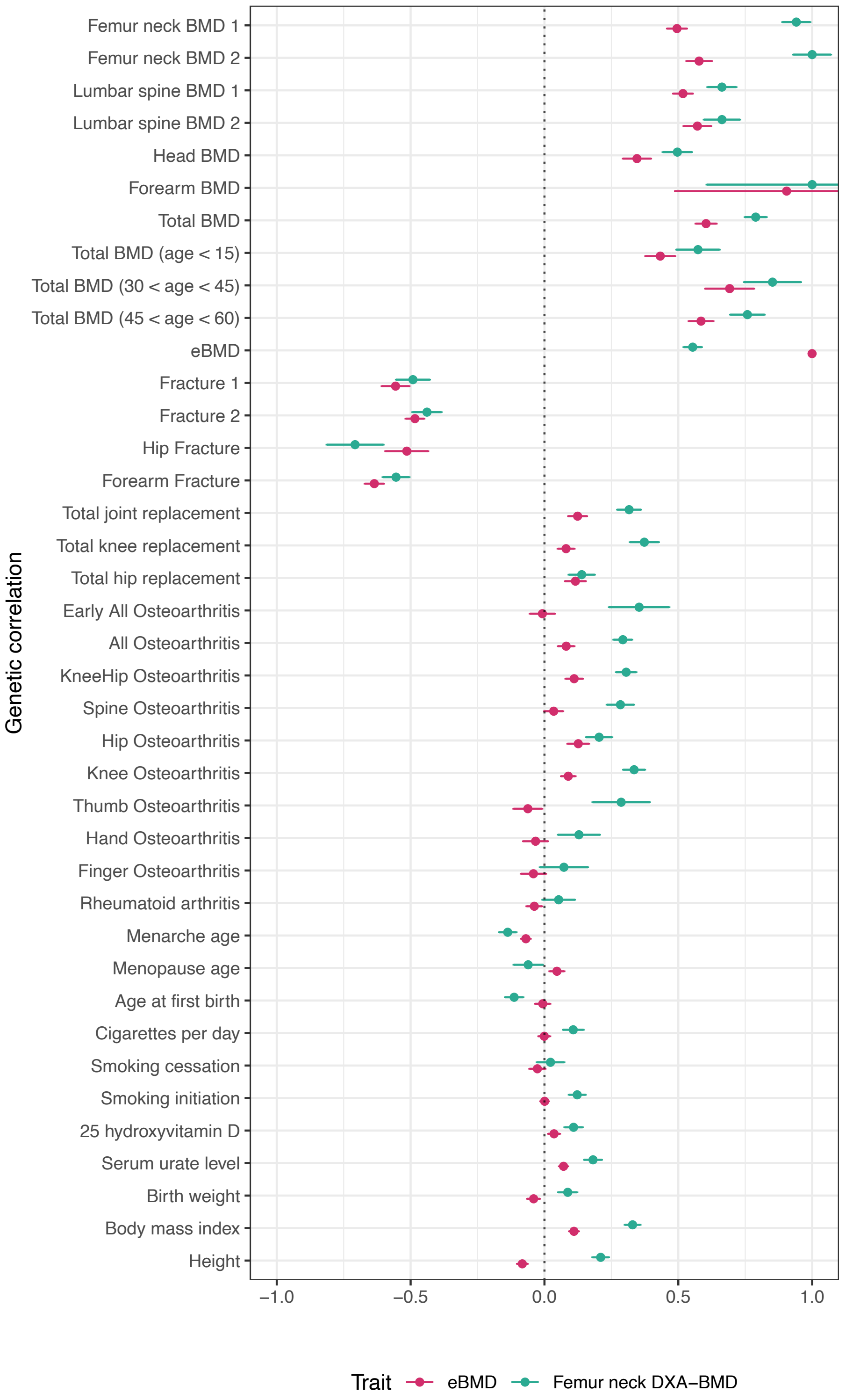


**Supplementary Figure 17. Comparison of genetic correlation results between femur neck DXA-BMD and heel eBMD.** The error bars denote standard errors.
