## Supplementary Note for "Valid inference for machine learning-assisted GWAS"

### Supplementary Note to
### "Valid inference for machine learning-assisted GWAS"

Jiacheng Miao[1], Yixuan Wu[1], Zhongxuan Sun[1], Xinran Miao[2], Tianyuan Lu[3,4], Jiwei Zhao[1,2], and Qiongshi Lu[1,2,5]

[1]Department of Biostatistics and Medical Informatics, University of Wisconsin-Madison
[2]Department of Statistics, University of Wisconsin-Madison
[3]Lady Davis Institute for Medical Research, Jewish General Hospital
[4]Department of Statistical Sciences, University of Toronto
[5]Center for Demography of Health and Aging, University of Wisconsin–Madison

### 1 Notations and Models

#### 1.1 Estimand and model

We first introduce the notations, estimand, and model setup for GWAS on imputed phenotype [machine learning (ML)-assisted GWAS]. We assume that $Y_i$ is the gold standard phenotype of interest.

We consider the estimand for GWAS to be the per-standardized-allele effect of SNP $G_{ij}$ on the standardized $Y_i$ with mean of 0 and variance of 1:

$$\beta_j = \frac{\mathrm{Cov}(G_{ij}, Y_i)}{\mathrm{Var}(G_{ij})} = \mathrm{Cor}(G_{ij}, Y_i)$$

The interpretation is the increase in standard deviation units of phenotype per standardized allele. Consider the simple linear regression model for GWAS:

$$Y_i = G_{ij}\beta_j + \epsilon_i,$$
$$\hat{Y}_i = G_{ij}\beta_j^* + \delta_i.$$

where

- $Y_i$ is the (standardized) observed phenotype with a mean of 0 for the $i$-th individual
- $G_{ij}$ is the $j$-th (standardized) SNP
- $\hat{Y}_i = \widehat{f}(\mathbf{Z}_i)$ is the (standardized) imputed phenotype based on (centerd) predictor variable $\mathbf{Z}_i$.
- the machine learning algorithm $\widehat{f} : Z_i \to \hat{Y}_i$ is obtained using training data $\boldsymbol{D}_{\text{train}} = (\boldsymbol{Z}_{\text{train}}, \boldsymbol{Y}_{\text{train}})$
- $\beta_j$ and $\beta_j^*$ denotes that the effect of SNP $G_{ij}$ on observed and imputed phenotypes, respectively.
- $\epsilon_i$ and $\delta_i$ are the residuals, where $\epsilon_i \perp G_{ij}, \mathbb{E}[\epsilon_i] = 0, \mathrm{Var}(\epsilon_i) = \sigma^2, \delta_i \perp G_{ij}, \mathbb{E}[\delta_i] = 0, \mathrm{Var}(\delta_i) = {\sigma^*}^2$
- The imputation quality is quantified by the correlation between observed and imputed phenotypes $r := \mathrm{Cor}(Y_i, \hat{Y}_i) \in [-1, 1]$.

We use the superscript " $*$ " to denote quantities related to the GWAS of the imputed phenotype $\hat{Y}$.

The above models can be rewritten in matrix form for notation purposes

$$\boldsymbol{Y} = \boldsymbol{G}_j\beta_j + \boldsymbol{\epsilon}$$
$$\hat{\boldsymbol{Y}} = \boldsymbol{G}_j\beta_j^* + \boldsymbol{\delta},$$

where $\boldsymbol{Y}$ and $\hat{\boldsymbol{Y}}$ are two $(N_{\text{lab}} + N_{\text{unlab}}) \times 1$ (standardized) vectors, $\boldsymbol{G}_j$ is a $(N_{\text{lab}} + N_{\text{unlab}}) \times 1$ vector. $\hat{\boldsymbol{Y}}$ is imputed based on a $(N_{\text{lab}} + N_{\text{unlab}}) \times t$ matrix $\boldsymbol{Z}$.

#### 1.2 Two data and three GWAS summary statistics

The data in the whole sample are denoted as

$$\boldsymbol{D}_{\text{whole}} = (\boldsymbol{G}_{\text{whole}_j}, \boldsymbol{Y}_{\text{whole}}, \hat{\boldsymbol{Y}}_{\text{whole}}, \boldsymbol{Z}_{\text{whole}}) = [(\boldsymbol{G}_{\text{lab}_j}, \boldsymbol{Y}_{\text{lab}}, \hat{\boldsymbol{Y}}_{\text{lab}}, \boldsymbol{Z}_{\text{lab}})^{\mathrm{T}}, (\boldsymbol{G}_{\text{unlab}_j}, \boldsymbol{Y}_{\text{unlab}}, \hat{\boldsymbol{Y}}_{\text{unlab}}, \boldsymbol{Z}_{\text{unlab}})^{\mathrm{T}}]^{\mathrm{T}}$$

with number of rows being $N_{\text{lab}} + N_{\text{unlab}}$. Note that $\boldsymbol{Y}_{\text{unlab}}$ is unobserved, but we keep it for completeness of notation. We also assume $\boldsymbol{D}_{\text{whole}} \perp \boldsymbol{D}_{\text{train}}$ to avoid overfitting.

**Two data:**

- Labeled data with gold standard phenotype observed at sample size $N_{\text{lab}}$:

$$(\boldsymbol{G}_{\text{lab}_j}, \boldsymbol{Y}_{\text{lab}}, \hat{\boldsymbol{Y}}_{\text{lab}}, \boldsymbol{Z}_{\text{lab}}),$$

  where $(\boldsymbol{G}_{\text{lab}_j}, \boldsymbol{Y}_{\text{lab}}, \hat{\boldsymbol{Y}}_{\text{lab}}, \boldsymbol{Z}_{\text{lab}})$ are the $N_{\text{lab}} \times 1$ vector for $j$-th, $N_{\text{lab}} \times 1$ gold standard phenotype vector, $N_{\text{lab}} \times 1$ imputed phenotype vector, and $N_{\text{lab}} \times K$ the variables used for the imputation matrix in the labeled data, respectively.

- Unlabeled data with sample size $N_{\text{unlab}}$ with missing gold-standard phenotype:

$$(\boldsymbol{G}_{\text{unlab}_j}, \boldsymbol{Y}_{\text{unlab}}, \hat{\boldsymbol{Y}}_{\text{unlab}}, \boldsymbol{Z}_{\text{unlab}})$$

**Three GWAS summary statistics:**

- GWAS on observed phenotype in labeled data:

$$\begin{aligned}
\hat{\beta}^{\text{lab}}_{Y,j} &= (\boldsymbol{G}^{\text{T}}_{\text{lab}_j}\boldsymbol{G}_{\text{lab}_j})^{-1}\boldsymbol{G}^{\text{T}}_{\text{lab}_j}\boldsymbol{Y}_{\text{lab}},\\
\text{SE}(\hat{\beta}^{\text{lab}}_{Y,j}) &= \sqrt{\hat{\sigma}^2_{\text{lab}}(\boldsymbol{G}^{\text{T}}_{\text{lab}_j}\boldsymbol{G}_{\text{lab}_j})^{-1}}\\
z^{\text{lab}}_{Y,j} &= \hat{\beta}^{\text{lab}}_{Y,j}/\,\text{SE}(\hat{\beta}^{\text{lab}}_{Y,j})
\end{aligned}$$

- GWAS on imputed phenotype in labeled data:

$$\begin{aligned}
\hat{\beta}^{\text{lab}}_{\hat{Y},j} &= (\boldsymbol{G}^{\text{T}}_{\text{lab}_j}\boldsymbol{G}_{\text{lab}_j})^{-1}\boldsymbol{G}^{\text{T}}_{\text{lab}_j}\hat{\boldsymbol{Y}}_{\text{lab}},\\
\text{SE}(\hat{\beta}^{\text{lab}}_{\hat{Y},j}) &= \sqrt{\hat{\sigma}^{*2}_{\text{lab}}(\boldsymbol{G}^{\text{T}}_{\text{lab}_j}\boldsymbol{G}_{\text{lab}_j})^{-1}}\\
z^{\text{lab}}_{\hat{Y},j} &= \hat{\beta}^{\text{lab}}_{\hat{Y},j}/\,\text{SE}(\hat{\beta}^{\text{lab}}_{\hat{Y},j})
\end{aligned}$$

- GWAS on imputed phenotype in unlabeled data:

$$\begin{aligned}
\hat{\beta}^{\text{unlab}}_{\hat{Y},j} &= (\boldsymbol{G}^{\text{T}}_{\text{unlab}_j}\boldsymbol{G}_{\text{unlab}_j})^{-1}\boldsymbol{G}^{\text{T}}_{\text{unlab}_j}\hat{\boldsymbol{Y}}_{\text{unlab}},\\
\text{SE}(\hat{\beta}^{\text{unlab}}_{\hat{Y},j}) &= \sqrt{\hat{\sigma}^{*2}_{\text{unlab}}(\boldsymbol{G}^{\text{T}}_{\text{unlab}_j}\boldsymbol{G}_{\text{unlab}_j})^{-1}}\\
z^{\text{unlab}}_{\hat{Y},j} &= \hat{\beta}^{\text{unlab}}_{\hat{Y},j}/\,\text{SE}(\hat{\beta}^{\text{unlab}}_{\hat{Y},j})
\end{aligned}$$

### 2 Comparing commonly-used methods for ML-assisted GWAS

Here, we first present the general form of the current estimator for ML-assisted GWAS. We then evaluate the unbiasedness and confidence validity (Rubin, 1996) of these estimators for several commonly used methods. Confidence validity is defined by Robin and its violation leads to invalid statistical inference (type I error inflation).

**Definition 1** (Confidence validity)**.** Confidence validity, meaning that for interval estimation, actual interval coverage > nominal interval coverage, and for hypothesis testing, actual rejection rate < nominal rejection rate.

We then have the following lemma for confidence validity.

**Lemma 1** (Robin 1996)**.** *Let $Q$ be an estimand of interest, $\hat{Q}$ be an estimator of $Q$ with sampling variance estimated by $\hat{U}$. Assume that the asymptotic sampling distribution of $\hat{Q}$ is normal, so that valid confidence intervals can be obtained from $\hat{Q}$ and $\hat{U}$. In this case, confidence validity requires*

$$\begin{aligned}
\mathbb{E}[\hat{Q}] &= Q\\
\mathbb{E}[\hat{U}] &\geq \text{Var}(\hat{Q})
\end{aligned}$$

Next, we derive the estimators for our methods for ML-assisted GWAS.

#### 2.1 Estimator for commonly-used methods

##### 2.1.1 GWAS on imputed phenotype in unlabeled data

$$\hat{\beta}^{\text{unlab}}_{\hat{Y},j} = (\boldsymbol{G}^{\text{T}}_{\text{unlab}_j}\boldsymbol{G}_{\text{unlab}_j})^{-1}\boldsymbol{G}^{\text{T}}_{\text{unlab}_j}\hat{\boldsymbol{Y}}_{\text{unlab}},$$

###### 2.1.2 GWAS on imputed phenotype in whole data

$$\begin{aligned}
\hat{\beta}^*_{\text{whole}_j} &= (\boldsymbol{G}^{\mathrm{T}}_{\text{whole}_j}\boldsymbol{G}_{\text{whole}_j})^{-1}\boldsymbol{G}^{\mathrm{T}}_{\text{whole}_j}\hat{\boldsymbol{Y}}_{\text{whole}},\\
&= (\boldsymbol{G}^{\mathrm{T}}_{\text{whole}_j}\boldsymbol{G}_{\text{whole}_j})^{-1}(\boldsymbol{G}^{\mathrm{T}}_{\text{lab}_j}\hat{\boldsymbol{Y}}_{\text{lab}} + \boldsymbol{G}^{\mathrm{T}}_{\text{unlab}_j}\hat{\boldsymbol{Y}}_{\text{unlab}})\\
&= \frac{1}{N_{\text{unlab}} + N_{\text{lab}}}(\boldsymbol{G}^{\mathrm{T}}_{\text{lab}_j}\hat{\boldsymbol{Y}}_{\text{lab}} + \boldsymbol{G}^{\mathrm{T}}_{\text{unlab}_j}\hat{\boldsymbol{Y}}_{\text{unlab}})\\
&= \frac{N_{\text{lab}}}{N_{\text{unlab}} + N_{\text{lab}}}\hat{\beta}^{\text{lab}}_{\hat{Y},j} + \frac{N_{\text{unlab}}}{N_{\text{unlab}} + N_{\text{lab}}}\hat{\beta}^{\text{unlab}}_{\hat{Y},j}
\end{aligned}$$

###### 2.1.3 GWAS on combined imputed and observed phenotype on the combined dataset

$$\begin{aligned}
\hat{\beta}^*_{\text{Combined}_j} &= (\boldsymbol{G}^{\mathrm{T}}_{\text{whole}_j}\boldsymbol{G}_{\text{whole}_j})^{-1}\boldsymbol{G}^{\mathrm{T}}_{\text{whole}_j}\boldsymbol{Y}^{\mathrm{T}}_{\text{lab}}, \hat{\boldsymbol{Y}}^{\mathrm{T}}_{\text{unlab}}]^{\mathrm{T}}\\
&= (\boldsymbol{G}^{\mathrm{T}}_{\text{whole}_j}\boldsymbol{G}_{\text{whole}_j})^{-1}\boldsymbol{G}^{\mathrm{T}}_{\text{lab}_j}\boldsymbol{Y}_{\text{lab}} + (\boldsymbol{G}^{\mathrm{T}}_{\text{whole}_j}\boldsymbol{G}_{\text{whole}_j})^{-1}\boldsymbol{G}^{\mathrm{T}}_{\text{unlab}_j}\boldsymbol{Y}_{\text{unlab}}\\
&= \frac{N_{\text{lab}}}{N_{\text{unlab}} + N_{\text{lab}}}\hat{\beta}^{\text{lab}}_{Y,j} + \frac{N_{\text{unlab}}}{N_{\text{unlab}} + N_{\text{lab}}}\hat{\beta}^{\text{unlab}}_{\hat{Y},j}
\end{aligned}$$

Next, we have two methods for accounting for phenotypic heterogeneity in meta-analysis.

###### 2.1.4 PheIMP

We consider the meta-analysis proposed in (Hormozdiari et al., 2016). To follow their notation, we denote $\tilde{Y}_i$ and $\tilde{\hat{Y}}_i$ as the standardized (variance = 1) version of $Y_i$. The assumptions are

$$\begin{bmatrix} \tilde{Y}_i \\ \tilde{\hat{Y}}_i \end{bmatrix} \sim \mathcal{N}_{\text{unlab}}\left(\begin{bmatrix} 0 \\ 0 \end{bmatrix}, \begin{bmatrix} 1 & r_{12} \\ r_{12} & 1 \end{bmatrix}\right) = \mathcal{N}_{\text{unlab}}\left(0, \begin{bmatrix} \boldsymbol{\Sigma}_{\neg 2} & \boldsymbol{r}_{\neg 22} \\ \boldsymbol{r}^{\mathrm{T}}_{\neg 22} & 1 \end{bmatrix}\right),$$

where $\boldsymbol{\Sigma}_{\neg 2} = [1]$ and $\boldsymbol{r}_{\neg 22} = [r_{12}]$.

The estimator in their proposed "Optimal Meta-analysis Strategy for Combining Imputed and Observed Values" is in the form of

$$\begin{aligned}
\hat{\beta}_{FE_j} &= \frac{N_{\text{lab}}}{N_{\text{lab}} + N_{\text{unlab}}\boldsymbol{r}^{\mathrm{T}}_{\neg 22}\Sigma^{-1}_{\neg 2}\boldsymbol{r}_{\neg 22}}\hat{\beta}^{\text{lab}}_{Y,j} + \frac{N_{\text{unlab}}\boldsymbol{r}^{\mathrm{T}}_{\neg 22}\Sigma^{-1}_{\neg 2}\boldsymbol{r}_{\neg 22}}{N_{\text{lab}} + N_{\text{unlab}}\boldsymbol{r}^{\mathrm{T}}_{\neg 22}\Sigma^{-1}_{\neg 2}\boldsymbol{r}_{\neg 22}}\hat{\beta}^{\text{unlab}}_{\hat{Y},j}\\
\text{SE}(\hat{\beta}_{FE_j}) &= \frac{1}{\sqrt{N_{\text{lab}} + N_{\text{unlab}}\boldsymbol{r}^{\mathrm{T}}_{\neg 22}\Sigma^{-1}_{\neg 2}\boldsymbol{r}_{\neg 22}}}\\
s_{FE_j} &= \frac{\hat{\beta}_{FE_j}}{\text{SE}(\hat{\beta}_{FE_j})} = \frac{N_{\text{lab}}\hat{\beta}^{\text{lab}}_{Y,j} + N_{\text{unlab}}\boldsymbol{r}^{\mathrm{T}}_{\neg 22}\Sigma^{-1}_{\neg 2}\boldsymbol{r}_{\neg 22}\hat{\beta}^{\text{unlab}}_{\hat{Y},j}}{\sqrt{N_{\text{lab}} + N_{\text{unlab}}\boldsymbol{r}^{\mathrm{T}}_{\neg 22}\Sigma^{-1}_{\neg 2}\boldsymbol{r}_{\neg 22}}}
\end{aligned}$$

After plugging in $\boldsymbol{r}^{\mathrm{T}}_{\neg 22}\Sigma^{-1}_{\neg 2}\boldsymbol{r}_{\neg 22} = r^2_{12}$, we have

$$\begin{aligned}
\hat{\beta}_{FE_j} &= \left[\frac{N_{\text{lab}}}{N_{\text{lab}} + N r^2_{12}}\hat{\beta}^{\text{lab}}_{Y,j} + \frac{N r^2_{12}}{N_{\text{lab}} + N r^2_{12}}\hat{\beta}^{\text{unlab}}_{\hat{Y},j}\right]\\
\text{SE}(\hat{\beta}_{FE_j}) &= \frac{1}{\sqrt{N_{\text{lab}} + N_{\text{unlab}}\boldsymbol{r}^{\mathrm{T}}_{\neg 22}\Sigma^{-1}_{\neg 2}\boldsymbol{r}_{\neg 22}}}\\
s_{FE_j} &= \frac{\hat{\beta}_{FE_j}}{\text{SE}(\hat{\beta}_{FE_j})} = \frac{N_{\text{lab}}\hat{\beta}^{\text{lab}}_{Y,j} + N_{\text{unlab}} r^2_{12}\hat{\beta}^{\text{unlab}}_{\hat{Y},j}}{\sqrt{N_{\text{lab}} + N_{\text{unlab}} r^2_{12}}}
\end{aligned}$$

By the unbiasedness of ordinary least squares estimation, we have

$$\mathbb{E}[\hat{\beta}_{FE_j}] = \frac{N_{\text{lab}}}{N_{\text{unlab}} r^2_{12} + N_{\text{lab}}}\,\text{Cor}(G_{ij}, Y_i) + \frac{N_{\text{unlab}} r^2_{12}}{N_{\text{unlab}} r^2_{12} + N_{\text{lab}}}\,\text{Cor}(G_{ij}, \hat{Y}_i)$$

###### 2.1.5 MTAG of GWAS on imputed phenotype in unlabeled data and observed outcome in labeled data

We follow the notations, model, and assumptions of multi-trait analysis of GWAS (MTAG) (Turley et al., 2018) to derive its estimator $\widehat{\boldsymbol{\beta}}_{\text{MTAG,j,1}}$. Here, we considered using MTAG for GWAS on imputed phenotype in unlabeled data and observed outcome in labeled data. MTAG assumes a random effects model:

$$\tilde{Y}_i = \sum_j^M X_{ij} b_{1j} + \epsilon_{Y_i}$$
$$\tilde{\hat{Y}}_q = \sum_j^M X_{qj} b_{2j} + \epsilon_{\hat{Y}_q},$$

where

$$\text{Var}\left(\begin{bmatrix} b_{1j} \\ b_{2j} \end{bmatrix}\right) = \frac{1}{M}\begin{bmatrix} h_Y^2 & \rho\sqrt{h_Y^2 h_{\hat{Y}}^2} \\ \rho\sqrt{h_Y^2 h_{\hat{Y}}^2} & h_{\hat{Y}}^2 \end{bmatrix} = \boldsymbol{\Omega}$$

The variancec-covariance matrix of the estimation error is denoted as $\text{Var}\left(\widehat{\boldsymbol{\beta}}_j \mid \boldsymbol{\beta}_j\right) = \boldsymbol{\Sigma}_j$. We have the MTAG effect for SNP $j$ on $Y_i$ (where $t = 1$ in the eqaution below) is

$$\widehat{\boldsymbol{\beta}}_{\text{MTAG,j},t} = \frac{\frac{\boldsymbol{\omega}_t^{\text{T}}}{\omega_{tt}}\left(\boldsymbol{\Omega} - \frac{\boldsymbol{\omega}_t\boldsymbol{\omega}_t^{\text{T}}}{\omega_{tt}} + \boldsymbol{\Sigma}_j\right)^{-1}}{\frac{\boldsymbol{\omega}_t^{\text{T}}}{\omega_{tt}}\left(\boldsymbol{\Omega} - \frac{\boldsymbol{\omega}_t\boldsymbol{\omega}_t^{\text{T}}}{\omega_{tt}} + \boldsymbol{\Sigma}_j\right)^{-1}\frac{\boldsymbol{\omega}_t}{\omega_{tt}}}\widehat{\boldsymbol{\beta}}_j$$

where

$$\frac{\boldsymbol{\omega}_t^{\text{T}}}{\omega_{tt}} = [1, \rho\sqrt{\frac{h_{\hat{Y}}^2}{h_Y^2}}]$$

$$\left(\boldsymbol{\Omega} - \frac{\boldsymbol{\omega}_t\boldsymbol{\omega}_t^{\text{T}}}{\omega_{tt}} + \boldsymbol{\Sigma}_j\right) = \begin{bmatrix} h_Y^2 & \rho\sqrt{h_Y^2 h_{\hat{Y}}^2} \\ \rho\sqrt{h_Y^2 h_{\hat{Y}}^2} & h_{\hat{Y}}^2 \end{bmatrix} - \begin{bmatrix} h_Y^2 & \rho\sqrt{h_Y^2 h_{\hat{Y}}^2} \\ \rho\sqrt{h_Y^2 h_{\hat{Y}}^2} & \rho^2 h_{\hat{Y}}^2 \end{bmatrix} + \begin{bmatrix} \frac{1}{N_{\text{lab}}} & 0 \\ 0 & \frac{1}{N_{\text{unlab}}} \end{bmatrix}$$
$$= \begin{bmatrix} \frac{1}{N_{\text{lab}}} & 0 \\ 0 & (1-\rho^2 h_{\hat{Y}}^2) + \frac{1}{N_{\text{unlab}}} \end{bmatrix}$$

$$\frac{\boldsymbol{\omega}_t^{\text{T}}}{\omega_{tt}}\left(\boldsymbol{\Omega} - \frac{\boldsymbol{\omega}_t\boldsymbol{\omega}_t^{\text{T}}}{\omega_{tt}} + \boldsymbol{\Sigma}_j\right)^{-1} = [N_{\text{lab}}, \rho\sqrt{\frac{h_{\hat{Y}}^2}{h_Y^2}}\frac{1}{(1-\rho^2 h_{\hat{Y}}^2) + \frac{1}{N_{\text{unlab}}}}] = [N_{\text{lab}}, \rho\sqrt{h_{\hat{Y}}^2 h_Y^2}\frac{1}{h_Y^2[(1-\rho^2 h_{\hat{Y}}^2) + \frac{1}{N_{\text{unlab}}}]}]$$

$$\frac{\boldsymbol{\omega}_t^{\text{T}}}{\omega_{tt}}\left(\boldsymbol{\Omega} - \frac{\boldsymbol{\omega}_t\boldsymbol{\omega}_t^{\text{T}}}{\omega_{tt}} + \boldsymbol{\Sigma}_j\right)^{-1}\frac{\boldsymbol{\omega}_t}{\omega_{tt}} = N_{\text{lab}} + \frac{\rho^2 h_{\hat{Y}}^2}{h_Y^2[(1-\rho^2 h_{\hat{Y}}^2) + \frac{1}{N_{\text{unlab}}}]}$$

Therefore,

$$\widehat{\beta}_{\text{MTAG,j,1}} = \left[\frac{N_{\text{lab}}}{N_{\text{lab}} + \frac{\rho^2 h_{\hat{Y}}^2}{h_Y^2[(1-\rho^2 h_{\hat{Y}}^2) + \frac{1}{N_{\text{unlab}}}]}}\widehat{\beta}_{Y,j}^{\text{lab}} + \frac{\rho\sqrt{h_{\hat{Y}}^2 h_Y^2}\frac{1}{h_Y^2[(1-\rho^2 h_{\hat{Y}}^2) + \frac{1}{N_{\text{unlab}}}]}}{N_{\text{lab}} + \frac{\rho^2 h_{\hat{Y}}^2}{h_Y^2[(1-\rho^2 h_{\hat{Y}}^2) + \frac{1}{N_{\text{unlab}}}]}}\widehat{\beta}_{\text{unlab}_j}\right]$$

By the unbiasedness of ordinary least squares estimation, we have

$$\mathbb{E}[\widehat{\beta}_{\mathrm{MTAG,j,1}}] = \left[ \frac{N_{\mathrm{lab}}}{N_{\mathrm{lab}} + \dfrac{\rho^2 h^2_{\hat{Y}}}{h^2_Y[(1-\rho^2 h^2_{\hat{Y}}) + \dfrac{1}{N_{\mathrm{unlab}}}]}} \mathrm{Cor}(G_{ij}, Y_i) + \frac{\rho\sqrt{h^2_{\hat{Y}} h^2_Y}\dfrac{1}{h^2_Y[(1-\rho^2 h^2_{\hat{Y}}) + \dfrac{1}{N_{\mathrm{unlab}}}]}}{N_{\mathrm{lab}} + \dfrac{\rho^2 h^2_{\hat{Y}}}{h^2_Y[(1-\rho^2 h^2_{\hat{Y}}) + \dfrac{1}{N_{\mathrm{unlab}}}]}} \mathrm{Cor}(G_{ij}, \hat{Y}_i) \right]$$

#### 2.2 Theoretical comparison of commonly-applied methods

**Proposition 1.** *The estimator for the four methods discussed above can all be rewritten as the non-negative weighted sum of three GWAS estimators*

$$w_1 \hat{\beta}^{\mathrm{lab}}_{Y,j} + w_2 \hat{\beta}^{\mathrm{unlab}}_{\hat{Y},j} + w_3 \hat{\beta}^{\mathrm{lab}}_{\hat{Y},j},$$

*where $w_1, w_2, w_3$ are all non-negative weights.*

*Proof.* This can be proved on the basis of our derivation above. □

**Proposition 2.** *The estimator for the four methods discussed above is unbiased and therefore confidence valid (has no type I error inflation) if and only if*

$$\mathrm{Cor}(G_{ij}, \hat{Y}_i) = C\,\mathrm{Cor}(G_{ij}, Y_i).$$

*Here, $C$ depends on specific methods: $C = 1$ for the first three methods and $C = \rho\sqrt{\frac{h^2_{\hat{Y}}}{h^2_Y}}$ for MTAG, where $\rho$ is the genetic correlation between $Y_i$, $h^2_{\hat{Y}}$ is the heritability for $\hat{Y}_i$, and $h^2_Y$ is the heritability for $Y_i$.*

*Proof.* The estimator is unbiased if its expectation is $\beta_j$. For MTAG estimator

$$\mathbb{E}[\widehat{\beta}_{\mathrm{MTAG,j,1}}] - \beta_j = 0 \Leftrightarrow \mathrm{Cor}(G_{ij}, \hat{Y}_i) = \rho\sqrt{\frac{h^2_{\hat{Y}}}{h^2_Y}}\,\mathrm{Cor}(G_{ij}, Y_i)$$

For other estimators, since the summation of weights is 1, they are unbiased if and only if $\mathrm{Cor}(G_{ij}, \hat{Y}_i) = \mathrm{Cor}(G_{ij}, Y_i)$. □

### 3 `POP-GWAS` (Post-prediction GWAS)

Here, we present our `POP-GWAS` method for ML-assisted GWAS analysis. We start with GWAS on quantitative phenotypes using unrelated samples, but later extend it to binary traits and linear mixed models to account for sample relatedness.

#### 3.1 Method

The `POP-GWAS` estimator is

$$\hat{\beta}_{\mathrm{Pop},j} = r\frac{N_{\mathrm{unlab}}}{N_{\mathrm{unlab}} + N_{\mathrm{lab}}}\hat{\beta}^{\mathrm{unlab}}_{\hat{Y},j} + \hat{\beta}^{\mathrm{lab}}_{Y,j} - r\frac{N_{\mathrm{unlab}}}{N_{\mathrm{unlab}} + N_{\mathrm{lab}}}\hat{\beta}^{\mathrm{lab}}_{\hat{Y},j}, \tag{1}$$

Its corresponded standard error is

$$\begin{aligned}
\mathrm{SE}(\hat{\beta}_{\mathrm{Pop},j}) &= \sqrt{\mathrm{Var}(r\frac{N_{\mathrm{unlab}}}{N_{\mathrm{unlab}}+N_{\mathrm{lab}}}\hat{\beta}^{\mathrm{unlab}}_{\widehat{Y},j}+\hat{\beta}^{\mathrm{lab}}_{Y,j}-r\frac{N_{\mathrm{unlab}}}{N_{\mathrm{unlab}}+N_{\mathrm{lab}}}\hat{\beta}^{\mathrm{lab}}_{\widehat{Y},j})} \\
&= \sqrt{(r\frac{N_{\mathrm{unlab}}}{N_{\mathrm{unlab}}+N_{\mathrm{lab}}})^2\,\mathrm{Var}(\hat{\beta}^{\mathrm{unlab}}_{\widehat{Y},j})+\mathrm{Var}(\hat{\beta}^{\mathrm{lab}}_{Y,j})+(r\frac{N_{\mathrm{unlab}}}{N_{\mathrm{unlab}}+N_{\mathrm{lab}}})^2\,\mathrm{Var}(\hat{\beta}^{\mathrm{lab}}_{\widehat{Y},j})-2r\frac{N_{\mathrm{unlab}}}{N_{\mathrm{unlab}}+N_{\mathrm{lab}}}\,\mathrm{Cov}(\hat{\beta}^{\mathrm{lab}}_{Y,j},\hat{\beta}^{\mathrm{lab}}_{\widehat{Y},j})} \\
&\approx \sqrt{(r\frac{N_{\mathrm{unlab}}}{N_{\mathrm{unlab}}+N_{\mathrm{lab}}})^2\frac{1}{N_{\mathrm{unlab}}}+\frac{1}{N_{\mathrm{lab}}}+(r\frac{N_{\mathrm{unlab}}}{N_{\mathrm{unlab}}+N_{\mathrm{lab}}})^2\frac{1}{N_{\mathrm{lab}}}-2r\frac{N_{\mathrm{unlab}}}{N_{\mathrm{unlab}}+N_{\mathrm{lab}}}\frac{r}{N_{\mathrm{lab}}}} \\
&\approx \sqrt{\frac{1}{N_{\mathrm{lab}}}-r^2\frac{N_{\mathrm{unlab}}}{(N_{\mathrm{unlab}}+N_{\mathrm{lab}})N_{\mathrm{lab}}}}
\end{aligned}$$

Therefore, The effective sample size can be calculated as

$$N_{\mathrm{eff}}=\frac{1}{\mathrm{SE}(\hat{\beta}_{\mathrm{Pop},j})^2}=\frac{N_{\mathrm{lab}}}{1-r^2\dfrac{N_{\mathrm{unlab}}}{N_{\mathrm{unlab}}+N_{\mathrm{lab}}}}$$

Our implemented algorithm is

**Algorithm 1** `POP-GWAS` for quantitative phenotype

**Input:** Z-score and sample size from three GWAS summary statistics: $(z^{\mathrm{lab}}_{Y,j},N_{\mathrm{lab}}),(z^{\mathrm{lab}}_{\widehat{Y},j},N_{\mathrm{lab}})$, and $(z^{\mathrm{unlab}}_{\widehat{Y},j},N_{\mathrm{unlab}})$ for all SNPs $j=1,\cdots,J$

1: Convert Z-score into effect size

$$\hat{\beta}^{\mathrm{lab}}_{Y,j}=\frac{z^{\mathrm{lab}}_{Y,j}}{\sqrt{N_{\mathrm{lab}}}},\hat{\beta}^{\mathrm{lab}}_{\widehat{Y},j}=\frac{z^{\mathrm{lab}}_{\widehat{Y},j}}{\sqrt{N_{\mathrm{lab}}}},\text{and }\hat{\beta}^{\mathrm{unlab}}_{\widehat{Y},j}=\frac{z^{\mathrm{unlab}}_{\widehat{Y},j}}{\sqrt{N_{\mathrm{unlab}}}}\text{ for all SNPs } j=1,\cdots,J$$

2: Apply bivariate-LD score regression on $(z^{\mathrm{lab}}_{Y,j},N_{\mathrm{lab}})$ and $(z^{\mathrm{lab}}_{\widehat{Y},j},N_{\mathrm{lab}})$ to get $r$ from the intercept.

3: Get `POP-GWAS` summary statistics for all SNPs $j=1,\cdots,J$.

$$\begin{aligned}
\hat{\beta}_{\mathrm{Pop},j} &= r\frac{N_{\mathrm{unlab}}}{N_{\mathrm{unlab}}+N_{\mathrm{lab}}}\hat{\beta}^{\mathrm{unlab}}_{\widehat{Y},j}+\hat{\beta}^{\mathrm{lab}}_{Y,j}-r\frac{N_{\mathrm{unlab}}}{N_{\mathrm{unlab}}+N_{\mathrm{lab}}}\hat{\beta}^{\mathrm{lab}}_{\widehat{Y},j} \\
\mathrm{SE}(\hat{\beta}_{\mathrm{Pop},j}) &= \sqrt{\frac{1}{N_{\mathrm{lab}}}-r^2\frac{N_{\mathrm{unlab}}}{(N_{\mathrm{unlab}}+N_{\mathrm{lab}})N_{\mathrm{lab}}}} \\
z_{\mathrm{Pop},j} &= \hat{\beta}_{\mathrm{Pop},j}/\,\mathrm{SE}(\hat{\beta}_{\mathrm{Pop},j}) \\
p_{\mathrm{Pop},j} &= 2\times[1-\Phi(z_{\mathrm{Pop},j})]
\end{aligned}$$

Here, $\Phi$ is the cumulative distribution function for a standard normal distribution.

**Output:** `POP-GWAS` summary statistics

Note that if the user specifies the effective allele frequency for $j$-th SNP $p_j$, we will output the beta corresponding to the per-allele increase in s.d. units of phenotype: $\hat{\beta}_{\mathrm{Pop},j}/\sqrt{2p_j(1-p_j)}$ and its corresponding standard error $\mathrm{SE}(\hat{\beta}_{\mathrm{Pop},j})/\sqrt{2p_j(1-p_j)}$.

#### 3.2 Theory on unbiasedness, consistency, and asymptotic normality

In this section, we establish the theoretical guarantees for valid inference (Miao et al., 2023). In the following, $\xrightarrow{P}$ denotes convergence in probability and $\xrightarrow{D}$ denotes convergence in distribution.

We first establish the unbiasedness for $\hat{\beta}_{\mathrm{Pop},j}$:

**Proposition 3.** *$\hat{\beta}_{\mathrm{Pop},j}$ is unbiased for true marginal effects of $j$-th SNP*

$$\mathbb{E}[\hat{\beta}_{\mathrm{Pop},j}]=\beta_j$$

*Proof.* See Appendix □

Next, we establish the consistency of $\hat{\beta}_{\text{Pop},j}$

**Proposition 4.** *As* $\dfrac{N_{\text{lab}}}{N_{\text{unlab}}} \to \rho \in (0, \infty)$ *and* $N_{\text{lab}} \to \infty$*,* $\hat{\beta}_{\text{Pop},j}$ *is consistent for the true marginal effect of* $j$*-th SNP*

$$\hat{\beta}_{\text{Pop},j} \xrightarrow{P} \beta_j$$

*Proof.* See Appendix □

Furthermore, we establish the asymptotic normality of $\hat{\beta}_{\text{Pop},j}$

**Theorem 1.** *As* $\dfrac{N_{\text{lab}}}{N_{\text{unlab}}} \to \rho \in (0, \infty)$ *and* $N_{\text{lab}} \to \infty$*,* $\hat{\beta}_{\text{Pop},j}$ *follows asymptotically normal distribution*

$$\sqrt{N_{\text{lab}}} V^{-\dfrac{1}{2}} \left( \hat{\beta}_{\text{Pop},j} - \beta_j \right) \xrightarrow{D} \mathcal{N}_{\text{unlab}}(0, 1)$$

*as* $min(N_{\text{lab}}, N_{\text{unlab}}) \to \infty$*, where* $V = (r \dfrac{N_{\text{unlab}}}{N_{\text{unlab}} + N_{\text{lab}}})^2 \operatorname{Var}(\hat{\beta}^{\text{lab}}_{\hat{Y},j}) + \operatorname{Var}(\hat{\beta}^{\text{lab}}_{Y,j}) + \dfrac{(r \dfrac{N_{\text{unlab}}}{N_{\text{unlab}} + N_{\text{lab}}})^2}{N_{\text{unlab}}/N_{\text{lab}}} \operatorname{Var}(\hat{\beta}^{\text{unlab}}_{\hat{Y},j}) - 2r \dfrac{N_{\text{unlab}}}{N_{\text{unlab}} + N_{\text{lab}}} \operatorname{Cov}(\hat{\beta}^{\text{lab}}_{\hat{Y},j}, \hat{\beta}^{\text{lab}}_{Y,j})$.

*Proof.* See Appendix □

#### 3.3 Theory on efficiency

In this section, we establish the theoretical guarantees for powerful inference.

**Proposition 5.** *The relative efficiency between the* $\hat{\beta}_{\text{Pop},j}$ *and* $\hat{\beta}^{\text{lab}}_{Y,j}$ *is*

$$\frac{\operatorname{Var}(\hat{\beta}_{\text{Pop},j})}{\operatorname{Var}(\hat{\beta}^{\text{lab}}_{Y,j})} = \frac{1}{1 - r^2 \dfrac{N_{\text{unlab}}}{N_{\text{unlab}} + N_{\text{lab}}}} \leq 1$$

*Proof.* See Appendix □

**Remark 1.** This shows that `POP-GWAS` is more effective than conventional GWAS based on observed phenotypes when the ML prediction accuracy is non-zero. It is equally effective as observed phenotype GWAS when ML prediction accuracy is is non-informative.

Next, we show that statistical optimality for `POP-GWAS`:

**Theorem 2** (Gauss Markov theorem)**.** *The* $\hat{\beta}_{\text{Pop},j}$ *has the lowest sampling variance within the class of linear unbiased estimators*

$$\hat{\beta}_j = \sum_{i=1}^{N_{\text{lab}}} q_{1i} Y_{i,lab} + \sum_{i=1}^{N_{\text{lab}}} q_{2i} \hat{Y}_{i,lab} + \sum_{i=N_{\text{lab}}+1}^{N_{\text{unlab}}+N_{\text{lab}}} q_{3i} \hat{Y}_{i,unlab}$$

*Proof.* See Appendix □

**Remark 2.** This suggests that any attempt to improve `POP-GWAS` with a linear estimator would result in either estimation bias or lower efficiency. This conclusion leads to a closed-form formula for the upper bound on the effective sample size of a valid ML-assisted GWAS. This conclusion leads to a closed-form formula for the upper bound on the effective sample size of a valid ML-assisted GWAS.

$$N_{\text{eff}} = \frac{N_{\text{lab}}}{1 - \dfrac{r^2 N_{\text{unlab}}}{N_{\text{unlab}} + N_{\text{lab}}}}$$

##### 3.4 Connection with other methods

With additional assumptions, POP-GWAS has several useful special cases and links to existing methods.

###### 3.4.1 Conventional GWAS on observed outcome

After setting $r = 0$, we have

$$\hat{\beta}_{\text{Pop},j} = \hat{\beta}^{\text{lab}}_{Y,j}$$

###### 3.4.2 Inverse-variance-weighted meta-analysis

After setting $r = 1$, we have

$$\begin{aligned}
\hat{\beta}_{\text{Pop},j} &= \hat{\beta}^{\text{lab}}_{Y,j} + \frac{N_{\text{unlab}}}{N_{\text{unlab}} + N_{\text{lab}}}\hat{\beta}^{\text{unlab}}_{\widehat{Y},j} - \frac{N_{\text{unlab}}}{N_{\text{unlab}} + N_{\text{lab}}}\hat{\beta}^{\text{lab}}_{\widehat{Y},j} \\
&= \hat{\beta}^{\text{lab}}_{Y,j} + \frac{N_{\text{unlab}}}{N_{\text{unlab}} + N_{\text{lab}}}\hat{\beta}^{\text{unlab}}_{\widehat{Y},j} - \frac{N_{\text{unlab}}}{N_{\text{unlab}} + N_{\text{lab}}}\hat{\beta}^{\text{lab}}_{Y,j} \\
&= \frac{N_{\text{lab}}}{N_{\text{unlab}} + N_{\text{lab}}}\hat{\beta}^{\text{lab}}_{Y,j} + \frac{N_{\text{unlab}}}{N_{\text{unlab}} + N_{\text{lab}}}\hat{\beta}^{\text{unlab}}_{\widehat{Y},j} \\
&\approx \frac{\frac{1}{\text{SE}(\hat{\beta}^{\text{lab}}_{Y,j})^2}}{\frac{1}{\text{SE}(\hat{\beta}^{\text{lab}}_{Y,j})^2} + \frac{1}{\text{SE}(\hat{\beta}^{\text{unlab}}_{\widehat{Y},j})^2}}\hat{\beta}^{\text{lab}}_{Y,j} + \frac{\frac{1}{\text{SE}(\hat{\beta}^{\text{unlab}}_{\widehat{Y},j})^2}}{\frac{1}{\text{SE}(\hat{\beta}^{\text{lab}}_{Y,j})^2} + \frac{1}{\text{SE}(\hat{\beta}^{\text{unlab}}_{\widehat{Y},j})^2}}\hat{\beta}^{\text{unlab}}_{\widehat{Y},j}
\end{aligned}$$

This is the formula for inverse-variance weighted meta-analysis of $\hat{\beta}^{\text{lab}}_{Y,j}$ and $\hat{\beta}^{\text{unlab}}_{\widehat{Y},j}$.

### 4 Extensions of POP-GWAS

#### 4.1 Binary traits

While the POP-GWAS estimator is ideally suited for z-scores from linear regression, we show here that z-scores from logistic regression closely approximate those from linear regression at large sample sizes. As a result, the use of summary statistics based on logistic regression should also provide accurate estimation and inference, which is supported by our simulation results. Recall that linear regression Z-scores are given by:

$$Z_j^{\text{linear}} = \frac{\hat{\beta}_j^{\text{linear}}}{\sqrt{\text{Var}\left(\hat{\beta}_j^{\text{linear}}\right)}} \approx \frac{1}{\sqrt{N_{\text{total}}}}\sum_i Y_i G_{ij}$$

where $Y_i$ as the standardized binary phenotype with mean of 0 and variance of 1 of individual $i$, where $N_{\text{total}} = N_{\text{case}} + N_{\text{control}}$. Logistic regression Z-scores are given by:

$$Z_j^{\text{logistic}} = \frac{\hat{\beta}_j^{\text{logistic}}}{\sqrt{\text{Var}\left(\hat{\beta}_j^{\text{logistic}}\right)}},$$

where $\hat{\beta}_j^{\text{logistic}}$ is the estimate of the logistic regression coefficient of SNP $j$. The following lemma derived in (Weissbrod et al., 2018) states that logistic regression z-scores are approximately the same as linear regression z-scores under large sample sizes.

**Lemma 2** (Weissbrod et al. 2018)**.** *Under large sample size and small SNP effect assumption, we have*

$$Z_j^{\textit{linear}} \approx Z_j^{\textit{logistic}}$$

Similarly, the effect size from linear regression can be transformed to logistic regression under the assumption of large sample size and small SNP effect. This has been shown in (Pirinen et al., 2013) and is explained below:

**Lemma 3** (Pirinen et al. 2013). *Under large sample size and small SNP effect assumption, we have the log-odds*

$$\hat{\beta}_j^{logistic} \approx \widehat{\beta}_j^{linear} \left( \phi(1-\phi) + 0.5(1-2\phi)(1-2p_j)\widehat{\beta}_j^{linear} - \frac{0.084 + 0.9\phi(1-2\phi)p_j(1-p_j)}{\phi(1-\phi)} \widehat{\beta}_j^{linear\ 2} \right)^{-1},$$

*where $\phi$ is the proportion of the cases in the data, $p_j$ is the reference allele frequency in the data.*

Using the formula above sometimes results in the inflated OR, so we follow BOLT-LMM (Loh et al., 2015) to use a simple version of it:

$$\hat{\beta}_j^{\text{logistic}} \approx \frac{\widehat{\beta}_j^{\text{linear}}}{\phi(1-\phi)}, \tag{2}$$

Then, we have our `POP-GWAS` algorithm for binary phenotype:

**Algorithm 2** `POP-GWAS` for binary phenotype

**Input:** Z-score and sample size from three GWAS summary statistics: $(z_{Y,j}^{\text{lab}}, N_{\text{lab}}), (z_{\widehat{Y},j}^{\text{lab}}, N_{\text{lab}})$, , $(z_{\widehat{Y},j}^{\text{unlab}}, N_{\text{unlab}})$ and the effective allele frequency $p_j$ for all SNPs $j = 1, \cdots, J$; Proportions of cases $\phi$ in the labeled data.

1: Apply the algorithm 1

2: Apply equation 2 to get the $\hat{\beta}_{\text{Pop},j}$ and its corresponding standard error.

**Output:** `POP-GWAS` summary statistics for binary phenotype

#### 4.2 Sample relatedness and population structure

To account for population structure and sample relatedness, the linear mixed model is often used in GWAS. Although our derivation for `POP-GWAS` is based on linear regression, it's a common practice in the GWAS community to treat summary statistics from linear mixed models as if they were from linear models. Mathematically speaking, GWAS association results derived using the linear mixed model are equivalent to those obtained using the marginal linear model on phenotypic residuals after adjustment for best linear unbiased prediction (BLUP). These phenotypic residuals essentially represent the phenotype after adjusting for sample relatedness as captured by the genetic relationship matrix.

For example, BOLT-LMM uses a two-step method for linear mixed model GWAS. The algorithm fits a Gaussian mixture model of the SNP, using a fast variational approximation to compute approximate phenotypic residuals. It then tests these residuals for association with potential markers using a retrospective score statistic. Consequently, BOLT-LMM's GWAS results are derived from the marginal linear regression between the phenotypic residuals and the SNP. Similarly, Regenie (Mbatchou et al., 2021) first regresses the leave-one-chr-out polygenic score from the phenotype. It then performs a simple linear regression between the phenotypic residuals and the SNP.

Since `POP-GWAS` is based on summary statistics, our algorithms can be applied directly to summary statistics derived from the linear mixed model for quantitative and binary phenotypes, respectively. However, to account for the differences between linear regression and the linear mixed model, it is essential to enter the effective sample size (for the linear mixed model) in `POP-GWAS`.

#### 4.3 Sample overlap

Next, we describe how to deal with the GWAS sample overlap problem. We assume that there are $N_{\text{ovp}}$ individuals in both the labeled and unlabeled data. Recall that the three GWAS summary statistics can be expressed as

$$\hat{\beta}^{\text{lab}}_{Y,j} = \frac{1}{N_{\text{lab}}}\sum_{i=1}^{N_{\text{lab}}} X_{\text{lab}_i}Y_{\text{lab}_i} = \frac{1}{N_{\text{lab}}}[\sum_{i\in\{N_{\text{lab}}-N_{\text{ovp}}\}} X_{\text{lab}_i}Y_{\text{lab}_i} + \sum_{i\in\{N_{\text{ovp}}\}} X_{\text{ovp}_i}Y_{\text{ovp}_i}]$$

$$\hat{\beta}^{\text{lab}}_{\hat{Y},j} = \frac{1}{N_{\text{lab}}}\sum_{i=1}^{N_{\text{lab}}} X_{\text{lab}_i}\hat{Y}_{\text{lab}_i} = \frac{1}{N_{\text{lab}}}[\sum_{i\in\{N_{\text{lab}}-N_{\text{ovp}}\}} X_{\text{lab}_i}\hat{Y}_{\text{lab}_i} + \sum_{i\in\{N_{\text{ovp}}\}} X_{\text{ovp}_i}\hat{Y}_{\text{ovp}_i}]$$

$$\hat{\beta}^{\text{unlab}}_{\hat{Y},j} = \frac{1}{N_{\text{unlab}}}\sum_{i=N_{\text{lab}}+1}^{N_{\text{unlab}}+N_{\text{lab}}} X_{\text{unlab}_i}Y_{\text{unlab}_i} = \frac{1}{N_{\text{unlab}}}[\sum_{i\in\{N_{\text{unlab}}-N_{\text{ovp}}\}} X_{\text{unlab}_i}\hat{Y}_{\text{unlab}_i} + \sum_{i\in\{N_{\text{ovp}}\}} X_{\text{ovp}_i}\hat{Y}_{\text{ovp}_i}]$$

The `POP-GWAS` estimator with weight $c$ is then

$$\begin{aligned}\hat{\beta}_{\text{Pop},j} &= \hat{\beta}^{\text{lab}}_{Y,j} + c\hat{\beta}^{\text{unlab}}_{\hat{Y},j} - c\hat{\beta}^{\text{lab}}_{\hat{Y},j} \\ &= \sum_{i\in\{N_{\text{ovp}}\}} X_{\text{ovp}_i}[\frac{1}{N_{\text{lab}}}Y_{\text{ovp}_i} - c(\frac{1}{N_{\text{lab}}} - \frac{1}{N_{\text{unlab}}})\hat{Y}_{\text{ovp}_i}] + \frac{1}{N_{\text{lab}}}\sum_{i\in\{N_{\text{lab}}-N_{\text{ovp}}\}} X_{\text{lab}_i}(Y_{\text{lab}_i} - c\hat{Y}_{\text{lab}_i}) + \\ &\quad c\frac{1}{N_{\text{unlab}}}\sum_{i\in\{N_{\text{unlab}}-N_{\text{ovp}}\}} X_{\text{unlab}_i}\hat{Y}_{\text{unlab}_i}\end{aligned}$$

Its standard error could be calculated as

$$\text{SE}(\hat{\beta}_{\text{Pop},j}) = \sqrt{\frac{1}{N_{\text{lab}}} + c^2(\frac{1}{N_{\text{unlab}}} + \frac{1}{N_{\text{lab}}} - \frac{2N_{\text{ovp}}}{N_{\text{lab}}N_{\text{unlab}}}) - 2cr(\frac{1}{N_{\text{lab}}} - \frac{N_{\text{ovp}}}{N_{\text{lab}}N_{\text{unlab}}})}$$

which reaches its minimum when

$$c = \frac{N_{\text{unlab}} - N_{\text{ovp}}}{N_{\text{unlab}} + N_{\text{lab}} - 2N_{\text{ovp}}}r$$

The corrsponding SE is then

$$\begin{aligned}\text{SE}(\hat{\beta}_{\text{Pop},j}) &= \sqrt{\frac{1}{N_{\text{lab}}} - \frac{r^2(\frac{1}{N_{\text{lab}}} - \frac{N_{\text{ovp}}}{N_{\text{lab}}N_{\text{unlab}}})^2}{\frac{1}{N_{\text{unlab}}} + \frac{1}{N_{\text{lab}}} - \frac{2N_{\text{ovp}}}{N_{\text{lab}}N_{\text{unlab}}}}} \\ &= \sqrt{\frac{1}{N_{\text{lab}}} - \frac{r^2(\frac{N_{\text{unlab}} - N_{\text{ovp}}}{N_{\text{lab}}N_{\text{unlab}}})^2}{\frac{1}{N_{\text{unlab}}} + \frac{1}{N_{\text{lab}}} - \frac{2N_{\text{ovp}}}{N_{\text{lab}}N_{\text{unlab}}}}} \\ &= \sqrt{\frac{1}{N_{\text{lab}}} - \frac{r^2(N_{\text{unlab}} - N_{\text{ovp}})^2}{N_{\text{lab}}N_{\text{unlab}}(N_{\text{unlab}} + N_{\text{lab}} - 2N_{\text{ovp}})}}\end{aligned}$$

When there is no sample overlap (i.e., $N_{\text{ovp}} = 0$), $c = \frac{N_{\text{unlab}}}{N_{\text{unlab}} + N_{\text{lab}}}r$. Therefore, it degenerates to the `POP-GWAS` with no sample overlap in equation (1).

### 5 Power and sample size calculations for ML-assisted GWAS

We derive the formula for power calculation under an additive model where the effect of each SNP is small, which is a common assumption in most GWAS.

#### 5.1 Quantitative phenotype

For quantitative traits, the power calculation formula incorporates:

- Effect size (the change in the raw phenotype value associated with each copy of the allele): $\theta$
- Effective allele frequency: $p$
- Sample size of labeled data: $N_{\text{lab}}$
- Sample size of unlabeled data: $N_{\text{unlab}}$
- Imputation correlation of the ML-algorithm: $r$
- Phenotypic variance: $\sigma^2$
- Significance threshold: $\alpha$

The formula for power calculation is

$$\text{Power } = 1 - \Phi\left(z_{1-\alpha/2} - \sqrt{N_{\text{eff}} 2p(1-p)}\frac{\theta}{\sigma}\right),$$

where $N_{\text{eff}} = \dfrac{N_{\text{lab}}}{1 - \dfrac{r^2 N_{\text{unlab}}}{N_{\text{unlab}} + N_{\text{lab}}}}$ and $\Phi$ is the cumulative distribution function for a standard normal distribution.

#### 5.2 Binary phenotype

For binary traits, the power calculation formula incorporates:

- Odds ratio (the per-allele increase in the odds of the disease): $\theta_{\text{OR}}$
- Effective allele frequency: $p$
- Sample size of labeled data: $N_{\text{lab}}$
- Sample size of unlabeled data: $N_{\text{unlab}}$
- Imputation correlation of the ML-algorithm: $r$
- Proportion of the case: $\phi$
- Significance threshold: $\alpha$

The formula for power calculation (Vukcevic et al., 2011) is

$$\text{Power } = 1 - \Phi\left[z_{1-\alpha/2} - \sqrt{N_{\text{eff}} 2p(1-p)\phi(1-\phi)}\log(\theta_{\text{OR}})\right],$$

where $N_{\text{eff}} = \dfrac{N_{\text{lab}}}{1 - \dfrac{r^2 N_{\text{unlab}}}{N_{\text{unlab}} + N_{\text{lab}}}}$ and $\Phi$ is the cumulative distribution function for a standard normal distribution.

Next, we consider the sample size calculations and imputation correlation calculations for a given power. We will

#### 5.3 Sample size calculations

We first calculate the effective sample size based on power.

For quantitative traits, we have

$$N_{\text{eff}} = \frac{\left[z_{1-\alpha/2} - \Phi^{-1}(1 - \text{Power})\right]^2}{2p(1-p)\dfrac{\theta^2}{\sigma^2}}$$

For binary traits, we have

$$N_{\text{eff}} = \frac{\left[z_{1-\alpha/2} - \Phi^{-1}(1 - \text{Power})\right]^2}{2p(1-p)\phi(1-\phi)[\log(\theta_{\text{OR}})]^2}$$

We have then two different situations:

- Given $N_{\text{lab}}$ and $r$,

$$N_{\text{unlab}} = \frac{N_{\text{lab}}\left(N_{\text{unlab}} + N_{\text{lab}}\right)}{N_{\text{eff}}(1-r^2) + N_{\text{lab}}}$$

- Given $N_{\text{unlab}}$ and $r$

$$N_{\text{lab}} = \frac{N_{\text{eff}}}{2} - \frac{N_{\text{unlab}}}{2} + \frac{\sqrt{N_{\text{eff}}^2 - 4N_{\text{eff}}N_{\text{unlab}}r^2 + 2N_{\text{eff}}N_{\text{unlab}} + N_{\text{unlab}}^2}}{2}$$

##### 5.4 Imputation correlation calculations

Based on the effective sample size given above, we have

$$r = \sqrt{\frac{\left(N_{\text{eff}} - N_{\text{lab}}\right)\left(N_{\text{lab}} + N_{\text{unlab}}\right)}{N_{\text{eff}}N_{\text{unlab}}}}$$

This could give us the required imputation accuracy for the ML algorithm.

#### 6 Impact of GWAS covariates on ML-assisted GWAS

##### 6.1 Summary

The literature on ML-assisted GWAS typically reports the correlation (or $R^2$) between the observed and imputed phenotypes to quantify the accuracy of the phenotype imputation. However, we argue that this is not a meaningful metric for quantifying the power gain of ML-assisted GWAS, despite many papers using it to calculate the effective sample size. This is because almost all GWAS adjust for covariates such as sex, age, and genetic principal components. Other covariates, like body mass index (BMI) and height, are also adjusted due to the specific problems of interest in the study. Therefore, if these GWAS covariates are used as imputation variables, they may yield a high correlation $r$, but result in almost no power gain in the ML-assisted GWAS since they are already adjusted for in the GWAS. For example, if we use only biological sex to impute the phenotype and perform the GWAS of the imputed phenotype, we should expect to find no associations for autosomal SNPs, even if this results in a high imputation $r$. Therefore, a more meaningful metric is the correlation between the observed and imputed phenotypes after adjustment for the GWAS covariates, rather than the correlation before adjustment.

These results have several implications. First, the current literature may overestimate the statistical power of GWAS after adjustment using raw correlation (e.g., the effective sample size calculated using raw correlation). Covariate-adjusted correlation (or $R^2$) should also be reported to account for the effect of GWAS covariates. Second, using GWAS covariates as predictor variables for imputation may not lead to improved ML-assisted GWAS. For example, some GWAS are adjusted for a heritable phenotype such as BMI, and using BMI to impute the phenotype for ML-assisted GWAS may not improve the GWAS. Third, the r used in `POP-GWAS` should be the residual correlation. It can be estimated directly from the residual correlation of the sample with the labeled data, or from the bivariate regression intercept of the LD score using $\hat{\beta}_{Y,j}^{\text{lab}}$ and $\hat{\beta}_{\hat{Y},j}^{\text{lab}}$.

##### 6.2 Detail

Without loss of generality, we assume that there is one covariate $U_i$ in GWAS. The derivation of multiple covariates can be easily derived with a small generalization of our results. We use $\tilde{}$ to denote the residual for variables after adjusting for covariates $U_i$. Using the Frisch-Waugh-Lovell theorem, we have

$$\tilde{Y}_i = \tilde{G}_{ij}\beta_j + u_i,$$
$$\tilde{\hat{Y}}_i = \tilde{G}_{ij}\beta_j^* + w_i.$$

This is precisely the marginal GWAS model. We denote $\tilde{r} = \mathrm{Cor}(\tilde{Y}_i, \tilde{\hat{Y}})$ as the residual correlation after adjusting for the covariate $U_i$. Using the `POP-GWAS` equation (1), we have

$$\hat{\beta}_{\mathrm{Pop},j} = \tilde{r}\frac{N_{\mathrm{unlab}}}{N_{\mathrm{unlab}} + N_{\mathrm{lab}}}\hat{\beta}^{\mathrm{unlab}}_{\hat{Y},j} + \hat{\beta}^{\mathrm{lab}}_{Y,j} - \tilde{r}\frac{N_{\mathrm{unlab}}}{N_{\mathrm{unlab}} + N_{\mathrm{lab}}}\hat{\beta}^{\mathrm{lab}}_{\hat{Y},j},$$

and the corresponding effective sample size is

$$N_{\mathrm{eff}} = \frac{N_{\mathrm{lab}}}{1 - \dfrac{\tilde{r}^2 N_{\mathrm{unlab}}}{N_{\mathrm{unlab}} + N_{\mathrm{lab}}}}.$$

Therefore, the power increase for ML-assisted GWAS depends on the residual correlation $\tilde{r}$ between the observed and imputed phenotypes after adjusting for the GWAS covariates.

### 7 Selection bias

#### 7.1 Summary

We investigated the impact of selection bias on ML-assisted GWAS, an aspect typically ignored in the literature. We considered scenarios in which phenotype missingness is not random. We found that GWAS of the observed phenotype lead to potentially biased and false positive associations. Even with perfect imputation, GWAS of imputed phenotypes also have biased false positive associations (Supplementary Figure 6). To address this issue, we further generalize `POP-GWAS` to incorporate sampling weights to address selection bias and demonstrate its statistical guarantees. Simulation results support the validity of the derivation and show that `POP-GWAS` successfully corrects for selection bias (Supplementary Figure 6).

#### 7.2 Detail

Here, we consider the impact of selection bias in ML-assisted GWAS. We use the Bernoulli indicator $R_k, k = 1, \cdots (N_{\mathrm{lab}} + N_{\mathrm{unlab}})$ to indicate whether $Y_k, k = 1, \cdots (N_{\mathrm{lab}} + N_{\mathrm{unlab}})$ were observed or not. We assume $R_k \overset{\text{i.i.d.}}{\sim} \mathrm{Bernoulli}(p_k)$ for $k = 1, \ldots, (N_{\mathrm{lab}} + N_{\mathrm{unlab}})$, which reflects selection bias. Let $q = \mathrm{pr}(R = 1) = N_{\mathrm{lab}}/(N_{\mathrm{lab}} + N_{\mathrm{unlab}})$. Suppose the true data generating process satisfies

$$\mathbb{E}(Y \mid X_j, R = 1) = \alpha_1^j + \beta_j^1 X_j, \text{ and}$$
$$\mathbb{E}(Y \mid X_j, R = 0) = \alpha_0^j + \beta_j^0 X_j.$$

Our estimand in GWAS for $j$-th SNP (defined on the population) is

$$\beta_j = q\beta_j^0 + (1-q)\beta_j^1.$$

#### 7.3 Bias of current methods

**GWAS of observed phenotype in labeled data** It is typical to construct an estimate $\widehat{\beta}_j^1$ that is unbiased for $\beta_j^1$ from the labeled sample, which could be biased for the target parameter $\beta_j$ since the bias

$$\begin{aligned}\mathbb{E}[\widehat{\beta}_j^1] - \beta_j &= \beta_j^1 - \beta_j \\ &= \beta_j^1 - q\beta_j^1 - (1-q)\beta_j^0 \\ &= (1-q)(\beta_j^1 - \beta_j^0)\end{aligned}$$

is not necessarily zero. The bias becomes zero when either $q = 1$ (all samples labeled) or $\beta_j^1 = \beta_j^0$ (missing completely at random).

The estimator of GWAS of observed phenotype in labeled data is

$$\mathrm{Cor}(X_{ij}, Y_i | R_i = 1) = \mathbb{E}[X_{ij} Y_i | R_i = 1]$$

**GWAS of imputed phenotype in unlabeled data** To investigate the bias due to selection bias, we assume that the imputation algorithm is perfect (obviously it will not be perfect). However, even if the sources of biased due to imperfect imputation is controlled, using unlabeled samples only will provide an estimate $\beta_j^0$ that is unbiased to $\beta_j$. Similar to the previous case, the resulting bias is not guaranteed to be zero,

$$\begin{aligned}\mathbb{E}(\widehat{\beta}_j^0) - \beta_j &= \beta_j^0 - \beta_j \\ &= \beta_j^0 - q\beta_j^1 - (1-q)\beta_j^0 \\ &= -q(\beta_j^1 - \beta_j^0),\end{aligned}$$

unless $q = 0$ (all samples unlabeled) or $\beta_j^1 = \beta_j^0$ (missing completely at random).

#### 7.4 Correction for selection bias in POP-GWAS

We propose POP-GWAS-IPW to correct for selection bias:

$$\hat{\beta}_{\mathrm{Pop},j}^{(\mathrm{IPW})} = r\frac{N_{\mathrm{unlab}}^{(\mathrm{IPW})}}{N_{\mathrm{unlab}}^{(\mathrm{IPW})} + N_{\mathrm{lab}}^{(\mathrm{IPW})}}\hat{\beta}_{\widehat{Y},j}^{\mathrm{unlab}\,(\mathrm{IPW})} + \hat{\beta}_{Y,j}^{\mathrm{lab}\,(\mathrm{IPW})} - r\frac{N_{\mathrm{unlab}}^{(\mathrm{IPW})}}{N_{\mathrm{unlab}}^{(\mathrm{IPW})} + N_{\mathrm{lab}}^{(\mathrm{IPW})}}\hat{\beta}_{\widehat{Y},j}^{\mathrm{lab}\,(\mathrm{IPW})},$$

where $\hat{\beta}_{\widehat{Y},j}^{\mathrm{unlab}\,(\mathrm{IPW})}$, $\hat{\beta}_{Y,j}^{\mathrm{lab}\,(\mathrm{IPW})}$, and $\hat{\beta}_{\widehat{Y},j}^{\mathrm{lab}\,(\mathrm{IPW})}$ is the inverse probability weighting (IPW) estimator for the three GWAS. $N_{\mathrm{unlab}}^{(\mathrm{IPW})}$, and $N_{\mathrm{lab}}^{(\mathrm{IPW})}$ is the effective sample size for labeled and unlabeled data due to weighted OLS.

Its corresponded standard error is

$$\begin{aligned}\mathrm{SE}(\hat{\beta}_{\mathrm{Pop},j}) &= \sqrt{\mathrm{Var}(r\frac{N_{\mathrm{unlab}}^{(\mathrm{IPW})}}{N_{\mathrm{unlab}}^{(\mathrm{IPW})} + N_{\mathrm{lab}}^{(\mathrm{IPW})}}\hat{\beta}_{\widehat{Y},j}^{\mathrm{unlab}} + \hat{\beta}_{Y,j}^{\mathrm{lab}} - r\frac{N_{\mathrm{unlab}}^{(\mathrm{IPW})}}{N_{\mathrm{unlab}}^{(\mathrm{IPW})} + N_{\mathrm{lab}}^{(\mathrm{IPW})}}\hat{\beta}_{\widehat{Y},j}^{\mathrm{lab}})} \\ &= \sqrt{(r\frac{N_{\mathrm{unlab}}^{(\mathrm{IPW})}}{N_{\mathrm{unlab}}^{(\mathrm{IPW})} + N_{\mathrm{lab}}^{(\mathrm{IPW})}})^2\,\mathrm{Var}(\hat{\beta}_{\widehat{Y},j}^{\mathrm{unlab}}) + \mathrm{Var}(\hat{\beta}_{Y,j}^{\mathrm{lab}}) + (r\frac{N_{\mathrm{unlab}}^{(\mathrm{IPW})}}{N_{\mathrm{unlab}}^{(\mathrm{IPW})} + N_{\mathrm{lab}}^{(\mathrm{IPW})}})^2\,\mathrm{Var}(\hat{\beta}_{\widehat{Y},j}^{\mathrm{lab}}) - 2r\frac{N_{\mathrm{unlab}}^{(\mathrm{IPW})}}{N_{\mathrm{unlab}}^{(\mathrm{IPW})} + N_{\mathrm{lab}}^{(\mathrm{IPW})}}\mathrm{Cov}(\hat{\beta}_{Y,j}^{\mathrm{lab}}, \hat{\beta}_{\widehat{Y},j}^{\mathrm{lab}})} \\ &\approx \sqrt{(r\frac{N_{\mathrm{unlab}}^{(\mathrm{IPW})}}{N_{\mathrm{unlab}}^{(\mathrm{IPW})} + N_{\mathrm{lab}}^{(\mathrm{IPW})}})^2\frac{1}{N_{\mathrm{unlab}}^{(\mathrm{IPW})}} + \frac{1}{N_{\mathrm{lab}}^{(\mathrm{IPW})}} + (r\frac{N_{\mathrm{unlab}}^{(\mathrm{IPW})}}{N_{\mathrm{unlab}}^{(\mathrm{IPW})} + N_{\mathrm{lab}}^{(\mathrm{IPW})}})^2\frac{1}{N_{\mathrm{lab}}^{(\mathrm{IPW})}} - 2r\frac{N_{\mathrm{unlab}}^{(\mathrm{IPW})}}{N_{\mathrm{unlab}}^{(\mathrm{IPW})} + N_{\mathrm{lab}}^{(\mathrm{IPW})}}\frac{r}{N_{\mathrm{lab}}^{(\mathrm{IPW})}}} \\ &\approx \sqrt{\frac{1}{N_{\mathrm{lab}}^{(\mathrm{IPW})}} - r^2\frac{N_{\mathrm{unlab}}^{(\mathrm{IPW})}}{(N_{\mathrm{unlab}}^{(\mathrm{IPW})} + N_{\mathrm{lab}}^{(\mathrm{IPW})})N_{\mathrm{lab}}^{(\mathrm{IPW})}}}\end{aligned}$$

Using GWAS on $y$ in labeled data as an example, the IPW GWAS is

$$\begin{aligned}\hat{\beta}_{Y,j}^{\mathrm{lab}\,(\mathrm{IPW})} &= \left(\boldsymbol{X}_{\mathrm{lab}_j}^{\mathrm{T}}\boldsymbol{W}\boldsymbol{X}_{\mathrm{lab}_j}\right)^{-1}\left(\boldsymbol{X}_{\mathrm{lab}_j}^{\mathrm{T}}\boldsymbol{W}\boldsymbol{Y}_{\mathrm{lab}}\right) \\ \mathrm{Var}\left(\hat{\beta}_{Y,j}^{\mathrm{lab}\,(\mathrm{IPW})}\right) &= \left(\boldsymbol{X}_{\mathrm{lab}_j}^{\mathrm{T}}\boldsymbol{W}\boldsymbol{X}_{\mathrm{lab}_j}\right)^{-1}\left(\boldsymbol{X}_{\mathrm{lab}_j}^{\mathrm{T}}\boldsymbol{W}\boldsymbol{D}\boldsymbol{W}\boldsymbol{X}_{\mathrm{lab}_j}\right)\left(\boldsymbol{X}_{\mathrm{lab}_j}^{\mathrm{T}}\boldsymbol{W}\boldsymbol{X}_{\mathrm{lab}_j}\right)^{-1}\end{aligned}$$

Here, $\boldsymbol{Y}_{\mathrm{lab}}$ is the phenotype vector, $\boldsymbol{X}_{\mathrm{lab}_j}$ is the $j$-th SNP vector, $\boldsymbol{W}$ is a diagonal matrix with the probability weights on the diagonal. The weights for each individual are the inverse of the propensity score: $w_i = 1/\Pr(R_i = 1|\boldsymbol{L}_i)$, where $\boldsymbol{L}_i$ is the variables that determines one's selection. $\boldsymbol{D}$ is the variance-covariance matrix for the residual, which we recommend estimating using robust standard errors. The effective sample size can then be calculated as $N_{\mathrm{lab}}^{(\mathrm{IPW})} = 1/\mathrm{Var}\left(\hat{\beta}_{Y,j}^{\mathrm{lab}\,(\mathrm{IPW})}\right)$.

### 8 Additional simulations

#### 8.1 `POP-GWAS` on Binary traits

We simulate the binary phenotype with the following model:

$$Y_i \sim \text{Binom}(1, \frac{1}{1+e^{-\log(\text{OR})}}), \text{ where } \log(\text{OR}) = -1 + G_i\beta,$$
$$Z_i = G_i\gamma + Y_i\alpha + \delta_i$$

where $Y_i$ is the phenotype, $G_i$ is the SNP, and $Z_i$ is the variable used for imputation. We first generated the SNP $G_i$ coded as 0,1,2 from Binom$(2, 0.25)$, where 0.25 is the minor allele frequency. The $\beta$ is simulated as $0, \sqrt{0.02}, \sqrt{0.04}$, and $\sqrt{0.06}$. The $\gamma$ is simulated such that $G_i$ explains 0.64% of the variance of $Z_i$, the $\delta_i$ is simulated from $\mathcal{N}(0, 0.1)$, and $\alpha$ is set to let $\text{Var}(Z_i) = 1$. All other setting remains the same as described in the Method section.

#### 8.2 `POP-GWAS` with sample overlap

We used the same settings as described in the "Methods" section, except that there were 5000 overlapping samples between the labeled and unlabeled data.

#### 8.3 `POP-GWAS` using cross-validation

We used the same setup as described in the "Methods" section, but 1) we removed the training data. 2) we employed 10-fold cross-validation to impute phenotypes in the labeled data.

#### 8.4 Correlation of the GWAS effects for top SNPs

We simulated the effects for 200 independent SNPs on observed and imputed phenotype from a mixture of multivariate normal distribution:

$$\begin{pmatrix} \beta_{\text{obs},j} \\ \beta_{\text{imp},j} \end{pmatrix} \sim (1-l)\mathcal{N}_{\text{unlab}}\left[\begin{pmatrix} 0 \\ 0 \end{pmatrix}, \begin{pmatrix} 0.0025 & 0.0025 \\ 0.0025 & 0.0025 \end{pmatrix}\right] + l\mathcal{N}_{\text{unlab}}\left[\begin{pmatrix} 0 \\ 0 \end{pmatrix}, \begin{pmatrix} 0.0025 & 0 \\ 0 & 0 \end{pmatrix}\right],$$

where $l$ represents percentages of SNPs that have effects on imputed but not observed phenotype.

We then simulate the observed phenotype in labeled data with 10,000 individuals and imputed phenotype in the unlabeled data with 150,000 individuals by

$$Y_{\text{obs}} = \sum_{j=1}^{200} X_j\beta_{\text{obs},j} + \epsilon_{\text{obs}}$$
$$Y_{\text{imp}} = \sum_{j=1}^{200} X_j\beta_{\text{imp},j} + \epsilon_{\text{imp}},$$

and conducted the GWAS on the observed and imputed phenotype, respectively.

#### 8.5 `POP-GWAS` with selection bias

We used the same settings as described in the "Methods" section, except that there are selection bias. We consider a binary variable $U_i$ that explains selection of $i$-th individual as the labeled data. In the population, $\Pr(U_i = 1) = \Pr(U_i = 0) = 0.5$, but in the labeled data $\Pr(U_i = 1|R_i = 1) = 0.3$ and $\Pr(U_i = 0|R_i = 1) = 0.7$, where $R_i$ indicates the selection in the labeled data.

When the true genetic effects in the population is 0, we simulated that phenotype $Y_i = G_i - \sqrt{6e-4} + \epsilon_i$ for individuals with $U_i = 0$ and $Y_i = G_i\sqrt{6e-4} + \epsilon_i$ for $U_i = 1$. When the true genetic effects in the population is $\beta \neq 0$, we simulated that phenotype $Y_i = G_i 0 + \epsilon_i$ for individuals with $U_i = 0$ and $Y_i = G_i 2\beta + \epsilon_i$ for $U_i = 1$. We use IPW and Huber-White SE to conduct the three GWAS and use it as input for `POP-GWAS`-IPW.

### 9 Appendix

#### 9.1 Proof of Proposition 3

*Proof.* By the unbiasedness of ordinary least squares, we have

$$\begin{aligned}\mathbb{E}[\hat{\beta}_{\text{Pop},j}] &= \mathbb{E}[c\hat{\beta}^{\text{unlab}}_{\widehat{Y},j} + \hat{\beta}^{\text{lab}}_{Y,j} - c\hat{\beta}^{\text{lab}}_{\widehat{Y},j}] \\ &= c\beta^*_j + \beta_j - c\beta^*_j \\ &= \beta_j\end{aligned}$$

□

#### 9.2 Proof of Proposition 4

*Proof.* By the consistency of ordinary least squares, we have as $N_{\text{lab}} \to \infty, N_{\text{unlab}} \to \infty$

$$\begin{aligned}c\hat{\beta}^{\text{lab}}_{\widehat{Y},j} &\xrightarrow{P} \beta^*_j \\ \hat{\beta}^{\text{lab}}_{Y,j} &\xrightarrow{P} \beta_j \\ c\hat{\beta}^{\text{lab}}_{\widehat{Y},j} &\xrightarrow{P} \beta^*_j\end{aligned}$$

By Slutsky's theorem, we have

$$\hat{\beta}_{\text{Pop},j} = c\hat{\beta}^{\text{unlab}}_{\widehat{Y},j} + \hat{\beta}^{\text{lab}}_{Y,j} - c\hat{\beta}^{\text{lab}}_{\widehat{Y},j} \xrightarrow{P} \beta_j$$

□

#### 9.3 Proof of Proposition 5

*Proof.*

$$\begin{aligned}\frac{\text{Var}(\hat{\beta}_{\text{Pop},j})}{\text{Var}(\hat{\beta}^{\text{lab}}_{Y,j})} &= \frac{\dfrac{1}{N_{\text{lab}}} - r^2 \dfrac{N_{\text{unlab}}}{(N_{\text{unlab}} + N_{\text{lab}})N_{\text{lab}}}}{\dfrac{1}{N_{\text{lab}}}} \\ &= \frac{1}{1 - r^2 \dfrac{N_{\text{unlab}}}{N_{\text{unlab}} + N_{\text{lab}}}} \leq 1\end{aligned}$$

□

#### 9.4 Proof of Theorem 1

*Proof.* By multivariate central limit theorem and asymptotic normality of ordinary least squares, we have as

$$\sqrt{N_{\text{lab}}}\left(\begin{bmatrix} c\hat{\beta}^{\text{lab}}_{\widehat{Y},j} \\ \hat{\beta}^{\text{lab}}_{Y,j} \\ c\hat{\beta}^{\text{unlab}}_{\widehat{Y},j}\end{bmatrix} - \begin{bmatrix} c\beta^*_j \\ \beta_j \\ c\beta^*_j \end{bmatrix}\right) \xrightarrow{D} \mathcal{N}_{\text{unlab}}\left(\begin{bmatrix} 0 \\ 0 \\ 0 \end{bmatrix}, \mathbf{\Sigma}, \right)$$

where

$$\mathbf{\Sigma} = \begin{bmatrix} c^2\,\text{Var}(\hat{\beta}^{\text{lab}}_{\widehat{Y},j}) & c\,\text{Cov}(\hat{\beta}^{\text{lab}}_{\widehat{Y},j}, \hat{\beta}^{\text{lab}}_{Y,j}) & \dfrac{c^2}{k}\,\text{Cov}(\hat{\beta}^{\text{lab}}_{\widehat{Y},j}, \hat{\beta}^{\text{unlab}}_{\widehat{Y},j}) \\ c\,\text{Cov}(\hat{\beta}^{\text{lab}}_{\widehat{Y},j}, \hat{\beta}^{\text{lab}}_{Y,j}) & \text{Var}(\hat{\beta}^{\text{lab}}_{Y,j}) & \dfrac{c}{k}\,\text{Cov}(\hat{\beta}^{\text{lab}}_{Y,j}, \hat{\beta}^{\text{unlab}}_{\widehat{Y},j}) \\ \dfrac{c^2}{k}\,\text{Cov}(\hat{\beta}^{\text{lab}}_{\widehat{Y},j}, \hat{\beta}^{\text{unlab}}_{\widehat{Y},j}) & \dfrac{c}{k}\,\text{Cov}(\hat{\beta}^{\text{lab}}_{\widehat{Y},j}, \hat{\beta}^{\text{unlab}}_{\widehat{Y},j}) & \dfrac{c^2}{k}\,\text{Var}(\hat{\beta}^{\text{unlab}}_{\widehat{Y},j}) \end{bmatrix}$$

By multivariate delta methods, denoting $h([x,y,z]^{\mathrm{T}}) = -x + y + z$, we have $\nabla h([x,y,z]^{\mathrm{T}}) =$ [-1, 1, 1], therefore

$$\sqrt{N_{\text{lab}}}\left( h(\begin{bmatrix} c\hat{\beta}^{\text{lab}}_{\hat{Y},j} \\ \hat{\beta}^{\text{lab}}_{Y,j} \\ c\hat{\beta}^{\text{unlab}}_{\hat{Y},j} \end{bmatrix}) - h(\begin{bmatrix} c\beta^*_j \\ \beta_j \\ c\beta^*_j \end{bmatrix}) \right) \xrightarrow{D} \mathcal{N}_{\text{unlab}}\left(0, \nabla h^{\mathrm{T}} \boldsymbol{\Sigma} \nabla h\right)$$

Since

$$h(\begin{bmatrix} c\hat{\beta}^{\text{lab}}_{\hat{Y},j} \\ \hat{\beta}^{\text{lab}}_{Y,j} \\ c\hat{\beta}^{\text{unlab}}_{\hat{Y},j} \end{bmatrix}) = \hat{\beta}_{\text{Pop},j}, h(\begin{bmatrix} c\beta^*_j \\ \beta_j \\ c\beta^*_j \end{bmatrix}) = \beta_j,$$

and

$$\begin{aligned}
\nabla h^{\mathrm{T}} \boldsymbol{\Sigma} \nabla h &= c^2 \operatorname{Var}(\hat{\beta}^{\text{lab}}_{\hat{Y},j}) + \operatorname{Var}(\hat{\beta}^{\text{lab}}_{Y,j}) + \frac{c^2}{k} \operatorname{Var}(\hat{\beta}^{\text{unlab}}_{\hat{Y},j}) - 2c \operatorname{Cov}(\hat{\beta}^{\text{lab}}_{\hat{Y},j}, \hat{\beta}^{\text{lab}}_{Y,j}) - \\
&\quad 2\frac{c^2}{k} \operatorname{Cov}(\hat{\beta}^{\text{lab}}_{\hat{Y},j}, \hat{\beta}^{\text{unlab}}_{\hat{Y},j}) + 2\frac{c}{k} \operatorname{Cov}(\hat{\beta}^{\text{lab}}_{\hat{Y},j}, \hat{\beta}^{\text{unlab}}_{\hat{Y},j}) \\
&= c^2 \operatorname{Var}(\hat{\beta}^{\text{lab}}_{\hat{Y},j}) + \operatorname{Var}(\hat{\beta}^{\text{lab}}_{Y,j}) + \frac{c^2}{k} \operatorname{Var}(\hat{\beta}^{\text{unlab}}_{\hat{Y},j}) - 2c \operatorname{Cov}(\hat{\beta}^{\text{lab}}_{\hat{Y},j}, \hat{\beta}^{\text{lab}}_{Y,j}) \\
&= V
\end{aligned}$$

Therefore,

$$\sqrt{N_{\text{lab}}} V^{-\frac{1}{2}} \left(\hat{\beta}_{\text{Pop},j} - \beta_j\right) \xrightarrow{D} \mathcal{N}_{\text{unlab}}(0, 1)$$

□

#### 9.5 Proof of Theorem 2

*Proof.* The `POP-GWAS` estimator is

$$\begin{aligned}
\hat{\beta}_{\text{Pop},j} &= r\frac{N_{\text{unlab}}}{N_{\text{unlab}} + N_{\text{lab}}} \hat{\beta}^{\text{unlab}}_{\hat{Y},j} + \hat{\beta}^{\text{lab}}_{Y,j} - r\frac{N_{\text{unlab}}}{N_{\text{unlab}} + N_{\text{lab}}} \hat{\beta}^{\text{lab}}_{\hat{Y},j}, \\
&= r\frac{1}{N_{\text{unlab}} + N_{\text{lab}}} \boldsymbol{G}^{\mathrm{T}}_{\text{unlab},j} \hat{\boldsymbol{Y}}_{\text{unlab}} + \frac{1}{N_{\text{lab}}} \boldsymbol{G}^{\mathrm{T}}_{\text{lab},j} \boldsymbol{Y}_{\text{lab}} - r\frac{N_{\text{unlab}}}{(N_{\text{unlab}} + N_{\text{lab}})N_{\text{lab}}} \boldsymbol{G}^{\mathrm{T}}_{\text{lab},j} \hat{\boldsymbol{Y}}_{\text{lab}},
\end{aligned}$$

with variance $\operatorname{Var}(\hat{\beta}_{\text{Pop},j}) = \dfrac{1}{N_{\text{lab}}} - r^2 \dfrac{N_{\text{unlab}}}{(N_{\text{unlab}} + N_{\text{lab}})N_{\text{lab}}}$

Let $\tilde{\beta}_j = \boldsymbol{C}_1 \boldsymbol{Y}_{\text{lab}} + \boldsymbol{C}_2 \hat{\boldsymbol{Y}}_{\text{lab}} + \boldsymbol{C}_3 \hat{\boldsymbol{Y}}_{\text{unlab}}$ be the other linear unbiased estimator of $\beta_j$ with

$$\begin{aligned}
\boldsymbol{C}_1 &= \frac{1}{N_{\text{lab}}} \boldsymbol{G}^{\mathrm{T}}_{\text{lab},j} + \boldsymbol{D}_1 \\
\boldsymbol{C}_2 &= -r\frac{N_{\text{unlab}}}{(N_{\text{lab}} + N_{\text{unlab}})N_{\text{lab}}} \boldsymbol{G}^{\mathrm{T}}_{\text{lab},j} + \boldsymbol{D}_2 \\
\boldsymbol{C}_3 &= r\frac{1}{N_{\text{unlab}} + N_{\text{lab}}} \boldsymbol{G}^{\mathrm{T}}_{\text{unlab},j} + \boldsymbol{D}_3
\end{aligned}$$

Here, $\boldsymbol{D}_1$, $\boldsymbol{D}_2$, and $\boldsymbol{D}_3$ are $1 \times N_{\text{lab}}$ matrix, and all of them cannot be zero at the same time (otherwise, it becomes `POP-GWAS`). By the unbiasedness of $\tilde{\beta}_j$, we have

$$\begin{aligned}
\mathbb{E}[\tilde{\beta}_j|\boldsymbol{G}_{\text{lab},j}, \boldsymbol{G}_{\text{unlab},j}] &= \mathbb{E}[\boldsymbol{C}_1\boldsymbol{Y}_{\text{lab}} + \boldsymbol{C}_2\hat{\boldsymbol{Y}}_{\text{lab}} + \boldsymbol{C}_3\hat{\boldsymbol{Y}}_{\text{unlab}}|\boldsymbol{G}_{\text{lab},j}, \boldsymbol{G}_{\text{unlab},j}] \\
&= (\frac{1}{N_{\text{lab}}}\boldsymbol{G}_{\text{lab},j}^{\text{T}} + \boldsymbol{D}_1)\boldsymbol{G}_{\text{lab},j}\beta_j + \left[-r\frac{N_{\text{unlab}}}{(N_{\text{lab}} + N_{\text{unlab}})N_{\text{lab}}}\boldsymbol{G}_{\text{lab},j}^{\text{T}} + \boldsymbol{D}_2\right]\boldsymbol{G}_{\text{lab},j}\beta_j^* \\
&\quad + (r\frac{1}{N_{\text{unlab}} + N_{\text{lab}}}\boldsymbol{G}_{\text{unlab},j}^{\text{T}} + \boldsymbol{D}_3)\boldsymbol{G}_{\text{unlab},j}\beta_j^* \\
&= (1 + \boldsymbol{D}_1\boldsymbol{G}_{\text{lab},j})\beta_j + (\boldsymbol{D}_2\boldsymbol{G}_{\text{lab},j} + \boldsymbol{D}_3\boldsymbol{G}_{\text{unlab},j})\beta_j^*
\end{aligned}$$

Since we have no assumptions between $\beta_j$ and $\beta_j^*$, $\tilde{\beta}_j$ is unbiased for $\beta_j$ if and only if $\boldsymbol{D}_1\boldsymbol{G}_{\text{lab},j} = 0$ and $\boldsymbol{D}_2\boldsymbol{G}_{\text{lab},j} + \boldsymbol{D}_3\boldsymbol{G}_{\text{unlab},j} = 0$. The varaince of $\tilde{\beta}_j$ can be written as

$$\begin{aligned}
\text{Var}(\tilde{\beta}_j|\boldsymbol{G}_{\text{lab},j}, \boldsymbol{G}_{\text{unlab},j}) &= \text{Var}(\boldsymbol{C}_1\boldsymbol{Y}_{\text{lab}} + \boldsymbol{C}_2\hat{\boldsymbol{Y}}_{\text{lab}} + \boldsymbol{C}_3\hat{\boldsymbol{Y}}_{\text{unlab}}|\boldsymbol{G}_{\text{lab},j}, \boldsymbol{G}_{\text{unlab},j}) \\
&= \boldsymbol{C}_1 \,\text{Var}\left(\boldsymbol{Y}_{\text{lab}}\right) \boldsymbol{C}_1^{\text{T}} + \boldsymbol{C}_2 \,\text{Var}\left(\hat{\boldsymbol{Y}}_{\text{lab}}\right) \boldsymbol{C}_2^{\text{T}} + \boldsymbol{C}_3 \,\text{Var}\left(\hat{\boldsymbol{Y}}_{\text{lab}}\right) \boldsymbol{C}_3^{\text{T}} + 2\boldsymbol{C}_1 \,\text{Cov}\left(\boldsymbol{Y}_{\text{lab}}, \hat{\boldsymbol{Y}}_{\text{lab}}\right) \boldsymbol{C}_2^{\text{T}} \\
&= \left(\frac{1}{N_{\text{lab}}}\boldsymbol{G}_{\text{lab},j}^{\text{T}} + \boldsymbol{D}_1\right)\left(\frac{1}{N_{\text{lab}}}\boldsymbol{G}_{\text{lab},j} + \boldsymbol{D}_1^{\text{T}}\right) \\
&\quad + \left(-r\frac{N_{\text{unlab}}}{N_{\text{lab}}(N_{\text{unlab}} + N_{\text{lab}})}\boldsymbol{G}_{\text{lab},j}^{\text{T}} + \boldsymbol{D}_2\right)\left(-r\frac{N_{\text{unlab}}}{N_{\text{lab}}(N_{\text{unlab}} + N_{\text{lab}})}\boldsymbol{G}_{\text{lab},j} + \boldsymbol{D}_2^{\text{T}}\right) \\
&\quad + \left(r\frac{1}{N_{\text{lab}} + N_{\text{unlab}}}\boldsymbol{G}_{\text{unlab},j}^{\text{T}} + \boldsymbol{D}_3\right)\left(r\frac{1}{N_{\text{lab}} + N_{\text{unlab}}}\boldsymbol{G}_{\text{unlab},j} + \boldsymbol{D}_3^{\text{T}}\right) \\
&\quad + 2r\left(\frac{1}{N_{\text{lab}}}\boldsymbol{G}_{\text{lab},j}^{\text{T}} + \boldsymbol{D}_1\right)\left(-r\frac{N_{\text{unlab}}}{N_{\text{lab}}(N_{\text{lab}} + N_{\text{unlab}})}\boldsymbol{G}_{\text{lab},j} + \boldsymbol{D}_2^{\text{T}}\right) \\
&= \frac{1}{N_{\text{lab}}} + r^2\frac{N_{\text{unlab}}^2}{(N_{\text{lab}} + N_{\text{unlab}})^2 N_{\text{lab}}} + r^2\frac{N_{\text{unlab}}}{(N_{\text{lab}} + N_{\text{unlab}})^2} - 2r^2\frac{N_{\text{unlab}}}{N_{\text{lab}}(N_{\text{lab}} + N_{\text{unlab}})} \\
&\quad - r\frac{1}{N_{\text{lab}}}\boldsymbol{G}_{\text{lab},j}^{\text{T}}\boldsymbol{D}_2^{\text{T}} - r\boldsymbol{D}_2\boldsymbol{G}_{\text{lab},j}\frac{1}{N_{\text{lab}}} + 2r\frac{1}{N_{\text{lab}}}\boldsymbol{G}_{\text{lab},j}^{\text{T}}\boldsymbol{D}_2^{\text{T}} \\
&\quad + 2r\boldsymbol{D}_1\boldsymbol{D}_2^{\text{T}} + \boldsymbol{D}_1\boldsymbol{D}_1^{\text{T}} + (\boldsymbol{D}_2\boldsymbol{D}_2^{\text{T}} + \boldsymbol{D}_3\boldsymbol{D}_3^{\text{T}}) \\
&= \text{Var}(\hat{\beta}_{\text{Pop},j}) + 2r\boldsymbol{D}_1\boldsymbol{D}_2^{\text{T}} + \boldsymbol{D}_1\boldsymbol{D}_1^{\text{T}} + \boldsymbol{D}_2\boldsymbol{D}_2^{\text{T}} + \boldsymbol{D}_3\boldsymbol{D}_3^{\text{T}}
\end{aligned}$$

Since $\boldsymbol{D}_1, \boldsymbol{D}_2$ and $\boldsymbol{D}_3$ cannot all be zero matrix, we have

$$2r\boldsymbol{D}_1\boldsymbol{D}_2^{\text{T}} + \boldsymbol{D}_1\boldsymbol{D}_1^{\text{T}} + \boldsymbol{D}_2\boldsymbol{D}_2^{\text{T}} = (\boldsymbol{D}_1 + r\boldsymbol{D}_2)(\boldsymbol{D}_1 + r\boldsymbol{D}_2)^{\text{T}} + (1 - r^2)\boldsymbol{D}_2\boldsymbol{D}_2^{\text{T}} + \boldsymbol{D}_3\boldsymbol{D}_3^{\text{T}} > 0$$

Therefore,

$$\text{Var}(\tilde{\beta}_j|\boldsymbol{G}_{\text{lab},j}, \boldsymbol{G}_{\text{unlab},j}) > \text{Var}(\hat{\beta}_{\text{Pop},j}|\boldsymbol{G}_{\text{lab},j}, \boldsymbol{G}_{\text{unlab},j}),$$

which completes the proof. □
